# The Contribution of Health Behaviors to Depression Risk across Birth Cohorts

**DOI:** 10.1101/2021.10.06.21264610

**Authors:** Maria Gueltzow, Maarten J. Bijlsma, Frank J. van Lenthe, Mikko Myrskylä

## Abstract

**Background:** More recent birth cohorts are at a higher depression risk than cohorts born in the early twentieth century. We aimed to investigate to what extent changes in alcohol consumption, smoking, physical activity and obesity, contribute to these birth cohort variations.

**Methods:** We analyzed panel data from US adults born 1916-1966 enrolled in the Health and Retirement Study (N=163,760 person-years). We performed a counterfactual decomposition analysis by combining age-period-cohort models with g-computation. This allowed us to compare the predicted probability of elevated depressive symptoms (CES-D 8 score ≥3) in the natural course to a counterfactual scenario where all birth cohorts had the health behavior of the 1945 birth cohort. We stratified analyses by sex and race/ethnicity.

**Results:** Depression risk of the 1916-1949 and 1950-1966 birth cohort would be on average 2% (-2.3 to -1.7) and 0.5% (-0.9 to -0.1) higher had they had the alcohol consumption levels of the 1945 cohort. In the counterfactual with the 1945 BMI distribution, depression risk is on average 2.1% (1.8 to 2.4) higher for the 1916-1940 cohorts and 1.8% (-2.2 to -1.5) lower for the 1950-1966 cohorts. We find no cohort variations in depression risk for smoking and physical activity. The contribution of alcohol is more pronounced for Whites than for other race/ethnicity groups, and the contribution of BMI more pronounced for women than for men.

**Conclusion:** Increased obesity levels exacerbated depression risk in recent birth cohorts in the US, while drinking patterns only played a minor role.

## Introduction

Depression is a common mental disorder and major cause of disease burden worldwide. In the US specifically, it is estimated that about 5% of the population suffers from major depression^1^, with females disproportionally affected.^2^ Depression is prevalent across all ages^1^, but is particularly high in older adults due to an accumulation of risk factors, such as multimorbidity, cognitive decline, or loneliness.^3^ Depression poses a major public health concern due to the high cost-of-illness and affects all aspects of an individual’s life, including productivity, work performance and social engagement.^4^ Additionally, depression is associated with an increased mortality risk, largely due to an increased risk of suicide.^5^

In population health, it is observed that some generations are healthier than others, independent of age and time period.^6-9^ The causes of generational differences vary, ranging from early life exposure to health determinants, such as famine, to overall variations in health behavior over the life course (e.g., smoking prevalence).^6,7,10^ Knowledge on birth cohort differences is crucial in assisting health policy making and predicting trends for future generations.^11^

Regarding depression, more recent birth cohorts have higher levels of depression, psychological distress and worse mental health compared to birth cohorts born earlier in the twentieth century.^12-18^ While some studies identify an overall increase in depression prevalence across birth cohorts^13-16^, others find an increase in depression only for birth cohorts born during or after 1935-1945.^12,18^

These cohort variations in depression risk could be partly explained by cohort differences in health behavior-related determinants of depression.^12,13,19^ Indeed, more recent birth cohorts are more likely to be obese^20^, have higher physical activity levels^21^, and decreased alcohol consumption- and smoking-related mortality than earlier born cohorts.^6,22^ Alcohol abuse, smoking and obesity increase depression risk, whereas physical activity acts as a protective factor.^23-26^

The aim of this study was to assess to which extent changes in health behavior-related determinants of depression, i.e., alcohol consumption, smoking, physical activity, and obesity, explain a the link between birth cohort and depression risk.

## Data and Methods

### Data source

For our analysis, we used the 2016 RAND HRS Longitudinal File of the Health and Retirement Study (HRS). The HRS is a nationally representative longitudinal survey based in the US and started in 1992 with biannual follow-up interviews ever since. It comprises data on over 37,000 adults over the age of 50 years and their spouses. ^27^ The HRS data is sponsored by the National Institute on Aging (grant number U01AG009740) and is conducted by the University of Michigan.

We excluded observations from wave 1, as reporting of the outcome measure changed from wave 2 onwards (eFigure S.1). Proxy respondents were excluded because the outcome measure was not administered.^28^ We furthermore excluded observations with an age below 50 and above 80 years due to data scarcity at the extremes of age. After exclusion of non-respondents and ineligible respondents, we identified missing observations for 6.2% of the outcome depressive symptoms, 0% of the key covariate birth year, 0.5-8.8% of the health-behavior variables and for up to 0.1% of the confounders (sex, race/ethnicity, education). The majority of missingness of health-behavior variables is due to alcohol consumption, which was not measured in wave 2. Exclusion of wave 2 resulted in missingness of 0.2-1.5% for health behavior variables. We therefore assumed missingness at random and performed a complete case analysis for 34,542 persons and 163,760 person-years.

### Outcome

Information on depressive symptoms was assessed with the 8-item Center for Epidemiological Studies – Depression scale (CES-D 8). The CES-D 8 includes dichotomous questions on six negative and two positive items and results in a score from 0-8 with a higher score indicating higher depressive symptomatology.^28^ We used a CES-D score of ≥3 as an indicator for elevated depressive symptoms.^28^ The CES-D 8 was validated in older adults in the US.^29^

### Measurement of exposure, health behavior and confounders

Information on the exposure birth year, age, health behaviors (alcohol consumption, smoking, physical activity, height and weight) and confounders (sex, race/ethnicity, education level) was collected through face-to-face or telephone interviews. Body mass index (BMI) was calculated based on self-reported height and weight of the respondents (weight(kg)/(height(m))^2^) and categorized into underweight (<18kg/m^2^), normal weight (18-<25 kg/m^2^), overweight (25-<30 kg/m^2^) or obese (≥ 30 kg/m^2^). We categorized smokers in current smoker or non-smoker. Alcohol consumption was reported in drinks per day and categorized into non-drinker, moderate drinker (1 drink/day for females, 1-2 drinks/day for males), heavy drinker (2-3 drinks/day for females, 3-4 drinks/day for males) or excessive drinker (≥ 4 drinks/day for females, ≥ 5 drinks/day for males). We defined physical activity as performing vigorous physical activity three or more times per week (yes/no). Education level was categorized into less than high-school degree, general education diploma, high-school graduate, some college, and college and above.

### Statistical Analysis

We performed our analyses in the total sample and stratified by sex (male/female) and by race/ethnicity (Whites/Blacks/Hispanic). To account for the oversampling of Hispanics and Blacks, we applied post-stratification weights provided by the HRS in descriptive graphs.^30^

To determine the presence of birth cohort patterns in depression, we used an age-period-cohort (APC) model to investigate the associations between birth cohort and depression. We specified the following logistic regression model to estimate the probability of elevated depressive symptoms (depr) as a function of age, period, and cohort:

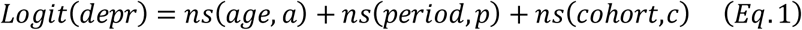

Where ns refers to a natural cubic spline function, which allows for non-linear patterns of age, period and cohort. One characteristic of APC models is the identification problem, which describes the collinearity between the age, period, and birth cohort dimension (*Age* = *Period* − *Cohort*). We addressed this problem with the Carstensen approach ^31^ and detrended the period dimension by replacing the part of the design matrix corresponding to period with a matrix with columns orthogonal to the intercept and period drift column. ^31^ The reference group was defined as age 50, 1996 for calendar year and 1945 for birth cohort.

To address our main objective of analyzing the contribution of health behaviors to depression risk across birth cohorts, we performed a counterfactual decomposition. The assumed directed acyclic graph (DAG) can be found in Figure 1. We performed the g-computation based causal decomposition for every health behavior (mediator), using an approach that is described in detail by Sudharsanan & Bijlsma. ^32^

**Figure 1.**
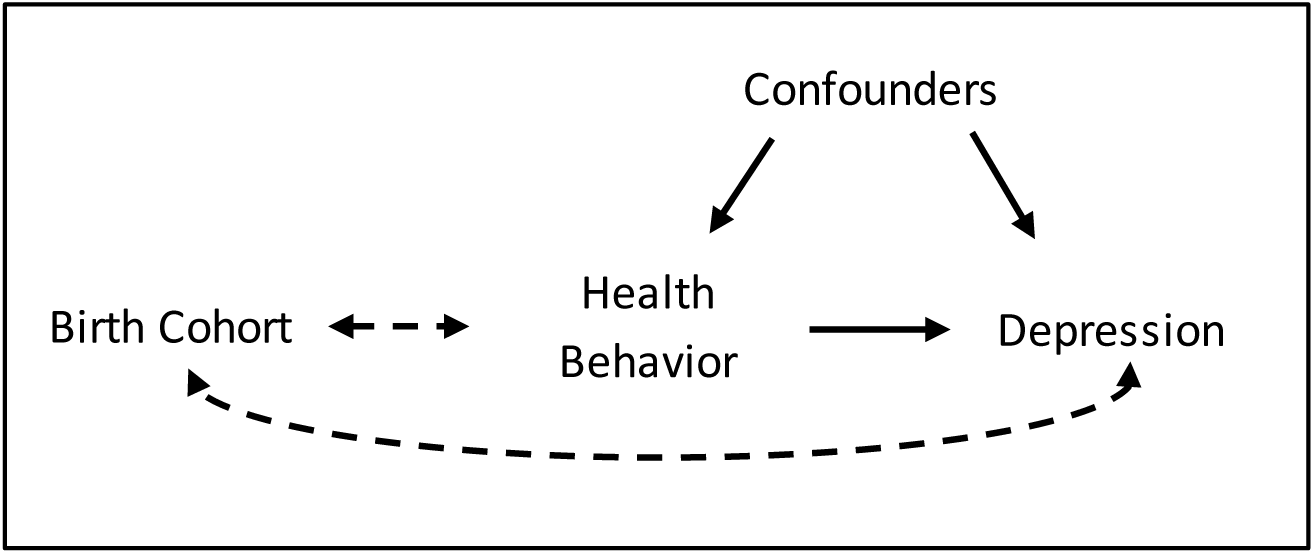
Assumed causal directed acyclic graph. Solid arrows represent causal effects, two-way dotted arrows represent associations.

First, we specified the probability of elevated depressive symptoms as our measure of interest, and the relative difference between each birth cohort and the birth cohort with the lowest probability of depressive symptoms in our sample, i.e., the 1945 birth cohort, as our contrast. Second, we fitted a logistic regression model with the probability of depressive symptoms as a function of age, period, birth cohort and the each of the health behaviors. Additionally, we modelled each respective health behavior as a function of age, period, and cohort. The model specifications of age, period and cohort were identical to the APC model specifications in *Equation 1* for both the outcome and mediator model. We used logistic regression for binary mediators and multinomial logistic regression for categorical mediators. Both the outcome and the mediator models were adjusted for education, sex and race/ethnicity in the total sample, education and race/ethnicity in sex strata and education and sex in race/ethnicity strata.

Third, we formed our natural course (nc) and counterfactual (cf) pseudo-populations. Mediator values were simulated by randomly sampling from either a binomial distribution or a multinomial distribution, depending on the mediator type, based on the predictions of the specified mediator model. We allowed the confounder distribution to vary between birth cohorts. We then predicted the probability of elevated depressive symptoms for each birth cohort based on the health behavior (mediator) and confounder distribution of each birth cohort, using the following formula:

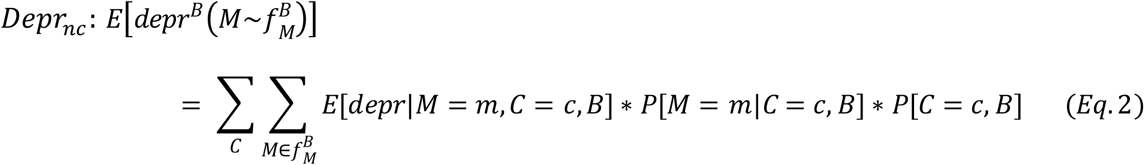

Where *depr*^*B*^ refers to the predicted probability of elevated depressive symptoms for each birth cohort (*B*), *M* indicates the mediator (alcohol consumption, smoking, physical activity, or BMI) with 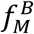 representing the distribution of mediator values by birth cohort, and *C* marks the confounding factors (sex, race/ethnicity, education).

In the natural course scenario, each cohort gets their own observed health behaviors. However, in the counterfactual scenario, each cohort’s health behaviors are taken from the 1945 birth cohort. The confounder distribution is not changed in the counterfactual:

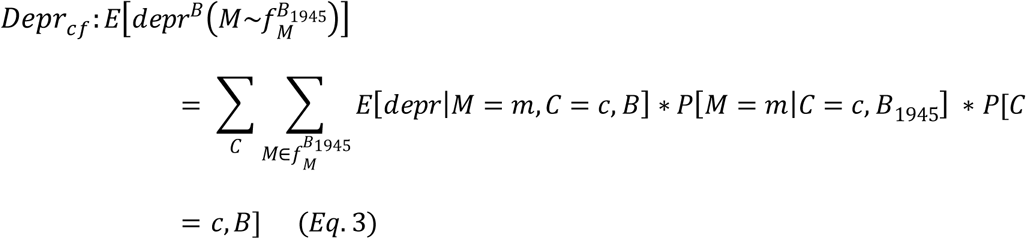

We analyzed the impact of the counterfactual by calculating the relative difference in predicted probabilities as 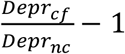. We furthermore calculated the contribution of each mediator to the difference with respect to the 1945 birth cohort as 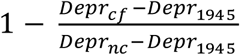.

The contribution can be interpreted as “How much does each health behavior attribute to a change in the probability of elevated depressive symptoms if every cohort had the health behavior distribution of the 1945 cohort?”.

We sampled from probability distributions, which results in Monte Carlo error. We performed Monte Carlo error reduction by repeating the simulation steps 50 times and averaging over their estimates. Furthermore, we performed 499 bootstrap iterations to compute 95% confidence intervals (see eAppendix section 4.1). We calculated the mean relative difference and contribution aggregated by birth cohorts using inverse variance weighting. ^33^

## Results

### Birth Cohort Patterns in Elevated Depressive Symptoms

Figure 2 shows the prevalence of elevated depressive symptoms by birth year for 10-year age groups. We find an overall decrease in the prevalence for cohorts 1916-1950 within age groups 60-69 and 70-80, with a more pronounced decrease in ages 70-80. In contrast, within age group 50-59, we find a steady increase in the prevalence of elevated depressive symptoms from 1936-1966. In subgroup analysis, we find a higher prevalence of elevated depressive symptoms across most cohorts in females, Hispanics, and Blacks, compared to males and Whites respectively. Information on the sample characteristics and descriptive plots by sex and race/ethnicity can be found in eAppendix section 1.

**Figure 2.**
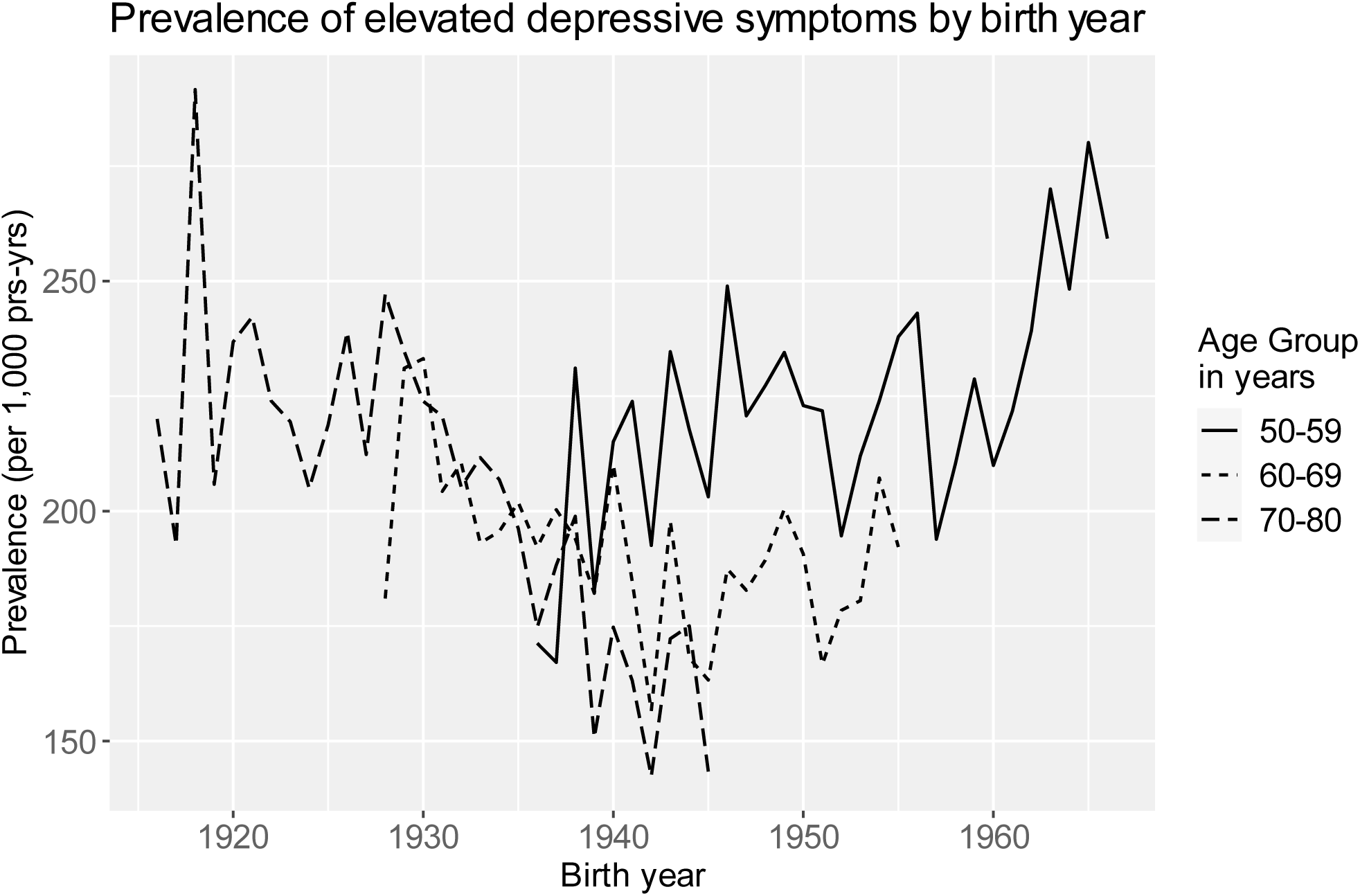
Prevalence of elevated depressive symptoms per 1,000 person-years by birth year for 10-year age groups.

We predicted the birth cohort patterns based on the specified APC model and find similar cohort patterns as in descriptive Figure 2. In short, individuals born before 1920 and after 1950 show a higher probability of elevated depressive symptoms compared to birth cohorts born between those years, when age and period are held constant. The description of age and period effects, and subgroup analysis by sex and race/ethnicity can be found in eAppendix section 1.

### Counterfactual decomposition of health behavior-related determinants of depression

The health behavior distribution for the natural course and counterfactual scenario can be found in eAppendix section 2. Our mediator models closely approximate the empirically observed health behavior patterns across cohorts. In the natural course (standardized for age and period), over cohorts, alcohol consumption and BMI increases, and smoking and physical activity show only small variations. For the counterfactual, we set the prevalence of heavy and excess drinking to 15%, smoking prevalence to 26%, prevalence of no vigorous physical activity to 44% and obesity prevalence to 28%.

Figure 3 shows the relative difference in the probability of elevated depressive symptoms between the counterfactual and natural course scenario for all health behavior factors. After the alcohol consumption distribution is set to that of birth cohort 1945, we find an average reduction in elevated depressive symptoms of 2% (−2.3 to −1.7) for birth cohorts 1916-1949 and 0.5% (−0.9 to −0.1) for cohorts 1950-1966. For physical activity and smoking, we do not find cohort variations in the relative differences in depression risk. After we counterfactually set all cohorts to have the BMI distribution of the 1945 cohort, the probability of elevated depressive symptoms increases on average by 2.1% (1.8 to 2.4) for cohorts born before 1940 and decreases on average by 1.8% (−2.2 to −1.5) for cohorts born 1945-1966.

**Figure 3.**
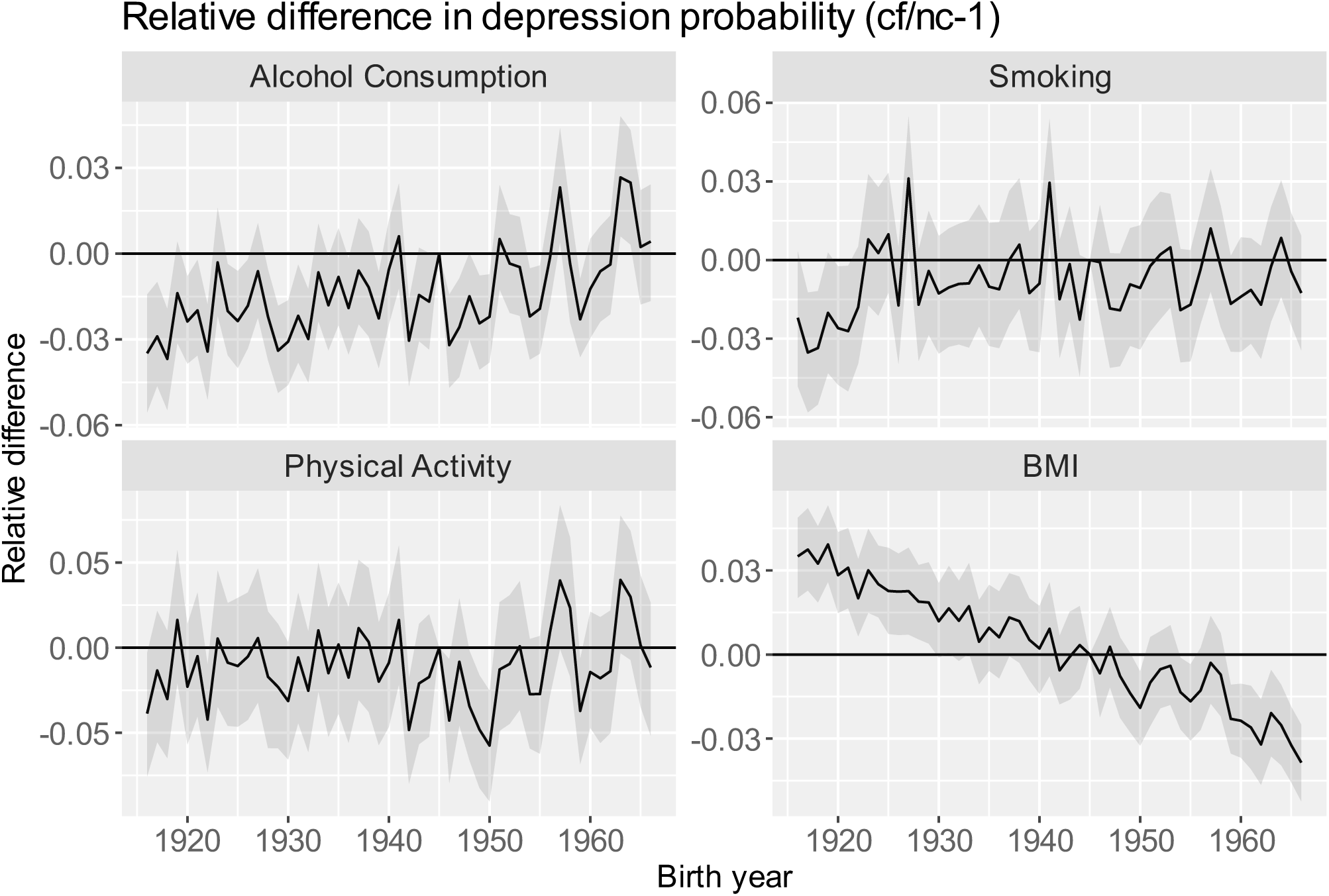
Relative difference (95% CI) between the counterfactual and natural course estimates of probability of elevated depressive symptoms by birth cohort (cf/nc-1) for each health behavior factor. Positive/negative values indicate that if the cohort had the behavioral profile of the 1945 cohort, their risk of depression would have been higher/lower.

To assess what fraction of cohort variations in depression risk are explained by health behavior, we calculated the contribution (eAppendix section 2). Alcohol consumption contributes on average 7.5% (7.5 to 7.6) to the probability of elevated depressive symptoms in cohorts born 1916-1949 and 1.9% (1.8 to 2) in cohorts born 1950-1966. We do not identify cohort patterns in the contribution of smoking and physical activity. BMI contributes on average 7.7% (−7.75 to − 7.6) to depression risk for cohorts born before 1940 and 5.5% (5.4 to 5.5) for cohorts born after 1948.

### Subgroup Analysis

The subgroup analysis revealed sex-specific differences for BMI only (Figure 4, eAppendix section 2). After counterfactually setting BMI to the 1945 distribution, the probability of elevated depressive symptoms increases for females, but not males, born before 1940 by an average of 3.5% (3.1 to 3.8) and decreases for female cohorts born after 1950 by 2.4% (−2.8 to −2).

**Figure 4.**
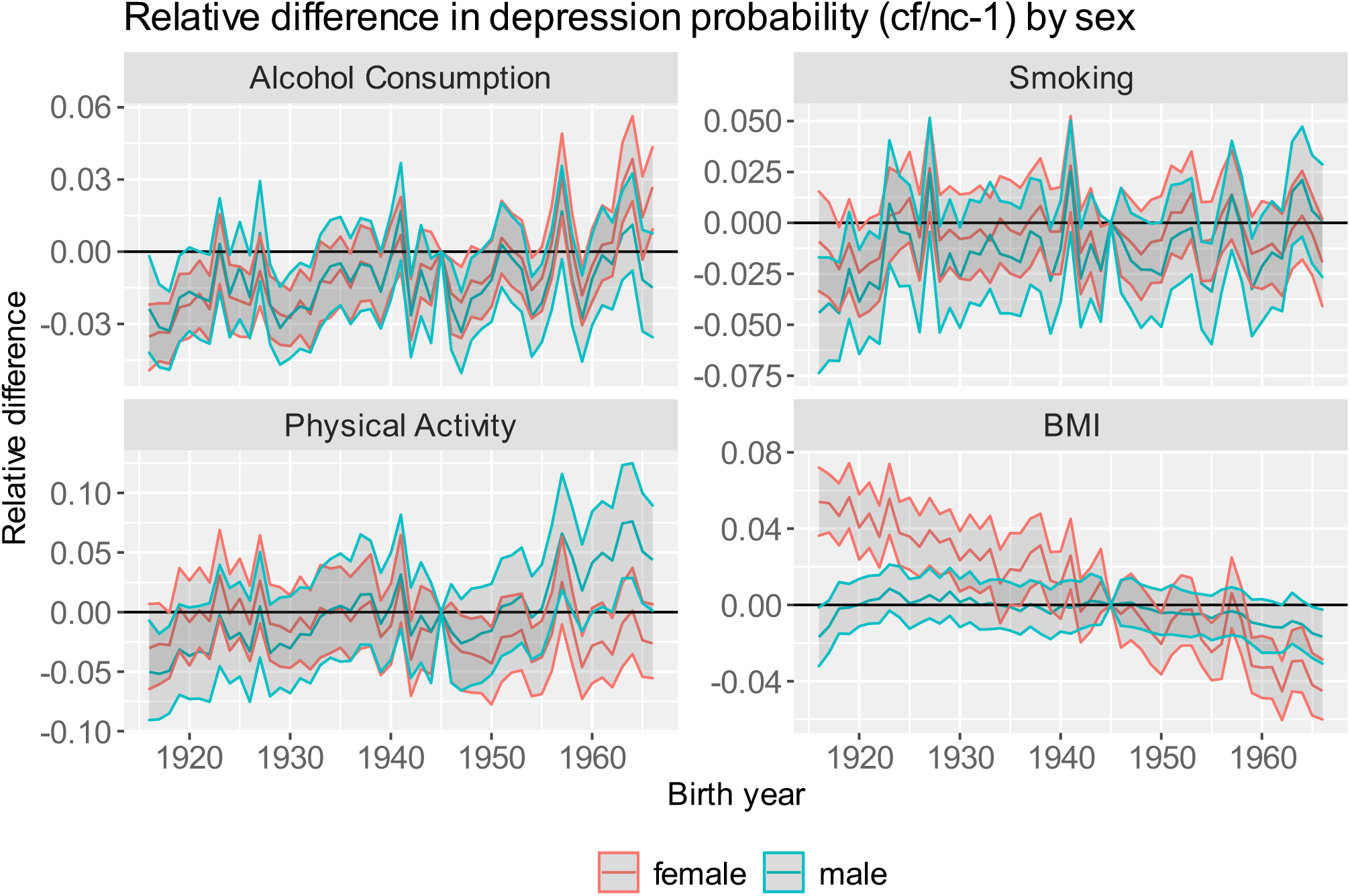
Relative difference (95% CI) between the counterfactual and natural course estimates of probability of elevated depressive symptoms by birth cohort (cf/nc-1) by sex for each health behavior factor. Relative difference is calculated as counterfactual/natural course-1. Positive values indicate that the counterfactual increases depression risk and negative values indicate that the counterfactual decreases depression risk.

For females born before 1940 and after 1950, we find average contributions of BMI of 11.5% (- 11.6 to −11.4) and 6.8% (6.7 to 6.9), respectively. We find no contributions in males.

The relative difference of alcohol consumption, physical activity and BMI differs by race/ethnicity (Figure 5). For alcohol consumption and BMI, adopting the counterfactual health behavior distributions results in a larger change in elevated depressive symptoms in Whites, compared to Hispanics and Blacks across all cohorts. In Hispanics born 1916-1935, adopting the 1945 physical activity distribution decreases the probability of elevated depressive symptoms on average by 9.8% (−10.5 to −9).

**Figure 5.**
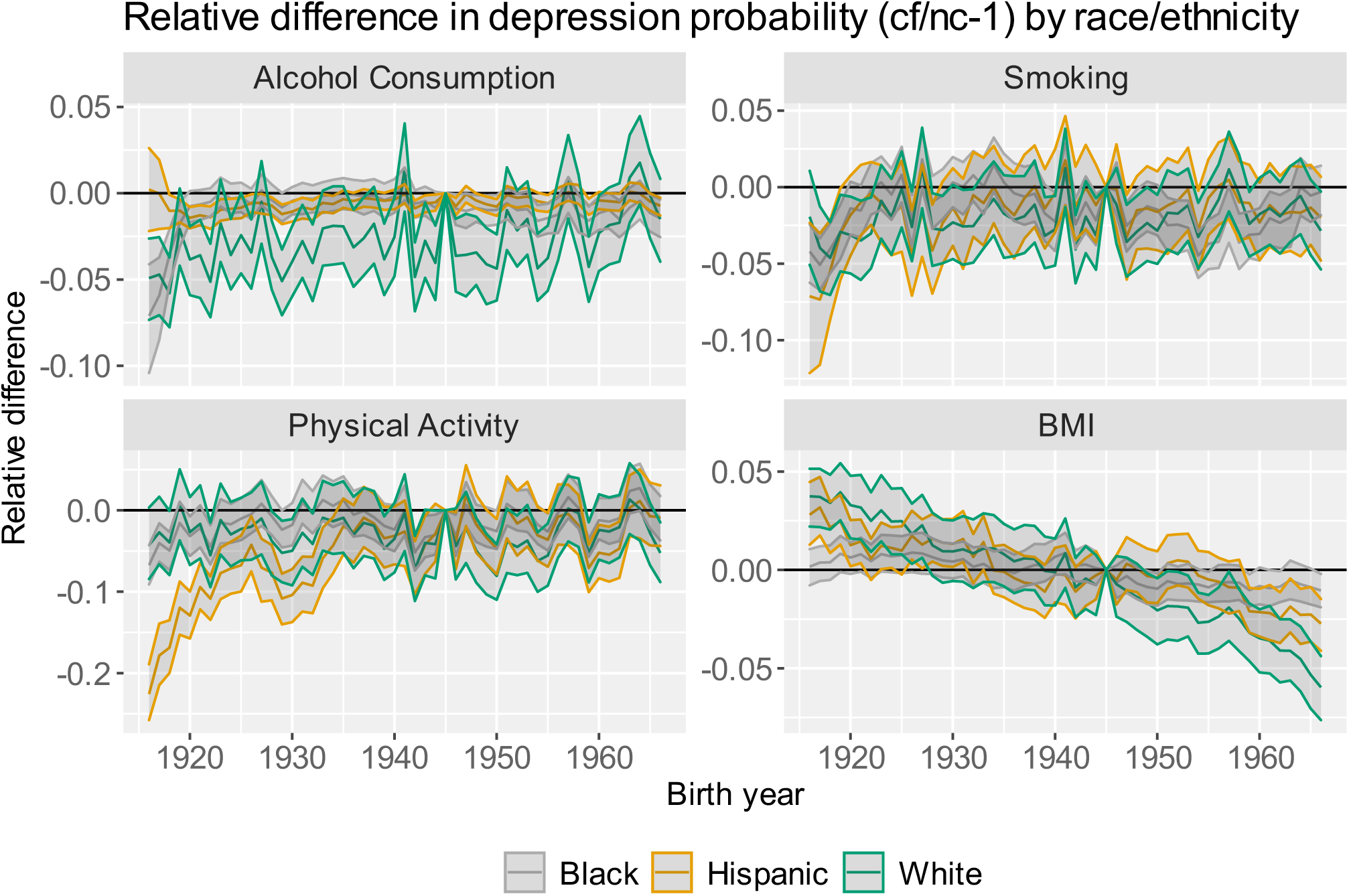
Relative difference (95% CI) between the counterfactual and natural course estimates of probability of elevated depressive symptoms by birth cohort (cf/nc-1) by race/ethnicity for each health behavior factor. Positive values indicate that the counterfactual increases depression risk and negative values indicate that the counterfactual decreases depression risk.

For alcohol consumption, we find larger contributions to depression risk in Whites, followed by Blacks and Hispanics across all cohorts (eAppendix section 2).

### Sensitivity Analysis

We repeated the analysis for smoking as a categorical variable (never/former/current smoker), which did not affect the results in any meaningful way (eAppendix section 4.2).

We assessed the sensitivity of our decomposition results to the underlying APC model constraints by calculating the relative difference for three sets of model constraints: (1) Drift (the linear time trends) is assigned to period (instead of the cohort dimension); (2) Drift is assigned to period and the reference is set to the 1996 period instead of 1945 cohort; (3) Drift is assigned to cohort and the reference is set to the 1996 period. For BMI, model constraint (2) results in a relative difference that follows the same linear downward trend as the results in Figure 4. For model constraints (1) and (3), however, we do not find a linear downward trend of BMI. We reflect on this in the discussion. The counterfactual decomposition of alcohol consumption, smoking or physical activity is not sensitive to changes in model constraints. Results from the sensitivity analysis can be found in eAppendix section 3.

## Discussion

### Summary of results

Our study finds that birth cohort differences in the probability of elevated depressive symptoms can be partly explained by health behavior. Depression risk in recent birth cohorts would have been lower for females and Whites if obesity prevalence had not increased past 1945. In contrast, alcohol consumption is a more important risk factor for depression in cohorts born before 1950 than in more recent birth cohorts. We find alcohol consumption to be a stronger contributor to depression risk in Whites compared to Hispanics and Blacks across all cohorts. We do not find evidence for contributions of smoking and physical activity.

### The link between birth cohort, health behavior and depression

Our study shows that birth cohort differences in depression risk might be partly explained by health behavior. For BMI, recent cohorts benefit from the counterfactual scenario with an average decrease in the probability of elevated depressive symptoms of 2% after the prevalence of obesity is reduced by approximately 7%-points and reassigned to normal weight and overweight. These results suggest that particularly obesity is a driver of depression risk, with higher depression in more obese populations.

Sex-stratified analyses show that the contribution of obesity to depression risk is strong for women but negligible for men across most birth cohorts. This is likely due to cohort patterns of obesity being more pronounced in females^20,34^ and that obesity increases depression risk for females but not for males.^35^ Therefore, mental health of females may have particularly suffered from the increase in obesity levels in the US.

The sensitivity analysis in which drift was assigned to period rather than cohort showed that a large part of the trend in the contribution of BMI to depression risk is attributed to the overall time trend (eAppendix section 3). Previous research indicates that environments are becoming more obesogenic over time^36^, which is indicative of period effects. Reither et al.^34^ on the other hand find cohort patterns independent of period and age, with recent cohorts being more susceptible to obesity. Hence, recent birth cohorts might be more strongly affected by increases in obesogenic forces than cohorts born earlier in the twentieth century. We interpret this as evidence for a combined effect of period and cohort. Regardless, the conclusion remains the same: over time, as the population became more obese, the prevalence of elevated depressive symptoms in recent birth cohorts is on average 5.5% attributable to this increase.

For alcohol consumption, we find that, in cohorts born before 1950, the counterfactual scenario decreases the number of non-drinkers and excess drinkers on average by 4%-points and 0.3%-points, respectively, and assigns them to either moderate or heavy drinkers. This leads to an average decrease in the probability of elevated depressive symptoms of 2%. These results imply that lower percentages of non-drinkers in the population decrease depression risk. This might be partially explained by the fact that non-drinkers are more likely to be older and in turn suffer from chronic conditions or take medication that require them to abstain from alcohol.^37^ Furthermore, non-drinkers are more likely to have a smaller social network^37^, which may in turn increase their depression risk.

For physical activity and smoking, we do not find a consistent mediating effect on birth cohort patterns of depression. These null findings are explained by the small variation of the mediator distribution between the estimated natural course and counterfactual scenario, rather than no effect of these mediators on depression risk (eAppendix section 2.2,2.3.).

### Age-Period-Cohort model results

Our analysis suggests that birth cohorts 1916-1920 and 1946-1966 experience an increased probability of elevated depressive symptoms compared to birth cohorts 1921-1945, independent of age and period. These results are partly in line with current literature that found recent birth cohorts to experience a higher risk of depression.^12-18^ In contrast, we also identified a higher depression risk in cohorts born in the early twentieth century. Keyes et al.^19^ identified a similar trend in cohort patterns of psychological distress in US adults for cohorts born 1912-1975, with increased psychological distress for most recent cohorts and cohorts born in the early twentieth century. Hence, our results are in line with the results by Keyes et al. for the cohorts available in our HRS sample (1916-1966).

### Evaluation of data and methods

Our sample is drawn from the Health and Retirement study, which is representative of US older adults.^27^ Since the HRS is a longitudinal survey, we investigated the possibility of panel attrition and panel conditioning bias^38^ and conclude that our results could be affected by differential panel attrition (eAppendix section 4.3); more depressed individuals might be more likely to leave the study. This could lead to an underestimation of depression prevalence within birth cohorts and in turn might lead to a misestimation of the relative difference and contribution.

Our outcome variable measures the presence of elevated depressive symptoms, which can be interpreted as an increased risk for depression rather than a direct measure of major depressive disorder.^39^ We may, therefore, overestimate the number of individuals suffering from a depressive disorder. However, populations with elevated depressive symptoms can be considered a target for primary prevention of depression.

Our APC model specification follows the Carstensen approach to address the identification problem. First, in this approach the drift (the linear time trend) is assigned to either cohort or period, which is a choice that is not empirically testable. We include the drift into the cohort dimension with the rationale that depression is more likely to be an accumulation of experiences shared by birth cohorts over their development, rather than a period effect.^40,41^ Second, our sensitivity analysis reveals that the results of the counterfactual decomposition of BMI cannot be accounted to non-linear cohort or period effects, but rather to the drift. This highlights an important limitation of the APC model: The current approaches that address the identification problem are able to attribute non-linear effects to each dimension, but not the total effects.

Our causal decomposition method relies on several assumptions. Most prominent of these is the assumption of no-unmeasured confounding.^32^ An advantage of causal decomposition relative to causal mediation analysis is that the exposure-mediator and exposure-outcome pathways cannot be confounded because the exposure is not a true ‘cause’.^32^ Regarding our mediator-outcome pathway, unmeasured confounding might be present as a result of social factors. We used education level as a proxy to control for social factors, but residual confounding cannot be ruled out. Additionally, because education might act as a mediator between health behavior and depression, including education as a confounder of the link between health behavior and depression might result in an underestimation of the contribution of health behavior.

Another point of interest is the potential bi-directionality of the relationship between the mediators and the outcome.^25,42-45^ For example, while alcohol consumption can be a cause of depression, depression may also cause increased alcohol consumption. We did not model this bidirectionality, which may result in an overestimation of the causal effect of the mediators on the outcome. This bidirectionality can be modelled with lag effects or the longitudinal g-formula. This would additionally allow to account for possible long-term effects of health behaviors on depression risk, though we argue that most of the long-term effect of health behavior is captured by present health behavior. While including lag effects would allow for a more accurate causal effect estimation, this would have resulted in exclusion of the oldest and youngest birth cohorts in our analysis.

## Conclusion

To our knowledge, this is the first study to investigate possible causes of birth cohort variation with the use of models embedded in the causal inference (potential outcomes) framework, specifically g-computation. Recent birth cohorts are at a higher depression risk than cohorts born in the early twentieth century and we provide insight into reasons for these birth cohort variations: BMI contributes on average 5.5% to depression risk in recent cohorts, whereas alcohol consumption is a more important risk factor in cohorts born before 1950, with average contributions of 7.5%. Our results highlight that particularly recent female cohorts could benefit from public health interventions targeted at obesity levels, which has the added benefit of decreasing depression risk in this group. Future research should investigate causes of generational differences in depression risk in cohorts born after 1966 and consider the bidirectionality between health behavior factors and depression.

## Data Availability

The Health and Retirement Study data is accessible at (http://hrsonline.isr.umich.edu/). The R code for reproducing our analysis can be requested from the corresponding author.

