## Supplementary Material for "The Contribution of Health Behaviors to Depression Risk across Birth Cohorts"

#### Flow Chart

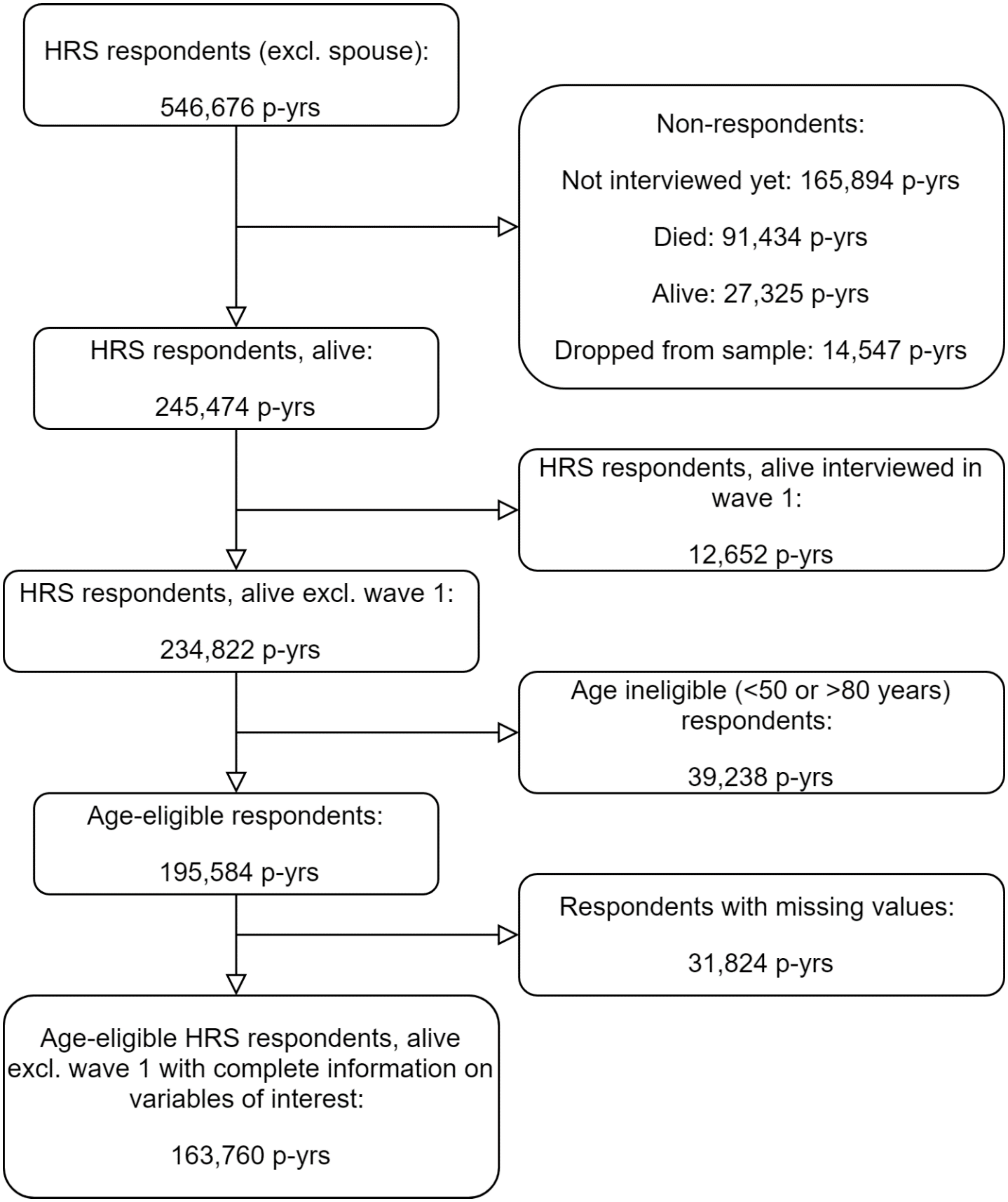

*Figure S. 1Flowchart of sample selection. p-yrs: person-years.*

### Section 1: Birth Cohort Patterns in Elevated Depressive Symptoms

#### Sample Characteristics

*Table S. 1 Sample Characteristics in total sample and stratified by sex and race/ethnicity*

| | | Total | Females | Males | White | Hispanic | Black |
|---|---|---|---|---|---|---|---|
| **N person-years** | | 163760 | 95451 | 68309 | 125446 | 5699 | 28047 |
| ***outcome*** | | | | | | | |
| **elevated depressive symptoms** | yes N(%) | 35,788 (21.85) | 23,907 (25.05) | 11,881 (17.39) | 25,056 (19.97) | 1,820 (31.94) | 7,746 (27.62) |
| ***Exposure*** | | | | | | | |
| **Age** | mean (sd) | 64.68 (8.21) | 64.55 (8.3) | 64.86 (8.08) | 65.32 (8.2) | 60.4 (7.1) | 63.14 (7.91) |
| **Period** | mean (sd) | 2006 (6) | 2006 (6) | 2006 (6) | 2006 (6) | 2010 (6) | 2007 (6) |
| **Birth cohort** | mean (sd) | 1942 (10) | 1942 (10) | 1941 (10) | 1940 (10) | 1949 (9) | 1944 (11) |
| ***Mediators*** | | | | | | | |
| **BMI N(%)** | underweight | 1,455 (0.89) | 1,166 (1.22) | 289 (0.42) | 1,133 (0.9) | 30 (0.53) | 237 (0.85) |
| | normal | 46,892 (28.63) | 30572 (32.03) | 16,320 (23.89) | 38,564 (30.74) | 1,173 (20.58) | 5,579 (19.89) |
| | overweight | 62,484 (38.16) | 31,515 (33.02) | 30,969 (45.35) | 48,654 (38.78) | 2,314 (40.6) | 9,882 (35.23) |
| | obese | 52,929 (32.32) | 32,198 (33.73) | 20,731 (30.35) | 37,095 (29.57) | 2,182 (38.29) | 12,349 (44.03) |

| | | | | | | | |
|---|---|---|---|---|---|---|---|
| **Alcohol Consumption N(%)** | non-drinker | 105,684 (64.54) | 68,333 (71.59) | 37,351 (54.68) | 77,835 (62.05) | 3,990 (70.01) | 20,473 (73) |
| | moderate drinker | 34,865 (21.29) | 14,307 (14.99) | 20,558 (30.1) | 29,438 (23.47) | 723 (12.69) | 4,003 (14.27) |
| | heavy drinker | 18,693 (11.41) | 11,449 (11.99) | 7,244 (10.6) | 14,902 (11.88) | 637 (11.18) | 2,824 (10.07) |
| | excessive drinker | 4,518 (2.76) | 1,362 (1.43) | 3,156 (4.62) | 3,271 (2.61) | 349 (6.12) | 747 (2.66) |
| **Smoking** | no N(%) | 137,277 (83.83) | 80,755 (84.6) | 56,522 (82.74) | 106,492 (84.89) | 4,803 (84.28) | 22,215 (79.21) |
| **Vigourous physical activity** | yes N(%) | 74,389 (45.43) | 38,002 (39.81) | 36,387 (53.27) | 58,584 (46.7) | 2,585 (45.36) | 11,079 (39.5) |
| ***Confounders*** | | | | | | | |
| **gender** | Female N(%) | 95451(58.29) | - | - | 72,032 (57.42) | 3,181 (55.82) | 17,672 (63.01) |
| **race/ethnicity N(%)** | White | 125,446 (76.6) | 72,032 (75.46) | 53,414 (78.19) | - | - | - |
| | Hispanic | 5,699 (3.48) | 3,181 (3.33) | 2,518 (3.63) | - | - | - |
| | Black | 28,047 (17.13) | 17,672 (18.5) | 10,375 (15,19) | - | - | - |
| | Other | 4,568 (2.79) | 2,566 (2.69) | 2,002 (2,93) | - | - | - |
| **education level N(%)** | Limited high-school | 33,160 (20.25) | 19,873 (20.82) | 13,287 (19.45) | 21,155 (16,86) | 2,779 (48,76) | 8,353 (29.78) |
| | GED | 8,155 (4.98) | 4,342 (4.55) | 3,813 (5.58) | 6,080 (4.85) | 300 (5.26) | 1,508 (5.38) |

| | | | | | | |
|---|---|---|---|---|---|---|
| High school graduate | 49,373 (30.15) | 31,164 (32.65) | 18,209 (26.66) | 39,793 (31.72) | 1,067 (18.72) | 7,606 (27.2) |
| Some college | 37,911 (23.15) | 22,559 (23.63) | 15,352 (22.47) | 29,050 (23.16) | 1,053 (18.48) | 6,735 (24.01) |
| College and above | 35,161 (21.47) | 17,513 (18.35) | 17,648 (25.84) | 29,368 (23.41) | 500 (8.77) | 3,845 (13.71) |

#### Descriptive Plots

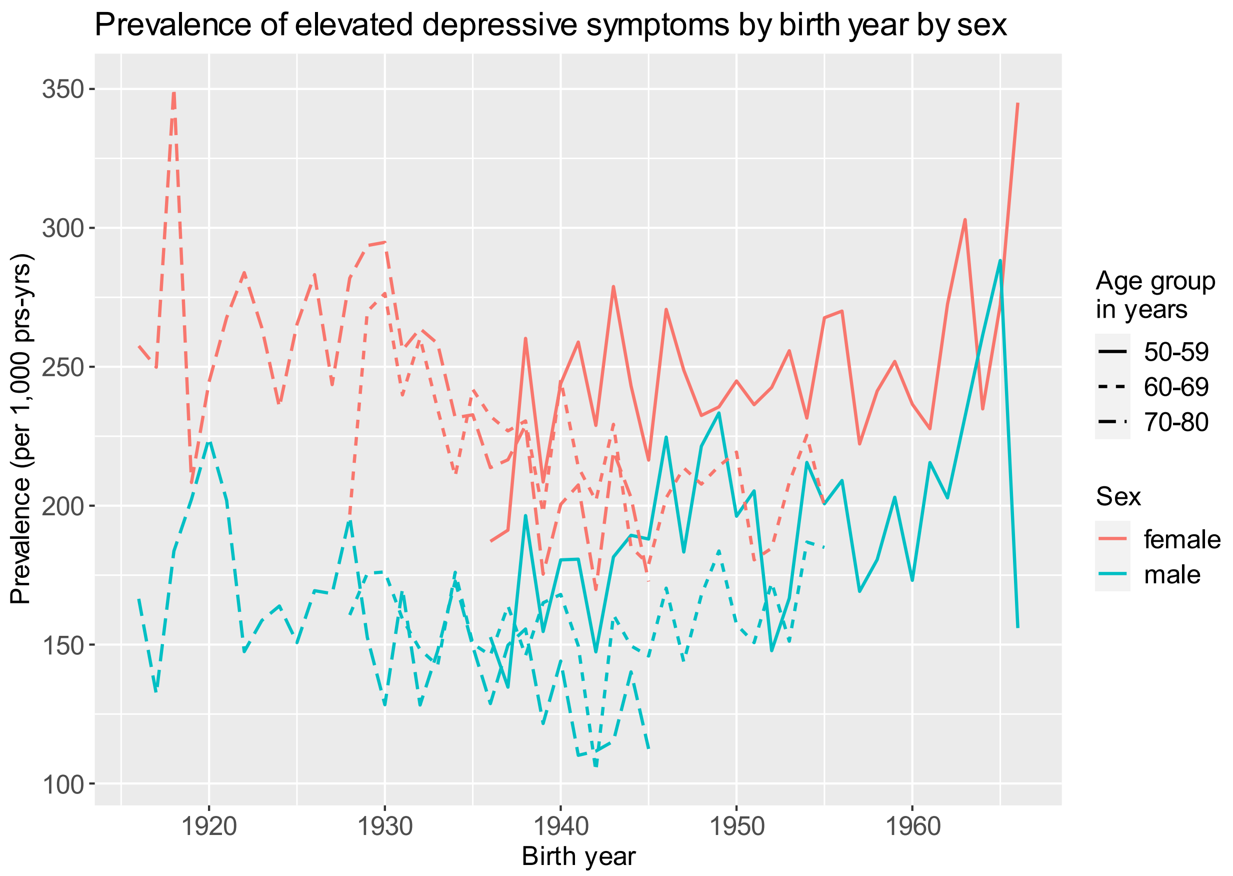

*Figure S. 2 Prevalence of elevated depressive symptoms per 1,000 person-years by birth year and sex for 10-year age groups.*

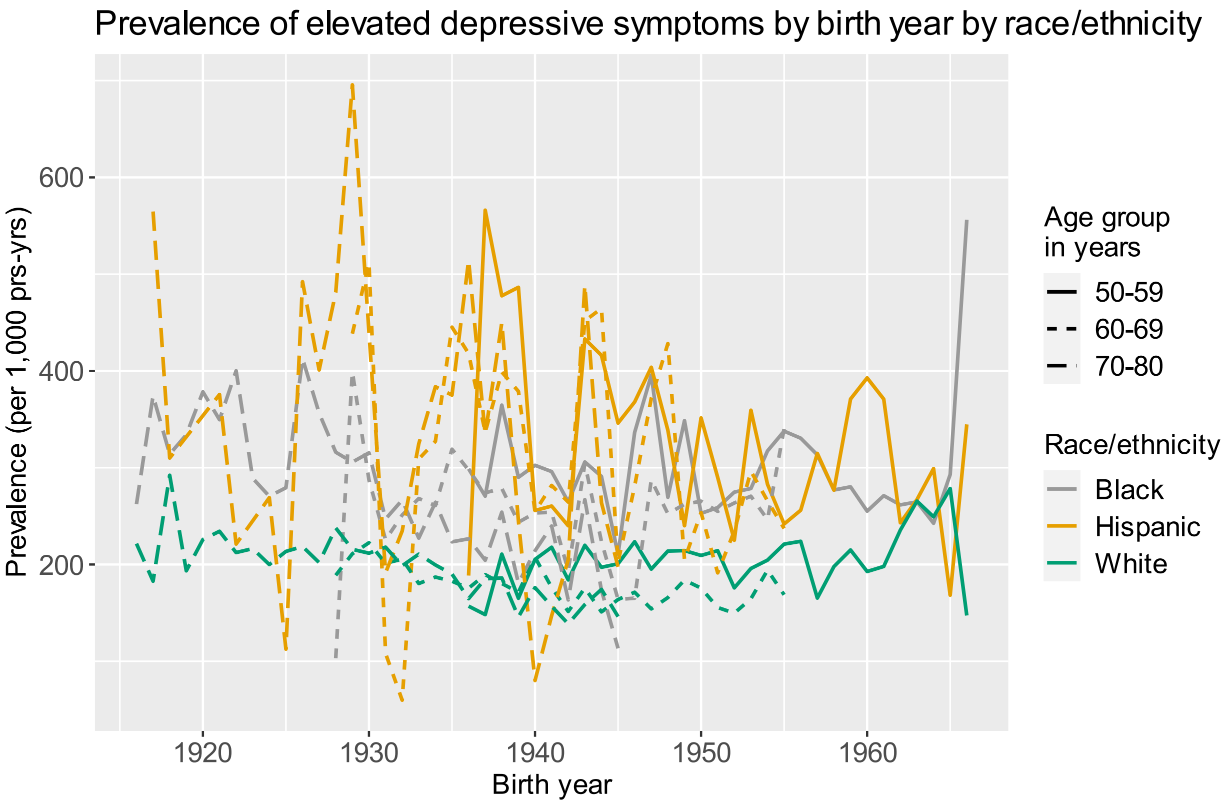

*Figure S. 3 Prevalence of elevated depressive symptoms per 1,000 person-years by birth year and race/ethnicity for 10-year age groups.*

#### Age-Period-Cohort Results

The results of the APC analysis are shown in Figure S.2. The probability of suffering from elevated depressive symptoms differs by age for the reference birth year 1945 and period 1996. The probability of elevated depressive symptoms increases between age 50 and 60, then decreases down to 14.8% (95%CI: 14-15.7) at age 70 and increases after. When stratified by sex (Figure S.3), females show a higher risk of elevated depressive symptoms from their mid-50s compared to males. The period effects show a similar distribution to the age-standardized prevalence rate of elevated depressive symptoms. The period effects show no sex-specific differences. The association between birth cohort and the probability of elevated depressive symptoms follows a u-shaped association, when age and period are kept constant at 50 years 1996, respectively. Individuals born before 1920 and after 1950 show a higher probability of elevated depressive symptoms compared to the birth cohorts born between those years. We do

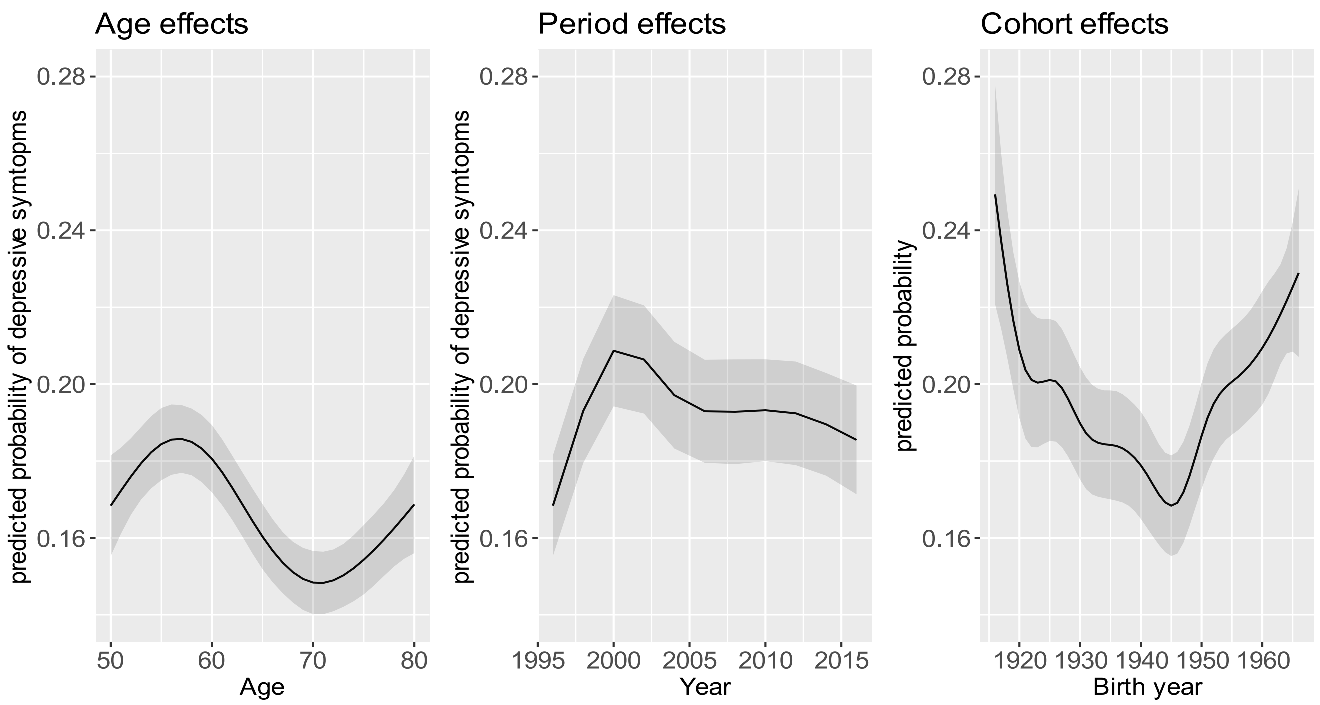

*Figure S. 4 Age-period-cohort model: Predicted age, period and cohort effects on the predicted probability of elevated depressive symptoms. 1994-2016, ages 50-80. Reference groups: Age 50, Period 1996, Birth cohort 1945.*

not observe differences by sex. The stratification by race/ethnicity revealed that Black populations and Hispanics born after 1919 experience higher probabilities of elevated depressive symptoms than White populations (Figure S.4).

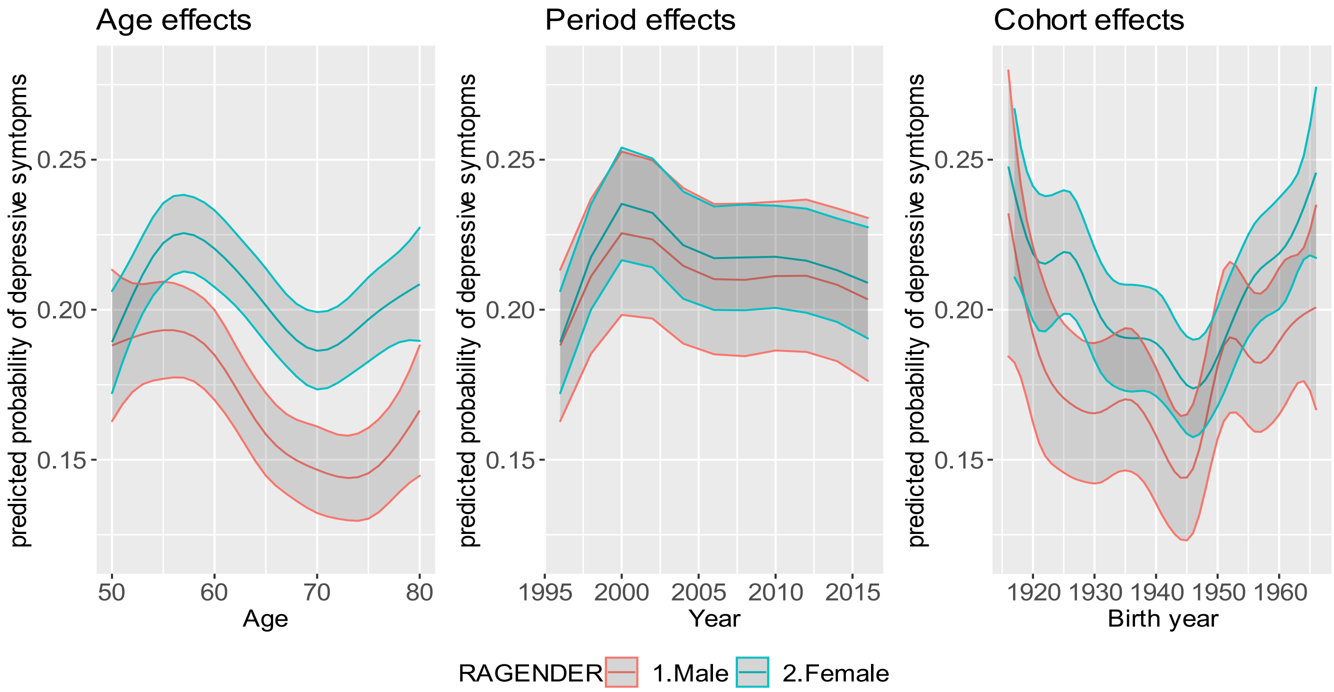

*Figure S. 5 Age-period-cohort model: Predicted age, period and cohort effects on the predicted probability of elevated depressive symptoms by sex. 1994-2016, ages 50-80. Reference groups: Age 50, Period 1996, Birth cohort 1945.*

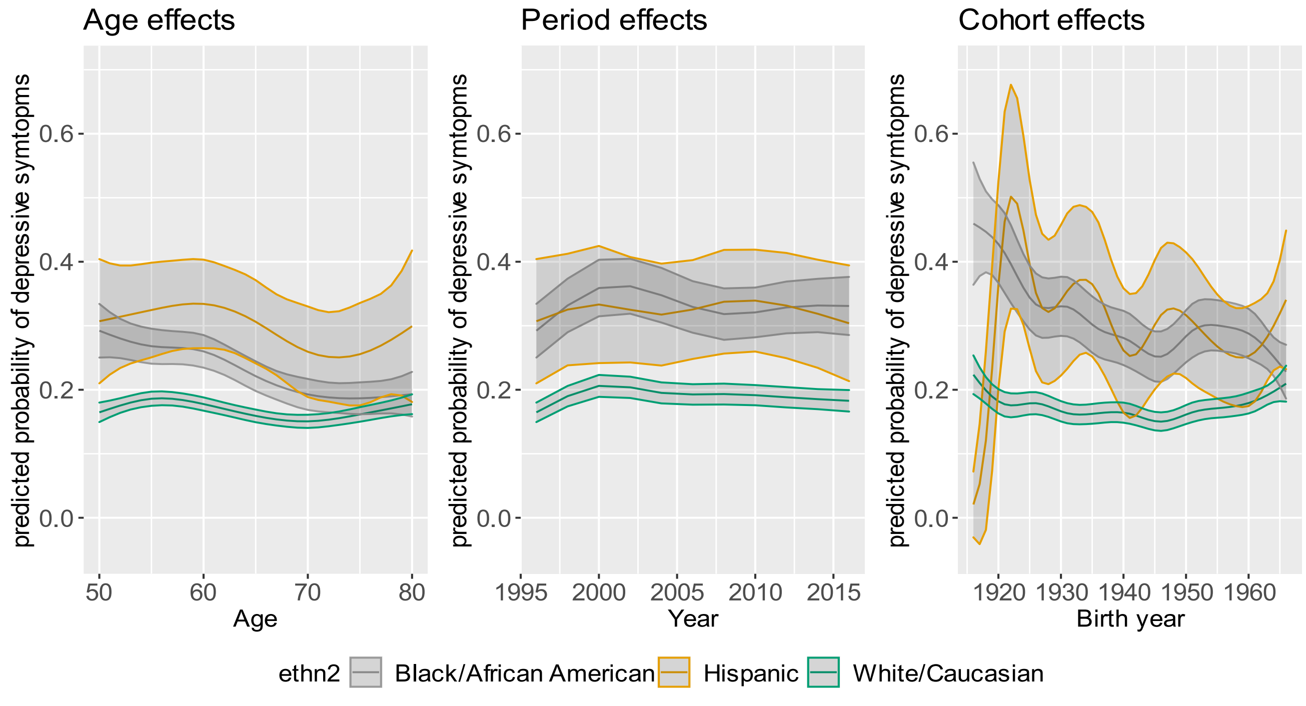

*Figure S. 6 Age-period-cohort model: Predicted age, period and cohort effects on the predicted probability of elevated depressive symptoms by race/ethnicity. 1994-2016, ages 50-80. Reference groups: Age 50, Period 1996, Birth cohort 1945.*

#### Section 2: Counterfactual Decomposition

##### Section 2.1: Alcohol Consumption

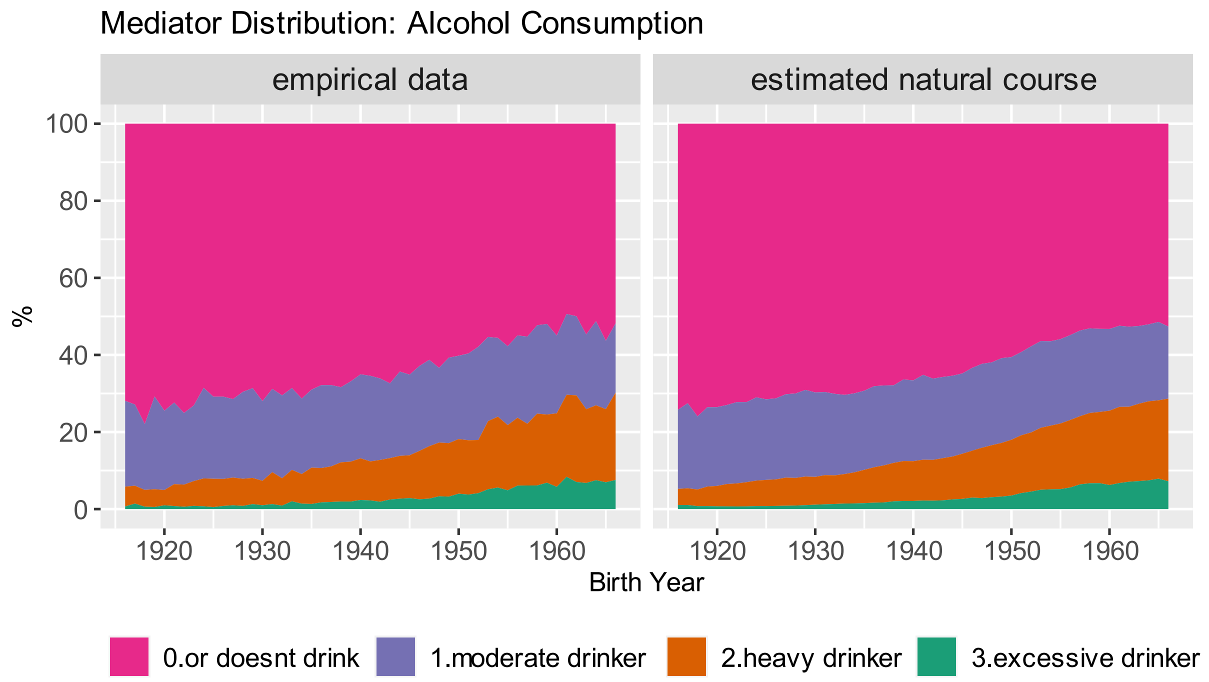

*Figure S. 7 Natural course distribution of smoking by birth cohort. "True natural course" shows the descriptive distribution as observed in the data. "Estimated natural course" is estimated based on the mediator model without holding age and period constant.*

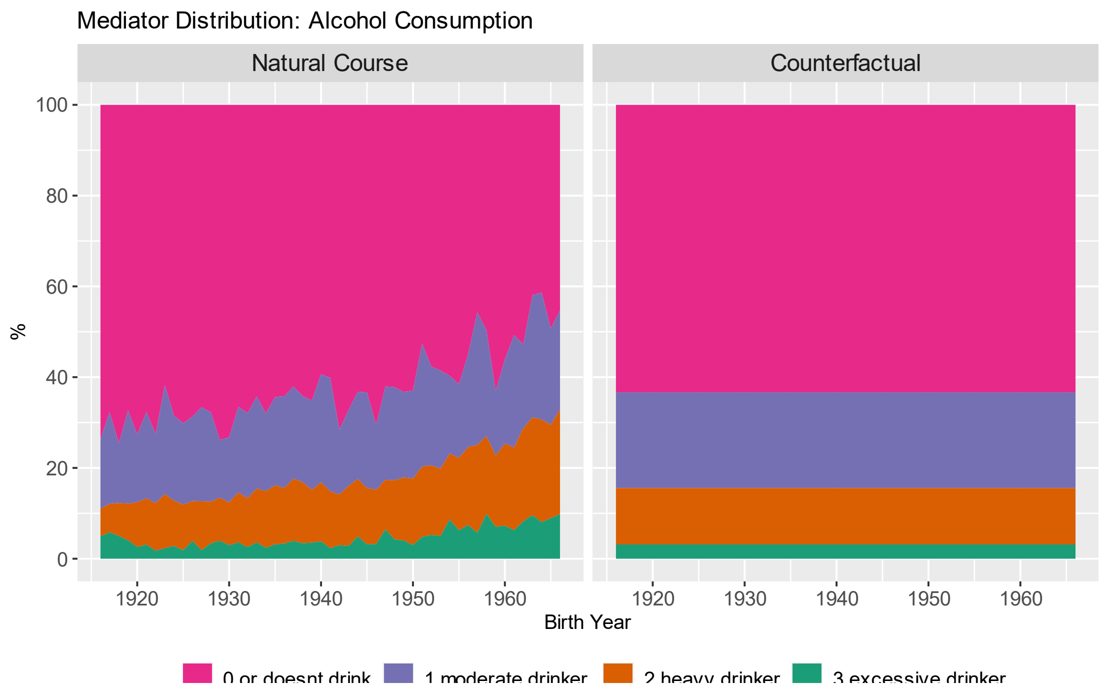

*Figure S. 8 Estimated distribution of alcohol consumption categories in natural course and counterfactual scenario by birth year. Age and period are held constant at age 50 and 1996 respectively.*

*Table S. 2 Predicted Probability of elevated depressive symptoms in natural course and counterfactual scenario, relative difference, and contribution by birth cohort. All estimates are presented with 95% confidence intervals. Positive contributions imply that the counterfactual decreases depression risk and negative contributions imply that the counterfactual increases depression risk*

| | **Probability of elevated depressive symptoms (% (95%CI))** | | **% relative difference** | **% contribution** |
|---|---|---|---|---|
| **Birth Cohort** | Natural Course Scenario | Counterfactual Scenario | | |
| **1916** | 20.24% (18.62-22.2) | 19.53% (17.98-21.28) | -3.49% (-5.55--1.43) | 11.2% (4.49-19.12) |
| **1917** | 19.47% (18.19-21.12) | 18.9% (17.64-20.41) | -2.89% (-4.62--0.99) | 10.08% (3.35-16.69) |
| **1918** | 22.29% (21.03-23.86) | 21.47% (20.23-22.92) | -3.68% (-5.47--1.93) | 9.81% (5.06-14.38) |
| **1919** | 16.43% (15.41-17.64) | 16.2% (15.23-17.35) | -1.38% (-3.12-0.42) | 8.87% (-2.72-22.35) |
| **1920** | 19.28% (18.05-20.54) | 18.82% (17.61-20.05) | -2.37% (-3.85--0.79) | 8.6% (3.01-14.34) |
| **1921** | 17.33% (16.27-18.56) | 16.98% (15.91-18.21) | -1.98% (-3.56--0.21) | 9.99% (1.16-19.46) |
| **1922** | 19.95% (18.75-21.18) | 19.26% (18.11-20.48) | -3.42% (-5.11--1.79) | 11.38% (5.97-17.27) |
| **1923** | 14.95% (14.02-15.94) | 14.9% (14.02-15.87) | -0.31% (-2.17-1.6) | 5.02% (-44.06-45.65) |
| **1924** | 18.33% (17.24-19.38) | 17.96% (16.92-19.06) | -2.01% (-3.55--0.36) | 8.47% (1.55-15.13) |
| **1925** | 16.1% (15.12-17.09) | 15.72% (14.81-16.7) | -2.36% (-4--0.61) | 17.49% (4.27-35.06) |
| **1926** | 18.2% (17.16-19.28) | 17.87% (16.89-18.89) | -1.82% (-3.31--0.21) | 7.99% (0.81-14.88) |
| **1927** | 14.79% (13.89-15.73) | 14.7% (13.8-15.66) | -0.61% (-2.23-1.07) | 10.84% (-60.45-97.23) |
| **1928** | 18.67% (17.65-19.71) | 18.25% (17.32-19.29) | -2.2% (-3.73--0.55) | 8.69% (2.09-14.99) |
| **1929** | 20.67% (19.53-21.87) | 19.97% (18.92-21.13) | -3.39% (-4.87--1.82) | 10.57% (5.58-15.24) |
| **1930** | 19.9% (18.82-21.14) | 19.29% (18.26-20.38) | -3.08% (-4.59--1.63) | 10.12% (5.46-15.48) |
| **1931** | 16.72% (15.73-17.79) | 16.36% (15.42-17.34) | -2.17% (-3.81--0.38) | 13.1% (2.73-23.8) |

| | | | | |
|---|---|---|---|---|
| **1932** | 17.73% (16.71-18.84) | 17.2% (16.26-18.28) | -2.98% (-4.51--1.24) | 14.21% (6.04-21.97) |
| **1933** | 14.43% (13.6-15.38) | 14.34% (13.5-15.29) | -0.65% (-2.36-1.05) | 13.91% (-239.89-206.29) |
| **1934** | 15.95% (15.03-17.02) | 15.67% (14.75-16.78) | -1.8% (-3.38--0.11) | 14.17% (1.03-31.31) |
| **1935** | 15.1% (14.19-16.09) | 14.97% (14.07-15.99) | -0.81% (-2.5-0.89) | 10.45% (-18-53.29) |
| **1936** | 16.1% (15.17-17.19) | 15.8% (14.84-16.95) | -1.9% (-3.53--0.17) | 14.05% (1.09-31.55) |
| **1937** | 14.33% (13.49-15.28) | 14.25% (13.35-15.23) | -0.59% (-2.47-1.24) | 13.88% (-298.52-342.78) |
| **1938** | 14.79% (13.95-15.74) | 14.62% (13.78-15.62) | -1.18% (-2.9-0.77) | 20.16% (-29.07-113.61) |
| **1939** | 17.91% (17-18.91) | 17.51% (16.62-18.53) | -2.26% (-4--0.65) | 9.85% (2.71-17.89) |
| **1940** | 15.19% (14.37-16.06) | 15.1% (14.27-16.02) | -0.55% (-2.48-1.16) | 6.83% (-16.88-35.09) |
| **1941** | 12.62% (11.86-13.44) | 12.69% (11.94-13.55) | 0.61% (-1.2-2.45) | 6.41% (-13.35-30.07) |
| **1942** | 21.41% (20.25-22.63) | 20.76% (19.67-21.95) | -3.04% (-4.64--1.6) | 8.65% (4.48-13.47) |
| **1943** | 16.77% (15.87-17.79) | 16.52% (15.62-17.55) | -1.45% (-3.06-0.21) | 8.18% (-1.35-18.94) |
| **1944** | 16.13% (15.23-17.13) | 15.86% (14.96-16.8) | -1.68% (-3.35-0.02) | 12% (-0.16-27.2) |
| **1945** | 13.9% (13.01-14.82) | 13.9% (13.01-14.82) | 0% (0-0) | 0% (0-0) |
| **1946** | 17.52% (16.48-18.62) | 16.96% (15.9-18.06) | -3.2% (-4.69--1.43) | 15.57% (7.18-25.16) |
| **1947** | 16.75% (15.7-17.84) | 16.32% (15.27-17.37) | -2.57% (-4.31--0.86) | 15.09% (5.06-28.53) |
| **1948** | 18.42% (17.36-19.54) | 18.15% (17.04-19.26) | -1.49% (-3-0.02) | 6.26% (-0.11-12.64) |
| **1949** | 18.98% (17.9-19.96) | 18.52% (17.48-19.59) | -2.44% (-4.06--0.76) | 9.1% (2.59-15.51) |
| **1950** | 20.5% (19.31-21.6) | 20.05% (18.88-21.17) | -2.21% (-3.81--0.72) | 6.88% (2.22-11.82) |
| **1951** | 16.43% (15.38-17.36) | 16.52% (15.46-17.51) | 0.51% (-1.25-2.4) | -3.34% (-16.79-8.1) |

| | | | | |
|---|---|---|---|---|
| **1952** | 17.81% (16.75-18.81) | 17.74% (16.71-18.75) | -0.35% (-2.07-1.38) | 1.51% (-6.26-9.4) |
| **1953** | 18.38% (17.39-19.31) | 18.29% (17.33-19.25) | -0.47% (-2.18-1.29) | 2.16% (-5.85-8.95) |
| **1954** | 24.19% (22.99-25.39) | 23.66% (22.48-24.81) | -2.2% (-3.71--0.51) | 5.03% (1.22-8.65) |
| **1955** | 22.85% (21.8-23.89) | 22.41% (21.3-23.5) | -1.93% (-3.5--0.41) | 4.95% (1.06-9.14) |
| **1956** | 19.01% (17.97-19.94) | 18.98% (17.98-19.99) | -0.14% (-1.73-1.67) | 0.55% (-6.41-6.57) |
| **1957** | 14.58% (13.77-15.37) | 14.92% (14.11-15.75) | 2.32% (0.42-4.39) | -42.93% (-477.87-207.59) |
| **1958** | 18% (17.09-18.9) | 17.94% (17.01-18.89) | -0.35% (-2.4-1.63) | 1.57% (-7.48-10.38) |
| **1959** | 28.13% (26.98-29.35) | 27.48% (26.27-28.67) | -2.3% (-3.62--0.9) | 4.54% (1.75-7.19) |
| **1960** | 21.6% (20.52-22.74) | 21.34% (20.23-22.5) | -1.24% (-2.96-0.52) | 3.4% (-1.46-8.48) |
| **1961** | 19.76% (18.72-20.75) | 19.63% (18.67-20.64) | -0.62% (-2.38-0.96) | 2.01% (-3.35-7.89) |
| **1962** | 22.15% (21.09-23.26) | 22.07% (20.96-23.12) | -0.38% (-2.12-1.33) | 0.98% (-3.64-5.59) |
| **1963** | 17.24% (16.39-18.05) | 17.7% (16.87-18.53) | 2.67% (0.61-4.8) | -13.65% (-27.16--3.5) |
| **1964** | 16.87% (16.07-17.63) | 17.29% (16.48-18.13) | 2.49% (0.32-4.33) | -14.73% (-27.97--1.64) |
| **1965** | 21.12% (20.07-22.2) | 21.17% (20.06-22.25) | 0.24% (-1.78-2.22) | -0.69% (-6.37-5.15) |
| **1966** | 22.81% (21.38-24.28) | 22.9% (21.39-24.45) | 0.43% (-1.66-2.42) | -1.08% (-6.22-4.14) |

*Table S. 3 Predicted Probability of elevated depressive symptoms in natural course and counterfactual scenario, relative difference, and contribution by birth cohort and **stratified by sex**. All estimates are presented with 95% confidence intervals. Positive contributions imply that the counterfactual decreases depression risk and negative contributions imply that the counterfactual increases depression risk.*

| | | **Probability of elevated depressive symptoms (% (95%CI))** | | **% relative difference** | **% contribution** |
|---|---|---|---|---|---|
| **Birth Cohort** | sex | Natural Course Scenario | Counterfactual Scenario | | |
| **1916** | female | 21.18% (18.62-23.82) | 20.43% (17.92-23.08) | -3.52% (-4.92--2.19) | 12.53% (6.86-20.48) |
| **1917** | female | 23.37% (21.26-25.61) | 22.58% (20.5-24.83) | -3.34% (-4.55--2.14) | 9.62% (5.82-13.94) |
| **1918** | female | 24.94% (22.83-26.97) | 24.1% (22.06-26.05) | -3.36% (-4.65--2.15) | 8.55% (5.25-12.51) |
| **1919** | female | 19.9% (18.3-21.6) | 19.45% (17.84-21.11) | -2.31% (-3.73--0.92) | 9.64% (3.96-17.35) |
| **1920** | female | 20.6% (18.98-22.38) | 20.14% (18.56-21.97) | -2.2% (-3.58--0.91) | 8.28% (3.33-14.17) |
| **1921** | female | 20.19% (18.57-21.88) | 19.84% (18.26-21.57) | -1.75% (-3.18--0.37) | 6.9% (1.54-13.95) |
| **1922** | female | 20.8% (19.13-22.54) | 20.31% (18.7-22.04) | -2.39% (-3.71--1) | 8.81% (3.5-14.93) |
| **1923** | female | 16.49% (15.2-17.93) | 16.51% (15.16-17.97) | 0.12% (-1.34-1.55) | -1.95% (-57.63-28.87) |
| **1924** | female | 20.96% (19.51-22.52) | 20.56% (19.11-22.14) | -1.88% (-3.3--0.54) | 6.75% (1.93-12.7) |
| **1925** | female | 17.87% (16.49-19.19) | 17.5% (16.13-18.83) | -2.04% (-3.53--0.61) | 13.42% (4.19-29.53) |
| **1926** | female | 22.24% (20.61-23.8) | 21.75% (20.18-23.26) | -2.2% (-3.54--0.94) | 6.91% (2.87-11.28) |
| **1927** | female | 15.84% (14.52-17.06) | 15.71% (14.42-16.86) | -0.81% (-2.26-0.66) | 12.9% (-175.75-158.95) |
| **1928** | female | 21.45% (19.91-22.93) | 20.97% (19.44-22.41) | -2.25% (-3.55--1.01) | 7.64% (3.35-12.33) |

| | | | | | |
|---|---|---|---|---|---|
| **1929** | female | 22.54% (20.89-24.08) | 21.95% (20.36-23.5) | -2.6% (-3.87--1.31) | 7.91% (3.9-12.55) |
| **1930** | female | 21.48% (19.89-22.99) | 20.89% (19.3-22.39) | -2.75% (-3.92--1.61) | 9.3% (5.23-14) |
| **1931** | female | 18.72% (17.29-20.17) | 18.34% (16.93-19.74) | -2.03% (-3.45--0.75) | 10.51% (3.68-20.43) |
| **1932** | female | 18.95% (17.62-20.26) | 18.46% (17.13-19.82) | -2.59% (-4.04--1.2) | 12.91% (6.13-23.56) |
| **1933** | female | 15.89% (14.63-17.01) | 15.7% (14.44-16.86) | -1.21% (-2.9-0.43) | 19.03% (-142.52-153.02) |
| **1934** | female | 16.18% (14.85-17.39) | 15.97% (14.64-17.17) | -1.29% (-2.74-0.14) | 17.79% (-80.63-98.61) |
| **1935** | female | 15.88% (14.59-16.95) | 15.76% (14.48-16.9) | -0.75% (-2.25-0.63) | 12.17% (-119.12-224.51) |
| **1936** | female | 16.67% (15.4-17.89) | 16.42% (15.2-17.64) | -1.49% (-2.88--0.07) | 15.51% (-3.33-57.41) |
| **1937** | female | 15.64% (14.41-16.76) | 15.58% (14.4-16.72) | -0.42% (-2.06-1.09) | 8.05% (-193.27-268.44) |
| **1938** | female | 15.48% (14.35-16.65) | 15.39% (14.27-16.59) | -0.59% (-2.02-0.93) | 8.47% (-229.05-203.67) |
| **1939** | female | 19.61% (18.25-20.96) | 19.29% (17.96-20.64) | -1.61% (-2.97--0.2) | 7.14% (0.8-14.15) |
| **1940** | female | 16.72% (15.52-18.01) | 16.67% (15.46-17.93) | -0.24% (-1.67-1.15) | 3.03% (-17.45-27.08) |
| **1941** | female | 13.56% (12.51-14.64) | 13.65% (12.59-14.74) | 0.69% (-0.84-2.27) | 5.78% (-9.2-27.21) |
| **1942** | female | 21.83% (20.32-23.39) | 21.31% (19.84-22.8) | -2.39% (-3.68--1.12) | 7.76% (3.75-12.36) |
| **1943** | female | 17.48% (16.24-18.67) | 17.39% (16.12-18.58) | -0.51% (-1.9-0.94) | 3.69% (-6.79-16.69) |
| **1944** | female | 18.13% (16.88-19.38) | 18% (16.74-19.25) | -0.72% (-2.22-0.82) | 4.35% (-5.01-14.62) |
| **1945** | female | 15.13% (13.99-16.27) | 15.13% (13.99-16.27) | 0% (0-0) | 0% (0-0) |

| | | | | | |
|---|---|---|---|---|---|
| **1946** | female | 17.69% (16.4-18.93) | 17.38% (16.14-18.76) | -1.73% (-3.41--0.31) | 11.94% (2.11-28.13) |
| **1947** | female | 19.92% (18.61-21.29) | 19.5% (18.2-20.81) | -2.1% (-3.59--0.63) | 8.8% (2.81-15.73) |
| **1948** | female | 19.3% (18.08-20.49) | 19.07% (17.85-20.34) | -1.19% (-2.7-0.23) | 5.42% (-1.02-12.94) |
| **1949** | female | 19.22% (17.98-20.41) | 18.97% (17.73-20.18) | -1.33% (-2.81-0.04) | 6.25% (-0.17-13.22) |
| **1950** | female | 19.61% (18.37-20.88) | 19.43% (18.15-20.72) | -0.94% (-2.28-0.43) | 4.04% (-2.12-10.48) |
| **1951** | female | 17.61% (16.43-18.86) | 17.71% (16.5-18.88) | 0.58% (-0.9-2.12) | -3.93% (-18.51-6.49) |
| **1952** | female | 18.71% (17.51-20) | 18.72% (17.42-19.97) | 0.08% (-1.44-1.6) | -0.59% (-8.47-7.79) |
| **1953** | female | 20.09% (18.81-21.42) | 20.04% (18.74-21.3) | -0.27% (-1.76-1.31) | 1.24% (-5.29-7.1) |
| **1954** | female | 26.51% (24.91-28.1) | 26.08% (24.5-27.58) | -1.59% (-2.77--0.25) | 3.69% (0.58-6.51) |
| **1955** | female | 24.26% (22.72-25.77) | 24% (22.5-25.42) | -1.11% (-2.46-0.19) | 2.98% (-0.51-6.51) |
| **1956** | female | 21.7% (20.31-23.15) | 21.79% (20.34-23.23) | 0.4% (-1.09-1.92) | -1.42% (-6.32-3.54) |
| **1957** | female | 16.6% (15.49-17.76) | 17.11% (15.94-18.33) | 3.1% (1.47-4.91) | -34.67% (-150.6--13.2) |
| **1958** | female | 21.71% (20.31-23.14) | 21.73% (20.36-23.22) | 0.12% (-1.31-1.63) | -0.24% (-5.54-4.52) |
| **1959** | female | 29.78% (28.08-31.61) | 29.24% (27.58-31.12) | -1.81% (-3.06--0.65) | 3.67% (1.29-6.2) |
| **1960** | female | 23.36% (21.74-25.09) | 23.22% (21.65-24.87) | -0.57% (-2.08-0.96) | 1.72% (-2.73-5.8) |
| **1961** | female | 21.64% (20.3-23.13) | 21.7% (20.35-23.16) | 0.29% (-1.2-1.93) | -0.93% (-6.77-4.07) |
| **1962** | female | 24.31% (22.91-25.81) | 24.41% (22.98-25.84) | 0.39% (-1.01-1.65) | -1.04% (-4.67-2.7) |

| | | | | | |
|---|---|---|---|---|---|
| **1963** | female | 20.84% (19.69-22.06) | 21.42% (20.24-22.61) | 2.81% (1.19-4.52) | -10.28% (-17.56--4.01) |
| **1964** | female | 19.65% (18.56-20.8) | 20.41% (19.2-21.62) | 3.85% (2.1-5.63) | -16.61% (-27.84--8.72) |
| **1965** | female | 24.03% (22.52-25.69) | 24.38% (22.8-26) | 1.43% (-0.33-3.13) | -3.86% (-8.59-0.86) |
| **1966** | female | 25.6% (23.55-27.78) | 26.29% (24.18-28.6) | 2.68% (0.93-4.34) | -6.69% (-11.14--2.09) |
| **1916** | male | 21.83% (18.88-25.01) | 21.31% (18.41-24.45) | -2.37% (-4.19--0.19) | 6.41% (0.57-11.78) |
| **1917** | male | 22.77% (20.14-25.65) | 22.05% (19.63-24.82) | -3.13% (-4.79--1.36) | 7.7% (3.23-12.5) |
| **1918** | male | 24.25% (21.69-26.82) | 23.45% (21.17-25.89) | -3.28% (-4.9--1.65) | 7.34% (3.68-11.21) |
| **1919** | male | 19.06% (17.13-21.09) | 18.7% (16.79-20.68) | -1.91% (-3.75--0.17) | 6.68% (0.62-13.31) |
| **1920** | male | 19.57% (17.62-21.46) | 19.24% (17.36-21.21) | -1.66% (-3.29-0.18) | 5.44% (-0.57-10.82) |
| **1921** | male | 18.17% (16.35-20.15) | 17.82% (16.03-19.79) | -1.89% (-3.64-0.01) | 7.35% (-0.03-15.4) |
| **1922** | male | 18.3% (16.42-20.17) | 17.92% (16.11-19.79) | -2.05% (-3.82--0.12) | 7.83% (0.41-15.79) |
| **1923** | male | 14.38% (12.9-15.72) | 14.42% (12.97-15.9) | 0.33% (-1.66-2.22) | -2.23% (-109.45-104.49) |
| **1924** | male | 17.15% (15.53-18.9) | 16.85% (15.27-18.53) | -1.75% (-3.55--0.13) | 8.12% (0.68-18.4) |
| **1925** | male | 14.78% (13.36-16.27) | 14.68% (13.28-16.25) | -0.63% (-2.81-1.23) | 7.36% (-39.14-56.96) |
| **1926** | male | 18.11% (16.48-19.89) | 17.76% (16.17-19.54) | -1.89% (-3.59--0.15) | 7.5% (0.6-14.66) |
| **1927** | male | 13.54% (12.17-14.98) | 13.65% (12.28-15.09) | 0.77% (-1.23-2.93) | -1.3% (-478.01-213.58) |
| **1928** | male | 18.06% (16.57-19.78) | 17.67% (16.19-19.35) | -2.18% (-3.84--0.52) | 8.72% (2-15.63) |

| | | | | | |
|---|---|---|---|---|---|
| **1929** | male | 19.46% (17.71-21.37) | 18.84% (17.22-20.8) | -3.16% (-4.69--1.46) | 10.26% (4.71-16.37) |
| **1930** | male | 19.41% (17.69-21.37) | 18.9% (17.22-20.72) | -2.63% (-4.42--0.88) | 8.68% (2.92-14.94) |
| **1931** | male | 17.65% (16.09-19.29) | 17.25% (15.76-18.92) | -2.27% (-4.03--0.46) | 9.57% (1.92-18.11) |
| **1932** | male | 17.51% (16-19.16) | 17.09% (15.55-18.81) | -2.4% (-4.18--0.64) | 10.35% (2.73-19.85) |
| **1933** | male | 15.38% (14-16.88) | 15.18% (13.8-16.62) | -1.25% (-3.14-0.7) | 9.87% (-7.83-37.79) |
| **1934** | male | 16.21% (14.62-17.75) | 16.11% (14.54-17.56) | -0.61% (-2.55-1.31) | 3.65% (-7.79-18.18) |
| **1935** | male | 15.82% (14.36-17.29) | 15.74% (14.27-17.17) | -0.48% (-2.22-1.45) | 3.2% (-9.63-19.05) |
| **1936** | male | 16.35% (14.87-17.87) | 16.15% (14.73-17.63) | -1.25% (-3-0.57) | 6.99% (-3.33-19.55) |
| **1937** | male | 15.05% (13.7-16.55) | 14.97% (13.67-16.38) | -0.51% (-2.47-1.4) | 4.8% (-19.03-38.84) |
| **1938** | male | 15.06% (13.68-16.44) | 14.97% (13.59-16.42) | -0.6% (-2.38-1.27) | 5.57% (-16.33-41.41) |
| **1939** | male | 18.28% (16.81-19.9) | 17.98% (16.53-19.51) | -1.66% (-3.19-0) | 6.21% (0.01-12.86) |
| **1940** | male | 15.85% (14.38-17.4) | 15.82% (14.39-17.34) | -0.19% (-1.99-1.56) | 0.85% (-11.65-14.04) |
| **1941** | male | 12.79% (11.6-14.01) | 13% (11.77-14.29) | 1.68% (-0.36-3.68) | 22% (-413.97-266.81) |
| **1942** | male | 19.59% (17.95-21.31) | 19.07% (17.44-20.89) | -2.64% (-4.37--1) | 8.41% (3.07-14.61) |
| **1943** | male | 15.78% (14.37-17.37) | 15.65% (14.24-17.17) | -0.78% (-2.68-1.11) | 5.04% (-7.74-22.2) |
| **1944** | male | 15.61% (14.28-17.04) | 15.29% (13.99-16.78) | -2.05% (-3.79--0.28) | 14.84% (2.15-37.45) |
| **1945** | male | 13.41% (12.22-14.81) | 13.41% (12.22-14.81) | 0% (0-0) | 0% (0-0) |

| | | | | | |
|---|---|---|---|---|---|
| **1946** | male | 15.38% (13.96-16.93) | 15.03% (13.61-16.53) | -2.3% (-4.08--0.42) | 18.48% (3.22-58.47) |
| **1947** | male | 17.67% (16.18-19.35) | 17.08% (15.56-18.78) | -3.33% (-5.04--1.68) | 13.94% (6.9-24.91) |
| **1948** | male | 17.94% (16.49-19.52) | 17.59% (16.14-19.22) | -1.95% (-3.67--0.26) | 7.71% (1.12-15.51) |
| **1949** | male | 19.16% (17.6-20.78) | 18.83% (17.29-20.49) | -1.72% (-3.21-0.06) | 5.95% (-0.2-11.8) |
| **1950** | male | 20.43% (18.67-22.01) | 20.19% (18.45-21.91) | -1.17% (-2.91-0.55) | 3.56% (-1.59-8.65) |
| **1951** | male | 19.03% (17.27-20.61) | 19.09% (17.38-20.81) | 0.3% (-1.47-2.04) | -0.79% (-7.33-4.96) |
| **1952** | male | 19.48% (17.83-21.08) | 19.43% (17.81-21.1) | -0.26% (-2.1-1.49) | 1% (-4.9-6.59) |
| **1953** | male | 20.11% (18.59-21.72) | 19.96% (18.47-21.55) | -0.72% (-2.5-1.05) | 2.16% (-3.13-7.64) |
| **1954** | male | 25.49% (23.6-27.43) | 24.8% (22.97-26.67) | -2.7% (-4.36--1.04) | 5.65% (2.26-9.07) |
| **1955** | male | 22.43% (20.69-24.12) | 21.95% (20.23-23.65) | -2.11% (-3.75--0.52) | 5.25% (1.29-9.35) |
| **1956** | male | 19.63% (18-21.21) | 19.49% (17.82-21.11) | -0.72% (-2.38-0.92) | 2.28% (-2.97-8.02) |
| **1957** | male | 14.86% (13.56-16.22) | 15.11% (13.8-16.46) | 1.67% (-0.31-3.56) | -17.45% (-101.33-6.54) |
| **1958** | male | 18.49% (16.99-20.04) | 18.28% (16.75-19.87) | -1.16% (-3.04-0.71) | 4.24% (-2.6-11.32) |
| **1959** | male | 24.64% (22.76-26.6) | 23.96% (22.15-25.85) | -2.77% (-4.56--0.98) | 6.03% (2.25-10.12) |
| **1960** | male | 20.33% (18.66-21.99) | 20.1% (18.46-21.91) | -1.12% (-3.05-0.73) | 3.25% (-2.22-9.06) |
| **1961** | male | 18.29% (16.89-19.73) | 18.26% (16.79-19.77) | -0.14% (-2.22-1.86) | 0.24% (-7.29-8.52) |
| **1962** | male | 20.55% (19.15-22.06) | 20.44% (18.95-21.95) | -0.51% (-2.4-1.22) | 1.42% (-3.65-6.81) |

| | | | | | |
|---|---|---|---|---|---|
| **1963** | male | 17.27% (16.08-18.57) | 17.39% (16.18-18.66) | 0.72% (-1.18-2.52) | -3.23% (-12.01-5.21) |
| **1964** | male | 16.35% (15.08-17.55) | 16.54% (15.25-17.78) | 1.12% (-0.78-3.25) | -6.09% (-20.3-4.77) |
| **1965** | male | 18.88% (17.06-20.53) | 18.66% (16.78-20.26) | -1.17% (-3.31-0.91) | 4.01% (-3.08-11.29) |
| **1966** | male | 19.92% (17.62-21.89) | 19.63% (17.33-21.68) | -1.49% (-3.55-0.77) | 4.71% (-2.1-11.31) |

*Table S. 4 Predicted Probability of elevated depressive symptoms in natural course and counterfactual scenario, relative difference, and contribution by birth cohort and **stratified by race/ethnicity**. All estimates are presented with 95% confidence intervals. Positive contributions imply that the counterfactual decreases depression risk and negative contributions imply that the counterfactual increases depression risk .*

| | | **Probability of elevated depressive symptoms (% (95%CI))** | | **% relative difference** | **% contribution** |
|---|---|---|---|---|---|
| **Birth Cohort** | race/ethnicity | Natural Course Scenario | Counterfactual Scenario | | |
| **1916** | Black | 36.03% (32.29-39.95) | 33.46% (30.07-37.07) | -7.11% (-10.42--4.12) | 16.29% (9.61-23.91) |
| **1917** | Black | 32.9% (30.06-35.78) | 30.95% (28.3-33.78) | -5.93% (-8.5--3.72) | 15.43% (9.74-22.73) |
| **1918** | Black | 34.7% (32.16-37.47) | 33.46% (31.06-36.1) | -3.55% (-5.26--2.1) | 8.48% (5.33-12.92) |
| **1919** | Black | 26.15% (24.16-28.24) | 25.69% (23.75-27.69) | -1.75% (-3.11--0.53) | 7.73% (2.47-14.82) |
| **1920** | Black | 28.63% (26.57-30.66) | 28.41% (26.37-30.47) | -0.76% (-1.67-0.13) | 2.54% (-0.44-5.85) |
| **1921** | Black | 25.94% (23.95-27.97) | 25.77% (23.85-27.7) | -0.65% (-1.55-0.19) | 3.07% (-0.8-7.55) |
| **1922** | Black | 28% (26.1-30.01) | 27.86% (25.9-29.84) | -0.5% (-1.34-0.29) | 1.83% (-1.12-5.1) |
| **1923** | Black | 23.02% (21.3-24.76) | 23.03% (21.31-24.8) | 0.06% (-0.77-0.91) | -0.26% (-9.54-8.39) |

| | | | | | |
|---|---|---|---|---|---|
| **1924** | Black | 24.72% (23.05-26.3) | 24.65% (22.92-26.31) | -0.3% (-1.16-0.56) | 1.71% (-3.23-7.14) |
| **1925** | Black | 21.25% (19.78-22.78) | 21.2% (19.68-22.72) | -0.24% (-1.09-0.61) | 4.64% (-68.92-77.98) |
| **1926** | Black | 23.68% (22.12-25.29) | 23.52% (21.94-25.12) | -0.64% (-1.51-0.26) | 4.48% (-1.84-11.81) |
| **1927** | Black | 19.1% (17.77-20.45) | 19.13% (17.75-20.48) | 0.17% (-0.7-1.09) | 2.4% (-39.43-71.75) |
| **1928** | Black | 24.15% (22.58-25.77) | 24.06% (22.49-25.67) | -0.4% (-1.21-0.51) | 2.46% (-2.99-7.99) |
| **1929** | Black | 26.38% (24.66-28.22) | 26.16% (24.44-27.99) | -0.82% (-1.64-0.02) | 3.57% (-0.08-7.22) |
| **1930** | Black | 24.3% (22.72-25.99) | 24.2% (22.63-25.88) | -0.44% (-1.23-0.36) | 2.69% (-2.07-8.44) |
| **1931** | Black | 21.47% (19.95-23.09) | 21.41% (19.89-23.05) | -0.28% (-1.21-0.6) | 4.6% (-43.51-57.73) |
| **1932** | Black | 22.04% (20.65-23.55) | 21.97% (20.52-23.51) | -0.32% (-1.11-0.45) | 4.24% (-6.67-26.69) |
| **1933** | Black | 21.05% (19.61-22.59) | 21.02% (19.58-22.53) | -0.15% (-1.15-0.64) | 3.13% (-83.7-77.24) |
| **1934** | Black | 19.81% (18.47-21.28) | 19.79% (18.52-21.25) | -0.08% (-1.03-0.78) | -0.56% (-113.59-98.41) |
| **1935** | Black | 20% (18.61-21.47) | 19.99% (18.63-21.49) | -0.02% (-0.92-0.88) | -0.28% (-139.9-141.66) |
| **1936** | Black | 21.2% (19.73-22.76) | 21.18% (19.72-22.67) | -0.08% (-0.99-0.77) | 1.18% (-56.88-82.8) |
| **1937** | Black | 19.5% (18.17-20.96) | 19.47% (18.17-20.9) | -0.18% (-1.24-0.71) | -2.82% (-104.53-90.03) |
| **1938** | Black | 18.23% (16.94-19.62) | 18.21% (16.95-19.57) | -0.13% (-1.02-0.82) | -0.98% (-16.05-9.7) |
| **1939** | Black | 20.01% (18.68-21.36) | 19.93% (18.63-21.37) | -0.4% (-1.42-0.56) | -5.13% (-204.48-139.11) |
| **1940** | Black | 18.15% (16.79-19.49) | 18.12% (16.76-19.42) | -0.19% (-1.12-0.81) | -1.44% (-14.27-7.56) |

| | | | | | |
|---|---|---|---|---|---|
| **1941** | Black | 14.53% (13.34-15.72) | 14.61% (13.44-15.81) | 0.57% (-0.4-1.49) | 1.4% (-1.03-4.1) |
| **1942** | Black | 24.82% (23.17-26.56) | 24.67% (23.04-26.33) | -0.61% (-1.42-0.16) | 3.31% (-0.94-8.23) |
| **1943** | Black | 21.01% (19.49-22.6) | 20.92% (19.48-22.51) | -0.46% (-1.3-0.39) | 8.13% (-111.74-146.75) |
| **1944** | Black | 23.81% (22.21-25.49) | 23.61% (22.01-25.27) | -0.84% (-1.74-0.08) | 5.64% (-0.52-12.99) |
| **1945** | Black | 20.28% (18.9-21.89) | 20.28% (18.9-21.89) | 0% (0-0) | 0% (0-0) |
| **1946** | Black | 26.33% (24.7-28.09) | 26.09% (24.44-27.88) | -0.9% (-1.84--0.07) | 3.95% (0.31-8.16) |
| **1947** | Black | 24.56% (22.99-26.3) | 24.31% (22.78-26.01) | -1.03% (-2.02--0.02) | 5.96% (0.15-12.43) |
| **1948** | Black | 27.74% (26.24-29.4) | 27.5% (25.97-29.19) | -0.86% (-1.66--0.11) | 3.2% (0.39-6.31) |
| **1949** | Black | 26.91% (25.34-28.54) | 26.64% (25.12-28.34) | -0.98% (-1.9--0.12) | 3.95% (0.49-8.07) |
| **1950** | Black | 27.17% (25.52-28.92) | 26.91% (25.28-28.65) | -0.97% (-1.83--0.12) | 3.74% (0.47-7.6) |
| **1951** | Black | 22.4% (20.91-23.9) | 22.27% (20.81-23.82) | -0.59% (-1.49-0.32) | 6.19% (-4.11-23.23) |
| **1952** | Black | 24.54% (22.91-26.23) | 24.3% (22.65-25.99) | -0.98% (-2.01--0.03) | 5.63% (0.14-13.51) |
| **1953** | Black | 24.66% (23.08-26.4) | 24.43% (22.83-26.05) | -0.93% (-1.85-0.09) | 5.26% (-0.52-11.57) |
| **1954** | Black | 32.73% (30.93-34.69) | 32.2% (30.37-34.12) | -1.6% (-2.51--0.69) | 4.23% (1.84-6.63) |
| **1955** | Black | 31.63% (29.9-33.44) | 31.18% (29.45-32.98) | -1.44% (-2.33--0.6) | 4.06% (1.69-6.67) |
| **1956** | Black | 25.84% (24.3-27.52) | 25.52% (24.05-27.21) | -1.24% (-2.3--0.27) | 5.86% (1.24-10.93) |
| **1957** | Black | 21.22% (19.88-22.7) | 21.21% (19.84-22.69) | -0.05% (-1.14-0.93) | 0.54% (-135.02-67.16) |

| | | | | | |
|---|---|---|---|---|---|
| **1958** | Black | 24.85% (23.24-26.48) | 24.56% (23.03-26.16) | -1.18% (-2.26--0.09) | 6.26% (0.48-13.47) |
| **1959** | Black | 33.17% (31.35-34.99) | 32.64% (30.79-34.47) | -1.6% (-2.51--0.77) | 4.13% (2-6.44) |
| **1960** | Black | 24.79% (23.2-26.38) | 24.5% (22.87-26.18) | -1.15% (-2.12--0.22) | 6.36% (1.18-12.41) |
| **1961** | Black | 22.1% (20.64-23.54) | 21.88% (20.4-23.28) | -0.97% (-2.05-0.01) | 12.25% (-0.2-65.15) |
| **1962** | Black | 24.01% (22.59-25.53) | 23.64% (22.14-25.17) | -1.51% (-2.56--0.43) | 9.8% (2.86-20.21) |
| **1963** | Black | 19.45% (18.4-20.67) | 19.28% (18.18-20.56) | -0.83% (-2.03-0.3) | -15.44% (-177.5-169.39) |
| **1964** | Black | 18.09% (17.12-19.25) | 18% (17.04-19.19) | -0.45% (-1.51-0.73) | -3.83% (-18.24-6.32) |
| **1965** | Black | 22.32% (20.95-23.83) | 22.07% (20.67-23.55) | -1.15% (-2.22--0.08) | 12.56% (0.69-38.93) |
| **1966** | Black | 25.48% (23.56-27.51) | 25.11% (23.24-27.18) | -1.45% (-2.54--0.36) | 7.09% (1.93-13.96) |
| **1916** | Hispanic | 11.59% (9.71-13.68) | 11.61% (9.68-13.76) | 0.21% (-2.19-2.62) | 0.29% (-2.86-3.46) |
| **1917** | Hispanic | 14.51% (12.67-16.41) | 14.5% (12.66-16.45) | -0.02% (-2.07-1.93) | 0.07% (-5.21-4.64) |
| **1918** | Hispanic | 22.37% (20.26-24.53) | 22.14% (20.04-24.3) | -1.02% (-2.02--0.1) | 13.16% (-195.78-134.69) |
| **1919** | Hispanic | 21.88% (20.14-23.75) | 21.66% (19.92-23.42) | -1.01% (-1.69--0.36) | 16.36% (-155.27-364.52) |
| **1920** | Hispanic | 29.31% (27.25-31.28) | 28.89% (26.87-30.78) | -1.43% (-2.04--0.8) | 5.07% (2.88-7.98) |
| **1921** | Hispanic | 30.45% (28.4-32.43) | 30.06% (28.03-32) | -1.28% (-1.85--0.7) | 4.17% (2.25-6.46) |
| **1922** | Hispanic | 33.68% (31.63-35.76) | 33.21% (31.17-35.25) | -1.4% (-2.07--0.77) | 3.75% (2.03-5.59) |
| **1923** | Hispanic | 26.25% (24.48-28) | 26% (24.26-27.69) | -0.96% (-1.6--0.34) | 4.89% (1.66-9) |

| | | | | | |
|---|---|---|---|---|---|
| **1924** | Hispanic | 25.87% (24.15-27.5) | 25.63% (23.91-27.25) | -0.9% (-1.49--0.33) | 4.84% (1.83-9.34) |
| **1925** | Hispanic | 21.89% (20.36-23.35) | 21.71% (20.17-23.15) | -0.84% (-1.45--0.28) | 16.78% (-110.76-275.34) |
| **1926** | Hispanic | 22.4% (20.81-23.94) | 22.27% (20.71-23.79) | -0.57% (-1.1--0.06) | 8.94% (-66.78-62.72) |
| **1927** | Hispanic | 17.75% (16.41-18.99) | 17.65% (16.32-18.9) | -0.56% (-1.18--0.04) | -2.91% (-7.64--0.22) |
| **1928** | Hispanic | 21.41% (19.93-22.86) | 21.25% (19.73-22.69) | -0.72% (-1.27--0.24) | 16.35% (-200.39-282.05) |
| **1929** | Hispanic | 25.16% (23.42-26.86) | 24.86% (23.11-26.57) | -1.21% (-1.79--0.66) | 7.52% (3.69-13.95) |
| **1930** | Hispanic | 24.21% (22.44-25.91) | 23.96% (22.19-25.66) | -1.07% (-1.64--0.49) | 8.27% (3.65-18.79) |
| **1931** | Hispanic | 20.62% (18.91-22.23) | 20.46% (18.78-22.04) | -0.82% (-1.47--0.25) | -15.76% (-264.81-400.61) |
| **1932** | Hispanic | 22.03% (20.29-23.56) | 21.82% (20.06-23.37) | -0.93% (-1.54--0.41) | 17.8% (-174.11-168.07) |
| **1933** | Hispanic | 20.76% (19.1-22.16) | 20.64% (18.95-22.04) | -0.56% (-1.09--0.05) | -9.36% (-241.75-289.03) |
| **1934** | Hispanic | 23.94% (22.18-25.48) | 23.79% (22.01-25.35) | -0.63% (-1.17--0.16) | 5.26% (1.28-15.97) |
| **1935** | Hispanic | 22.82% (21.17-24.29) | 22.74% (21.08-24.22) | -0.34% (-0.84-0.16) | 4.41% (-3.35-24.07) |
| **1936** | Hispanic | 22.44% (20.88-23.92) | 22.34% (20.79-23.77) | -0.45% (-0.98-0.02) | 7.04% (-26.32-47.21) |
| **1937** | Hispanic | 21.95% (20.39-23.46) | 21.89% (20.32-23.37) | -0.28% (-0.78-0.23) | 4.87% (-96-67.69) |
| **1938** | Hispanic | 22.05% (20.52-23.59) | 21.98% (20.36-23.5) | -0.34% (-0.83-0.13) | 5.87% (-46.43-75.35) |
| **1939** | Hispanic | 22.35% (20.81-23.87) | 22.25% (20.69-23.77) | -0.44% (-0.96-0.02) | 7.19% (-48.62-56.44) |
| **1940** | Hispanic | 21.12% (19.66-22.58) | 21.04% (19.61-22.51) | -0.37% (-0.93-0.21) | -1.02% (-205.01-182.25) |

| | | | | | |
|---|---|---|---|---|---|
| **1941** | Hispanic | 17.13% (15.81-18.4) | 17.12% (15.83-18.39) | -0.04% (-0.58-0.53) | -0.16% (-2.74-2.32) |
| **1942** | Hispanic | 26.61% (24.96-28.35) | 26.38% (24.75-28.11) | -0.87% (-1.37--0.42) | 4.19% (2.02-7.57) |
| **1943** | Hispanic | 24.16% (22.56-25.78) | 24.01% (22.42-25.66) | -0.59% (-1.11--0.12) | 4.67% (1.02-11.27) |
| **1944** | Hispanic | 24.37% (22.73-25.95) | 24.24% (22.64-25.83) | -0.53% (-1.01--0.08) | 3.95% (0.54-10.13) |
| **1945** | Hispanic | 21.1% (19.57-22.5) | 21.1% (19.57-22.5) | 0% (0-0) | 0% (0-0) |
| **1946** | Hispanic | 24.16% (22.43-25.72) | 23.98% (22.27-25.51) | -0.77% (-1.28--0.27) | 5.9% (2.25-14.73) |
| **1947** | Hispanic | 21.5% (19.85-22.95) | 21.49% (19.9-22.93) | -0.05% (-0.54-0.42) | 1.17% (-59.34-65.38) |
| **1948** | Hispanic | 24.04% (22.36-25.47) | 23.93% (22.28-25.34) | -0.42% (-0.88--0.02) | 3.37% (0.12-10.43) |
| **1949** | Hispanic | 23.14% (21.56-24.57) | 22.99% (21.41-24.44) | -0.64% (-1.11--0.2) | 7% (1.15-23.65) |
| **1950** | Hispanic | 23% (21.49-24.53) | 22.82% (21.31-24.33) | -0.78% (-1.31--0.34) | 9.26% (2.99-34.53) |
| **1951** | Hispanic | 19.63% (18.27-20.95) | 19.61% (18.25-20.92) | -0.09% (-0.66-0.42) | -1.31% (-19.21-9.47) |
| **1952** | Hispanic | 19.85% (18.52-21.16) | 19.8% (18.46-21.06) | -0.27% (-0.84-0.21) | -3.87% (-39.43-9.29) |
| **1953** | Hispanic | 20.46% (19.1-21.72) | 20.42% (19.05-21.65) | -0.2% (-0.73-0.31) | -3.23% (-134.2-91.94) |
| **1954** | Hispanic | 25.29% (23.7-26.8) | 25.17% (23.61-26.69) | -0.46% (-1.02-0.03) | 2.77% (-0.19-6.56) |
| **1955** | Hispanic | 25.75% (24.19-27.25) | 25.6% (24.04-27.09) | -0.58% (-1.08--0.11) | 3.25% (0.58-6.3) |
| **1956** | Hispanic | 23.53% (22.04-24.88) | 23.48% (22.01-24.85) | -0.2% (-0.69-0.31) | 2.09% (-3.22-9.25) |
| **1957** | Hispanic | 19.61% (18.33-20.8) | 19.63% (18.32-20.79) | 0.06% (-0.44-0.56) | 0.86% (-10.89-15.64) |

| | | | | | |
|---|---|---|---|---|---|
| **1958** | Hispanic | 21.62% (20.25-22.98) | 21.6% (20.26-22.98) | -0.06% (-0.61-0.48) | 1.36% (-84.68-74.71) |
| **1959** | Hispanic | 31.1% (29.43-32.89) | 30.87% (29.21-32.6) | -0.73% (-1.25--0.32) | 2.24% (1.01-4.08) |
| **1960** | Hispanic | 24.42% (22.9-26.04) | 24.32% (22.82-25.98) | -0.42% (-0.92-0.08) | 3.05% (-0.64-8.01) |
| **1961** | Hispanic | 22.5% (21.1-24) | 22.4% (20.95-23.86) | -0.46% (-0.98-0) | 7.25% (-5.29-60.83) |
| **1962** | Hispanic | 25.48% (23.96-26.93) | 25.34% (23.75-26.84) | -0.52% (-1.02--0.03) | 3.04% (0.2-6.78) |
| **1963** | Hispanic | 20.98% (19.73-22.18) | 21% (19.76-22.17) | 0.09% (-0.41-0.62) | 1.6% (-115.57-192.7) |
| **1964** | Hispanic | 19.69% (18.55-20.8) | 19.69% (18.53-20.82) | -0.01% (-0.57-0.56) | -0.15% (-15.84-13.78) |
| **1965** | Hispanic | 22.31% (20.93-23.76) | 22.2% (20.82-23.68) | -0.48% (-1.01--0.01) | 8.1% (-12.13-80.48) |
| **1966** | Hispanic | 25.14% (23.3-27.12) | 24.96% (23.18-26.9) | -0.73% (-1.29--0.26) | 4.58% (1.4-9.35) |
| **1916** | White | 16.65% (14.57-18.94) | 15.83% (13.91-17.97) | -4.92% (-7.34--2.63) | 15.5% (7.9-25.11) |
| **1917** | White | 16.09% (14.42-17.94) | 15.33% (13.66-17.11) | -4.71% (-7.07--2.54) | 16.11% (8.55-27.43) |
| **1918** | White | 18.22% (16.48-19.92) | 17.17% (15.51-18.93) | -5.79% (-7.77--3.72) | 15.24% (9.7-21.79) |
| **1919** | White | 13.05% (11.83-14.38) | 12.78% (11.55-14.06) | -2.07% (-4.2-0.28) | 16.27% (-2.09-58.22) |
| **1920** | White | 15.46% (14.05-16.9) | 14.86% (13.56-16.25) | -3.84% (-5.91--1.92) | 14.48% (6.71-24.65) |
| **1921** | White | 14.12% (12.78-15.48) | 13.61% (12.32-14.91) | -3.6% (-6.07--1.54) | 18.29% (6.98-38.1) |
| **1922** | White | 16.2% (14.76-17.62) | 15.37% (13.96-16.74) | -5.13% (-7.19--2.92) | 17.27% (9.01-26.94) |
| **1923** | White | 12.65% (11.5-13.71) | 12.43% (11.29-13.58) | -1.74% (-3.99-0.79) | 17.11% (-12.59-78.22) |

| | | | | | |
|---|---|---|---|---|---|
| **1924** | White | 15.1% (13.86-16.3) | 14.56% (13.32-15.82) | -3.58% (-5.58--1.49) | 14.55% (5.84-25.62) |
| **1925** | White | 13.06% (11.92-14.16) | 12.75% (11.62-13.83) | -2.43% (-4.64-0.02) | 18.31% (-0.77-53.41) |
| **1926** | White | 15.57% (14.32-16.76) | 15.03% (13.74-16.27) | -3.44% (-5.53--1.52) | 12.77% (5.42-21.9) |
| **1927** | White | 12.21% (11.23-13.21) | 12.14% (11.11-13.15) | -0.5% (-2.92-1.86) | 5.58% (-89.21-89.79) |
| **1928** | White | 16.48% (15.21-17.82) | 15.87% (14.61-17.16) | -3.69% (-5.58--1.6) | 11.94% (5.11-18.57) |
| **1929** | White | 17.93% (16.57-19.42) | 17.01% (15.59-18.4) | -5.09% (-7.07--3.26) | 13.84% (8.74-19.92) |
| **1930** | White | 16.29% (14.94-17.71) | 15.64% (14.35-16.95) | -3.96% (-5.98--1.9) | 12.88% (6.33-20.63) |
| **1931** | White | 13.95% (12.78-15.24) | 13.57% (12.39-14.76) | -2.77% (-4.91--0.67) | 14.91% (3.44-30.03) |
| **1932** | White | 14.47% (13.3-15.76) | 13.88% (12.78-15.15) | -4.05% (-6.25--1.81) | 18.86% (8.56-33.38) |
| **1933** | White | 12.28% (11.24-13.41) | 12.03% (10.98-13.12) | -2.04% (-4.2-0.15) | 25% (-98.98-196.41) |
| **1934** | White | 12.4% (11.38-13.48) | 12.17% (11.15-13.22) | -1.89% (-4.13-0.4) | 21.99% (-9.34-162.81) |
| **1935** | White | 12.43% (11.41-13.5) | 12.19% (11.15-13.24) | -1.92% (-4.05-0.4) | 22.23% (-8.95-157.6) |
| **1936** | White | 13.12% (12.08-14.18) | 12.71% (11.62-13.79) | -3.14% (-5.42--0.96) | 22.71% (7.53-64.85) |
| **1937** | White | 12.56% (11.51-13.53) | 12.23% (11.18-13.27) | -2.61% (-4.77--0.24) | 26.91% (1.1-149.81) |
| **1938** | White | 12.4% (11.41-13.44) | 12.18% (11.18-13.19) | -1.77% (-4.08-0.37) | 20.3% (-9.91-140.95) |
| **1939** | White | 15.4% (14.25-16.61) | 14.81% (13.64-16) | -3.83% (-5.9--1.76) | 14.35% (6.69-24.12) |
| **1940** | White | 13.51% (12.47-14.62) | 13.17% (12.1-14.27) | -2.54% (-4.81--0.27) | 15.53% (1.53-37.09) |

| | | | | | |
|---|---|---|---|---|---|
| **1941** | White | 10.52% (9.67-11.49) | 10.67% (9.8-11.61) | 1.43% (-1.06-4.04) | 16.45% (-80.82-128.33) |
| **1942** | White | 18.74% (17.28-20.08) | 17.83% (16.54-19.21) | -4.85% (-6.84--2.85) | 12.41% (7.51-17.25) |
| **1943** | White | 14.41% (13.32-15.6) | 14.01% (12.94-15.16) | -2.76% (-4.9--0.55) | 12.87% (2.71-26.43) |
| **1944** | White | 14.79% (13.68-16) | 14.19% (13.11-15.35) | -4.04% (-6.19--1.88) | 17.26% (8.32-28.63) |
| **1945** | White | 11.34% (10.45-12.32) | 11.34% (10.45-12.32) | 0% (0-0) | 0% (0-0) |
| **1946** | White | 13.81% (12.71-14.96) | 13.3% (12.29-14.48) | -3.63% (-5.7--1.49) | 20.24% (8.66-39.63) |
| **1947** | White | 13.62% (12.55-14.77) | 13.13% (12.11-14.24) | -3.57% (-5.98--1.2) | 21.16% (7.72-44.81) |
| **1948** | White | 15.64% (14.5-16.94) | 15.1% (14.02-16.29) | -3.43% (-5.32--1.36) | 12.59% (5.08-20.59) |
| **1949** | White | 15.42% (14.28-16.69) | 14.78% (13.67-16.02) | -4.12% (-6.43--2.02) | 15.6% (7.51-26.09) |
| **1950** | White | 16.58% (15.32-17.88) | 15.86% (14.57-17.11) | -4.37% (-6.22--2.38) | 13.75% (7.33-21.2) |
| **1951** | White | 13.53% (12.4-14.63) | 13.4% (12.29-14.58) | -0.98% (-3.29-1.5) | 6.12% (-9.14-22.85) |
| **1952** | White | 14.8% (13.65-15.96) | 14.48% (13.32-15.67) | -2.15% (-4.25-0.16) | 9.06% (-0.76-19.91) |
| **1953** | White | 14.82% (13.7-15.87) | 14.6% (13.43-15.72) | -1.49% (-3.56-0.69) | 6.43% (-3-16.08) |
| **1954** | White | 20.63% (19.18-22.05) | 19.73% (18.29-21.17) | -4.35% (-6.24--2.34) | 9.7% (5.41-14.23) |
| **1955** | White | 19.16% (17.87-20.45) | 18.44% (17.05-19.78) | -3.76% (-5.65--1.88) | 9.18% (4.66-14.18) |
| **1956** | White | 15.84% (14.64-17.05) | 15.59% (14.33-16.75) | -1.53% (-3.89-0.8) | 5.38% (-2.98-14.25) |
| **1957** | White | 13.22% (12.2-14.19) | 13.34% (12.3-14.38) | 0.9% (-1.47-3.37) | -6.49% (-31.8-13.14) |

| | | | | | |
|---|---|---|---|---|---|
| **1958** | White | 15.68% (14.48-16.84) | 15.48% (14.29-16.65) | -1.29% (-3.59-1.07) | 4.86% (-3.92-13.35) |
| **1959** | White | 24.27% (22.64-25.88) | 23.23% (21.61-24.83) | -4.27% (-6.29--2.52) | 7.98% (4.86-11.69) |
| **1960** | White | 18.09% (16.73-19.53) | 17.67% (16.31-19.01) | -2.31% (-4.51--0.13) | 6.25% (0.36-12.4) |
| **1961** | White | 16.83% (15.63-18.08) | 16.52% (15.32-17.78) | -1.85% (-4.13-0.33) | 5.76% (-0.98-12.8) |
| **1962** | White | 19.11% (17.79-20.49) | 18.78% (17.46-20.02) | -1.71% (-3.96-0.58) | 4.38% (-1.44-9.41) |
| **1963** | White | 15.87% (14.78-16.89) | 16.01% (14.9-17.03) | 0.9% (-1.71-3.37) | -3.37% (-12.94-5.86) |
| **1964** | White | 15.47% (14.42-16.68) | 15.74% (14.53-16.82) | 1.76% (-0.64-4.47) | -6.82% (-17.21-2.46) |
| **1965** | White | 18.78% (17.21-20.42) | 18.76% (17.13-20.37) | -0.13% (-2.61-2.33) | 0.4% (-6.17-6.35) |
| **1966** | White | 22.58% (20.33-25.06) | 22.24% (19.9-24.68) | -1.5% (-3.97-0.83) | 3.03% (-1.7-8.14) |

#### Section 2.2: Smoking

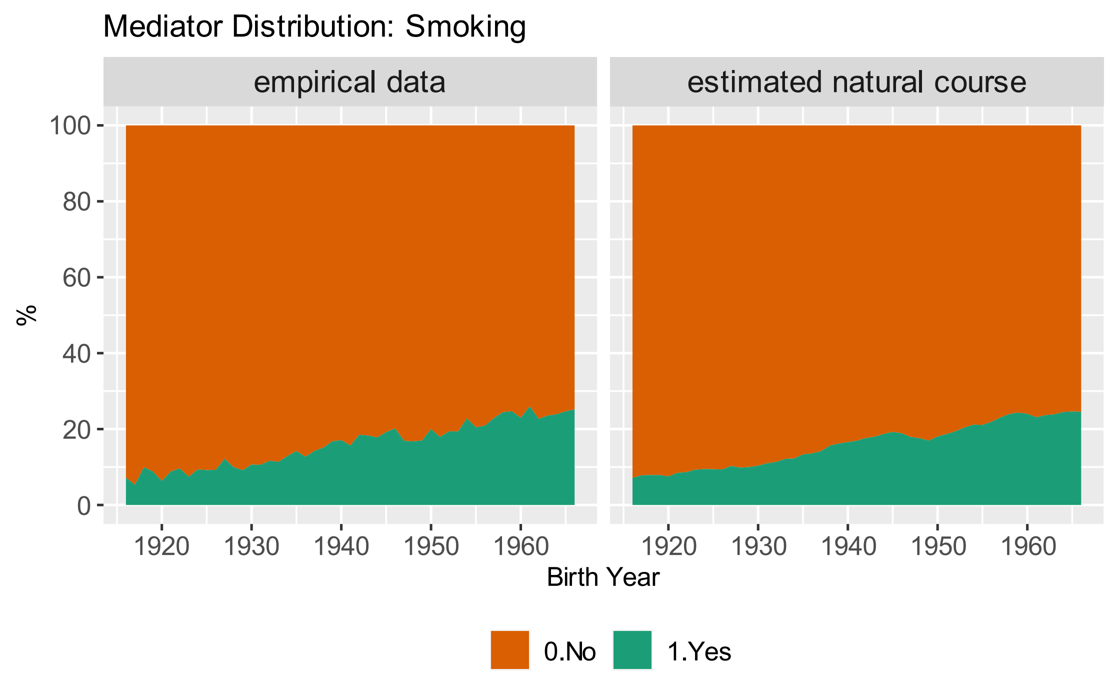

*Figure S. 10 Natural course distribution of smoking by birth cohort. "Empirical data" shows the descriptive distribution as observed in the data. "Estimated natural course" is estimated based on the mediator model without holding age and period constant.*

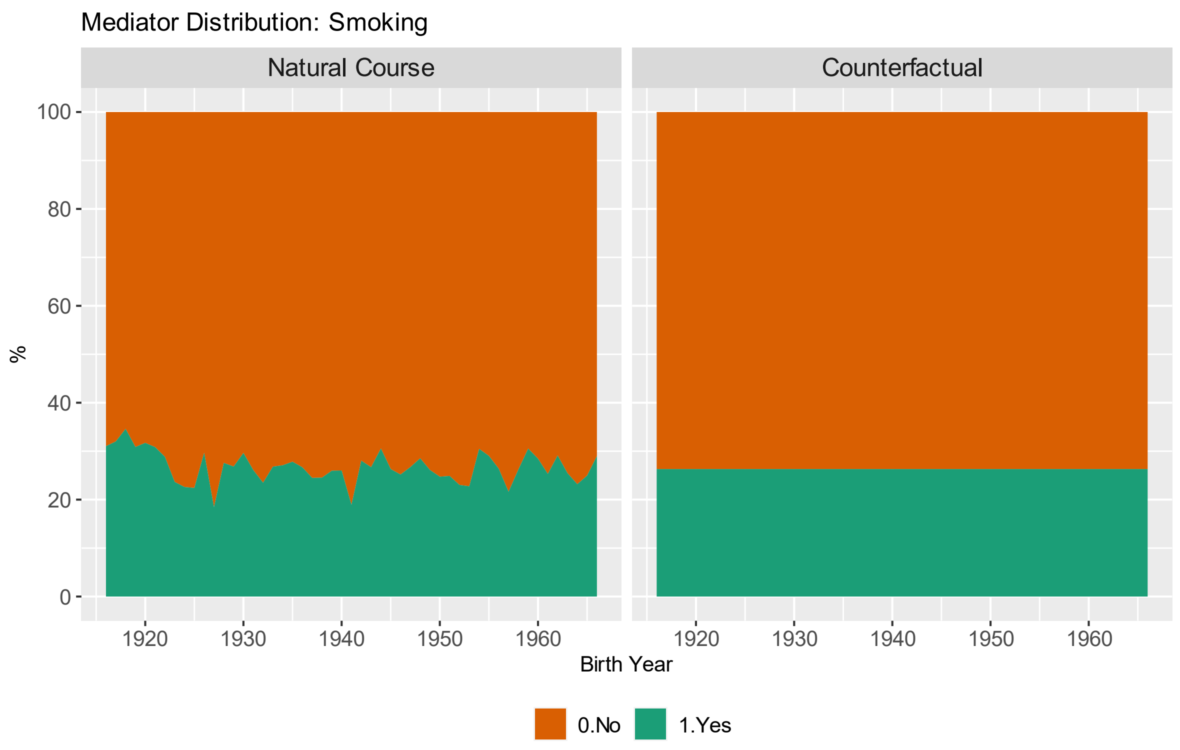

*Figure S. 9 Estimated distribution of smoking categories in natural course and counterfactual scenario by birth year. Age and period are held constant at age 50 and 1996 respectively.*

*Table S. 5 Predicted Probability of elevated depressive symptoms in natural course and counterfactual scenario, relative difference, and contribution by birth cohort. All estimates are presented with 95% confidence intervals. Positive contributions imply that the counterfactual decreases depression risk and negative contributions imply that the counterfactual increases depression risk*

| | **Probability of elevated depressive symptoms (% (95%CI))** | | **% relative difference** | **% contribution** |
|---|---|---|---|---|
| **Birth Cohort** | Natural Course Scenario | Counterfactual Scenario | | |
| **1916** | 20.45% (18.51-22.27) | 20% (18.25-21.99) | -2.2% (-4.82-0.32) | 7.03% (-0.94-14.95) |
| **1917** | 19.77% (18.31-21.24) | 19.08% (17.73-20.57) | -3.53% (-5.81--1.24) | 11.8% (4.48-19.86) |
| **1918** | 22.55% (21.12-23.97) | 21.79% (20.46-23.22) | -3.36% (-5.51--1.18) | 8.63% (3.16-14.24) |
| **1919** | 16.56% (15.54-17.58) | 16.22% (15.26-17.26) | -2.02% (-4.31-0.29) | 12.37% (-1.85-27.41) |
| **1920** | 19.44% (18.29-20.72) | 18.93% (17.81-20.08) | -2.6% (-4.76--0.24) | 9.05% (0.85-15.94) |
| **1921** | 17.49% (16.4-18.62) | 17.01% (15.94-18.1) | -2.71% (-5.01--0.22) | 13.14% (1.15-24.57) |
| **1922** | 20.16% (19.06-21.3) | 19.79% (18.69-20.92) | -1.8% (-3.95-0.51) | 5.98% (-1.66-12.52) |
| **1923** | 15% (14.13-15.94) | 15.12% (14.27-16.07) | 0.79% (-1.73-3.27) | -9.61% (-84.06-23.39) |
| **1924** | 18.36% (17.37-19.31) | 18.41% (17.48-19.32) | 0.27% (-2.12-2.78) | -1% (-12.64-8.58) |
| **1925** | 16% (15.01-16.94) | 16.16% (15.27-17.08) | 0.98% (-1.24-3.33) | -7.21% (-31.99-8.77) |
| **1926** | 18.26% (17.22-19.19) | 17.94% (16.95-18.92) | -1.73% (-3.96-0.47) | 7.19% (-1.8-16.15) |
| **1927** | 14.6% (13.69-15.43) | 15.06% (14.14-15.96) | 3.11% (0.42-5.48) | -54.59% (-509.39-246.62) |

| | | | | |
|---|---|---|---|---|
| **1928** | 18.74% (17.71-19.77) | 18.42% (17.4-19.29) | -1.7% (-3.85-0.41) | 6.49% (-1.62-15.15) |
| **1929** | 20.74% (19.6-21.89) | 20.65% (19.53-21.71) | -0.42% (-2.85-1.87) | 1.15% (-5.69-8.33) |
| **1930** | 20.08% (18.94-21.21) | 19.82% (18.79-20.85) | -1.27% (-3.57-0.92) | 4.22% (-2.95-11.49) |
| **1931** | 16.72% (15.69-17.7) | 16.55% (15.6-17.5) | -1.05% (-3.31-1.21) | 6.06% (-8.13-19.63) |
| **1932** | 17.81% (16.7-18.9) | 17.65% (16.68-18.61) | -0.91% (-3.22-1.38) | 4.09% (-6.4-14.3) |
| **1933** | 14.43% (13.56-15.23) | 14.3% (13.48-15.16) | -0.89% (-3.34-1.52) | 20.52% (-199.67-240.6) |
| **1934** | 15.96% (15.02-16.87) | 15.93% (14.93-16.93) | -0.21% (-2.53-2.12) | 1.53% (-16.45-19.97) |
| **1935** | 15.1% (14.13-16.01) | 14.95% (14.01-15.93) | -1.02% (-3.28-1.42) | 12.13% (-22.53-53.27) |
| **1936** | 16.21% (15.15-17.24) | 16.03% (15.05-17.05) | -1.12% (-3.36-1.44) | 7.43% (-11.96-23.88) |
| **1937** | 14.26% (13.43-15.06) | 14.26% (13.42-15.09) | 0% (-2.47-2.64) | 4.44% (-252.23-305.47) |
| **1938** | 14.78% (13.94-15.6) | 14.86% (13.96-15.75) | 0.59% (-1.86-3.12) | -6.72% (-149.33-35.86) |
| **1939** | 17.98% (17.02-18.97) | 17.75% (16.81-18.71) | -1.26% (-3.44-1.09) | 5.64% (-5.5-14.9) |
| **1940** | 15.27% (14.33-16.15) | 15.13% (14.22-16.02) | -0.9% (-3.52-1.55) | 9.86% (-25.55-44.75) |
| **1941** | 12.52% (11.67-13.35) | 12.89% (12.02-13.73) | 2.95% (0.66-5.37) | 28.5% (5.43-83.99) |
| **1942** | 21.55% (20.36-22.73) | 21.23% (20.07-22.39) | -1.49% (-3.74-0.76) | 4.08% (-2.22-10.28) |
| **1943** | 16.76% (15.77-17.75) | 16.73% (15.75-17.73) | -0.16% (-2.33-2.05) | 0.99% (-13.18-12.86) |
| **1944** | 16.17% (15.24-17.1) | 15.81% (14.94-16.71) | -2.27% (-4.45-0.19) | 15.66% (-1.33-32.76) |

| | | | | |
|---|---|---|---|---|
| **1945** | 13.83% (12.97-14.71) | 13.83% (12.97-14.71) | 0% (0-0) | 0% (0-0) |
| **1946** | 17.48% (16.38-18.53) | 17.46% (16.4-18.49) | -0.08% (-2.17-2.1) | 0.65% (-10.51-10.48) |
| **1947** | 16.63% (15.56-17.66) | 16.32% (15.37-17.33) | -1.85% (-4.11-0.26) | 11.14% (-1.57-25.57) |
| **1948** | 18.55% (17.57-19.49) | 18.19% (17.16-19.13) | -1.92% (-4.05-0.25) | 7.68% (-0.95-15.42) |
| **1949** | 19.06% (18.05-20.03) | 18.88% (17.9-19.87) | -0.93% (-3.27-1.22) | 3.29% (-4.42-11.46) |
| **1950** | 20.66% (19.61-21.81) | 20.44% (19.39-21.56) | -1.06% (-3.41-1.33) | 3.25% (-4.18-10.28) |
| **1951** | 16.52% (15.6-17.54) | 16.48% (15.53-17.52) | -0.23% (-2.73-2.16) | 1.18% (-13.93-16.44) |
| **1952** | 17.7% (16.77-18.76) | 17.73% (16.67-18.77) | 0.21% (-2.26-2.6) | -1.07% (-12.45-9.79) |
| **1953** | 18.36% (17.39-19.34) | 18.45% (17.45-19.46) | 0.48% (-1.81-2.52) | -1.92% (-11.35-7.23) |
| **1954** | 24.3% (23-25.43) | 23.83% (22.62-25) | -1.91% (-3.9-0.42) | 4.56% (-1.01-8.96) |
| **1955** | 22.96% (21.84-24.08) | 22.57% (21.49-23.64) | -1.7% (-3.87-0.38) | 4.34% (-0.95-9.62) |
| **1956** | 18.98% (17.91-19.93) | 18.91% (17.86-19.88) | -0.35% (-2.43-1.9) | 1.43% (-7.39-8.65) |
| **1957** | 14.5% (13.74-15.25) | 14.68% (13.85-15.46) | 1.2% (-1.2-3.47) | -22.25% (-302.63-194.69) |
| **1958** | 17.86% (16.91-18.82) | 17.81% (16.89-18.72) | -0.26% (-2.57-2.07) | 1.16% (-9.75-11.07) |
| **1959** | 28.28% (27.09-29.57) | 27.81% (26.58-29.07) | -1.67% (-3.5-0.16) | 3.3% (-0.31-6.77) |
| **1960** | 21.56% (20.49-22.66) | 21.26% (20.16-22.38) | -1.4% (-3.51-0.87) | 3.77% (-2.5-9.56) |
| **1961** | 19.76% (18.81-20.73) | 19.54% (18.57-20.49) | -1.14% (-3.19-0.83) | 3.81% (-2.77-10.56) |

| | | | | |
|---|---|---|---|---|
| **1962** | 22.17% (21.11-23.18) | 21.79% (20.74-22.77) | -1.7% (-3.77-0.54) | 4.55% (-1.47-9.7) |
| **1963** | 17.15% (16.39-17.8) | 17.1% (16.33-17.82) | -0.24% (-2.35-2.02) | 1.18% (-11.16-11.33) |
| **1964** | 16.75% (15.97-17.42) | 16.89% (16.16-17.63) | 0.84% (-1.4-3.05) | -5.02% (-19.12-7.66) |
| **1965** | 21.03% (20-22.02) | 20.94% (19.93-21.94) | -0.43% (-2.48-1.79) | 1.31% (-5.31-7.1) |
| **1966** | 22.85% (21.3-24.26) | 22.56% (21.02-23.87) | -1.26% (-3.44-0.93) | 3.22% (-2.33-8.48) |

*Table S. 6 Predicted Probability of elevated depressive symptoms in natural course and counterfactual scenario, relative difference, and contribution by birth cohort and* ***stratified by sex.*** *All estimates are presented with 95% confidence intervals. Positive contributions imply that the counterfactual decreases depression risk and negative contributions imply that the counterfactual increases depression risk.*

| | | **Probability of elevated depressive symptoms (% (95%CI))** | | **% relative difference** | **% contribution** |
|---|---|---|---|---|---|
| **Birth Cohort** | sex | Natural Course Scenario | Counterfactual Scenario | | |
| **1916** | female | 21.33% (19.05-23.82) | 21.14% (18.94-23.61) | -0.92% (-3.35-1.54) | 3.2% (-5.92-11.94) |
| **1917** | female | 23.68% (21.58-25.82) | 23.35% (21.36-25.44) | -1.38% (-3.67-1) | 4.04% (-2.94-10.08) |
| **1918** | female | 25.27% (23.29-27.37) | 24.7% (22.81-26.76) | -2.27% (-4.31--0.21) | 5.5% (0.54-10.58) |
| **1919** | female | 20.06% (18.42-21.74) | 19.86% (18.22-21.58) | -1.01% (-3.22-1.15) | 4.04% (-4.44-13.48) |
| **1920** | female | 20.81% (19.06-22.57) | 20.3% (18.52-22.14) | -2.44% (-4.61--0.35) | 8.67% (1.35-17.72) |
| **1921** | female | 20.39% (18.59-22.09) | 19.98% (18.21-21.64) | -2% (-4.33-0.21) | 7.8% (-0.83-17.04) |
| **1922** | female | 20.95% (19.06-22.63) | 20.59% (18.84-22.31) | -1.74% (-3.82-0.44) | 6.17% (-1.55-13.87) |

| | | | | | |
|---|---|---|---|---|---|
| **1923** | female | 16.51% (15-17.93) | 16.56% (15.07-17.94) | 0.36% (-1.94-2.71) | -3.84% (-58.65-37.54) |
| **1924** | female | 20.96% (19.33-22.51) | 21.07% (19.5-22.58) | 0.52% (-1.34-2.42) | -1.81% (-9.05-4.86) |
| **1925** | female | 17.72% (16.3-19.1) | 17.93% (16.52-19.34) | 1.21% (-0.95-3.48) | -7.79% (-27.7-5.96) |
| **1926** | female | 22.23% (20.65-23.76) | 22.08% (20.53-23.63) | -0.71% (-2.83-1.4) | 2.31% (-4.55-8.66) |
| **1927** | female | 15.61% (14.35-16.87) | 16.02% (14.68-17.28) | 2.67% (0.53-5.12) | -45.71% (-808.4-520.79) |
| **1928** | female | 21.49% (20.02-22.95) | 21.3% (19.84-22.77) | -0.86% (-2.88-1.36) | 2.81% (-4.75-9.31) |
| **1929** | female | 22.58% (21.03-24.12) | 22.5% (20.88-24.01) | -0.34% (-2.18-1.8) | 0.79% (-5.17-6.68) |
| **1930** | female | 21.52% (20.02-23.1) | 21.35% (19.83-22.88) | -0.79% (-2.74-1.4) | 2.73% (-4.86-9.05) |
| **1931** | female | 18.78% (17.43-20.14) | 18.64% (17.23-20) | -0.72% (-2.81-1.46) | 3.5% (-8.47-14.97) |
| **1932** | female | 18.95% (17.65-20.22) | 18.89% (17.47-20.2) | -0.31% (-2.27-1.82) | 1.45% (-10.24-11.9) |
| **1933** | female | 15.86% (14.62-17) | 15.72% (14.45-16.89) | -0.88% (-2.95-1.23) | 13.54% (-96-177.73) |
| **1934** | female | 16.08% (14.79-17.26) | 16.11% (14.83-17.26) | 0.21% (-1.97-2.29) | -3.12% (-104.45-65.81) |
| **1935** | female | 15.82% (14.59-17.01) | 15.76% (14.49-16.9) | -0.36% (-2.49-2.1) | 6.8% (-203.02-258.18) |
| **1936** | female | 16.68% (15.36-18.03) | 16.58% (15.22-17.84) | -0.57% (-2.68-1.72) | 5.66% (-28.82-43.72) |
| **1937** | female | 15.57% (14.35-16.7) | 15.6% (14.34-16.72) | 0.21% (-2.16-2.48) | -0.31% (-193.57-164.57) |
| **1938** | female | 15.43% (14.18-16.61) | 15.55% (14.27-16.7) | 0.82% (-1.19-3.17) | -9.81% (-384.45-199.41) |
| **1939** | female | 19.68% (18.34-20.96) | 19.59% (18.19-20.91) | -0.44% (-2.53-1.65) | 2.09% (-7.47-10.4) |

| | | | | | |
|---|---|---|---|---|---|
| **1940** | female | 16.74% (15.53-17.91) | 16.67% (15.43-17.86) | -0.42% (-2.52-1.75) | 4.69% (-25.88-34.35) |
| **1941** | female | 13.42% (12.36-14.49) | 13.79% (12.68-14.92) | 2.81% (0.53-5.24) | 23.65% (3.79-79.14) |
| **1942** | female | 21.91% (20.4-23.43) | 21.59% (20.17-23.01) | -1.47% (-3.49-0.6) | 4.51% (-1.94-11.1) |
| **1943** | female | 17.49% (16.29-18.69) | 17.42% (16.21-18.63) | -0.41% (-2.43-1.7) | 3.26% (-14.37-18.54) |
| **1944** | female | 18.24% (16.99-19.56) | 17.81% (16.56-18.97) | -2.37% (-4.44--0.18) | 13.49% (1.07-26.91) |
| **1945** | female | 15.01% (13.86-16.08) | 15.01% (13.86-16.08) | 0% (0-0) | 0% (0-0) |
| **1946** | female | 17.63% (16.27-18.82) | 17.54% (16.23-18.75) | -0.5% (-2.61-1.72) | 3.64% (-13.49-18.07) |
| **1947** | female | 19.89% (18.42-21.23) | 19.7% (18.35-21.03) | -0.96% (-3.04-1.14) | 3.79% (-4.78-12.14) |
| **1948** | female | 19.28% (18-20.56) | 18.98% (17.65-20.25) | -1.54% (-3.78-0.57) | 6.63% (-2.68-17.3) |
| **1949** | female | 19.21% (17.94-20.46) | 19.03% (17.77-20.23) | -0.94% (-2.95-1.14) | 4.26% (-5.41-14.62) |
| **1950** | female | 19.61% (18.28-20.95) | 19.45% (18.13-20.72) | -0.85% (-2.92-1.32) | 3.74% (-5.58-12.33) |
| **1951** | female | 17.52% (16.3-18.8) | 17.61% (16.33-18.86) | 0.51% (-1.5-2.81) | -3.75% (-25.07-11.33) |
| **1952** | female | 18.53% (17.15-19.72) | 18.62% (17.24-19.88) | 0.5% (-1.76-2.49) | -2.68% (-13.98-8.76) |
| **1953** | female | 19.96% (18.57-21.32) | 20.24% (18.83-21.52) | 1.43% (-0.79-3.5) | -5.95% (-15.3-3.22) |
| **1954** | female | 26.49% (24.86-28.2) | 26.24% (24.66-27.85) | -0.91% (-2.86-1.02) | 1.92% (-2.39-6.49) |
| **1955** | female | 24.21% (22.74-25.77) | 24.01% (22.5-25.42) | -0.83% (-2.83-1.04) | 2.25% (-2.82-7.31) |
| **1956** | female | 21.61% (20.21-23.05) | 21.71% (20.31-23.08) | 0.45% (-1.49-2.52) | -1.32% (-8.38-4.74) |

| | | | | | |
|---|---|---|---|---|---|
| **1957** | female | 16.48% (15.35-17.62) | 16.71% (15.55-17.88) | 1.39% (-0.82-3.56) | -15.81% (-88.23-9.47) |
| **1958** | female | 21.61% (20.22-23.06) | 21.6% (20.18-23.04) | -0.06% (-1.97-1.95) | 0.14% (-6.88-6.14) |
| **1959** | female | 29.7% (27.83-31.4) | 29.25% (27.44-30.97) | -1.51% (-3.11-0.3) | 3.08% (-0.61-6.4) |
| **1960** | female | 23.26% (21.64-24.9) | 22.95% (21.34-24.54) | -1.31% (-3.25-1.07) | 3.59% (-3.05-9.27) |
| **1961** | female | 21.52% (20.03-22.95) | 21.29% (19.78-22.68) | -1.03% (-2.98-0.92) | 3.44% (-3.28-9.87) |
| **1962** | female | 24.32% (22.74-25.73) | 23.93% (22.4-25.33) | -1.6% (-3.59-0.45) | 4.24% (-1.17-9.29) |
| **1963** | female | 20.82% (19.43-22.13) | 20.76% (19.39-22.03) | -0.3% (-2.28-1.83) | 1.07% (-7.01-8.12) |
| **1964** | female | 19.61% (18.27-20.85) | 19.68% (18.38-20.96) | 0.34% (-1.79-2.55) | -1.41% (-11.3-7.84) |
| **1965** | female | 23.95% (22.18-25.67) | 23.82% (21.99-25.64) | -0.54% (-2.57-1.35) | 1.39% (-3.64-7.12) |
| **1966** | female | 25.73% (23.37-28) | 25.23% (22.95-27.47) | -1.93% (-4.08-0.19) | 4.68% (-0.46-9.67) |
| **1916** | male | 21.99% (19.35-25.07) | 21.02% (18.46-24.09) | -4.41% (-7.37--1.69) | 11.28% (4.21-19.58) |
| **1917** | male | 22.97% (20.78-25.61) | 22.06% (19.9-24.73) | -3.95% (-6.76--1.7) | 9.36% (3.9-16.46) |
| **1918** | male | 24.51% (22.29-27.05) | 23.42% (21.27-26) | -4.44% (-6.78--1.94) | 9.61% (3.99-14.97) |
| **1919** | male | 19.14% (17.41-21.13) | 18.7% (17.01-20.67) | -2.27% (-4.71-0.53) | 7.61% (-1.62-15.79) |
| **1920** | male | 19.71% (17.79-21.7) | 18.95% (17.24-20.8) | -3.87% (-6.44--1.32) | 11.77% (4.39-20.46) |
| **1921** | male | 18.27% (16.6-20.18) | 17.73% (16.11-19.59) | -2.92% (-5.58--0.41) | 10.65% (1.49-21.4) |
| **1922** | male | 18.38% (16.6-20.29) | 17.79% (16.16-19.68) | -3.25% (-5.94--0.76) | 11.61% (2.95-22.17) |

| | | | | | |
|---|---|---|---|---|---|
| **1923** | male | 14.33% (12.93-15.8) | 14.46% (13.11-16) | 0.94% (-2.01-4.05) | -8.29% (-154.99-182.11) |
| **1924** | male | 17.11% (15.6-18.73) | 17.04% (15.58-18.7) | -0.42% (-3.12-2.31) | 2.03% (-11.51-13.8) |
| **1925** | male | 14.64% (13.34-16.05) | 14.56% (13.28-15.98) | -0.56% (-3.09-1.86) | 5.4% (-53.04-83.1) |
| **1926** | male | 18.13% (16.57-19.67) | 17.63% (16.17-19.13) | -2.75% (-5.37--0.1) | 10.43% (0.45-21.37) |
| **1927** | male | 13.31% (12.12-14.55) | 13.63% (12.43-14.9) | 2.41% (-0.45-5.16) | -6.35% (-699.85-1132.52) |
| **1928** | male | 18.11% (16.64-19.54) | 17.59% (16.15-19.05) | -2.85% (-5.38--0.18) | 10.62% (0.68-19.95) |
| **1929** | male | 19.49% (17.79-21.25) | 19.19% (17.62-20.85) | -1.51% (-4.18-1.15) | 4.68% (-4.07-13.18) |
| **1930** | male | 19.47% (17.76-21.18) | 18.93% (17.32-20.58) | -2.73% (-5.15--0.22) | 8.35% (0.75-16.99) |
| **1931** | male | 17.63% (16.13-19.24) | 17.38% (15.92-18.87) | -1.41% (-3.91-1.15) | 5.55% (-4.54-16.51) |
| **1932** | male | 17.44% (16.01-18.96) | 17.17% (15.74-18.69) | -1.55% (-4.07-1.01) | 6.78% (-4.99-18.22) |
| **1933** | male | 15.26% (13.93-16.69) | 15.16% (13.86-16.54) | -0.59% (-3.25-2.01) | 4.71% (-24.18-29.05) |
| **1934** | male | 16.13% (14.69-17.7) | 15.89% (14.53-17.42) | -1.52% (-4.44-1.07) | 7.95% (-6.52-26.2) |
| **1935** | male | 15.76% (14.4-17.33) | 15.5% (14.24-17) | -1.68% (-4.44-0.98) | 10.64% (-7.42-31.12) |
| **1936** | male | 16.33% (15.02-17.91) | 16.02% (14.71-17.45) | -1.92% (-4.57-0.7) | 10.16% (-4.1-26.56) |
| **1937** | male | 14.92% (13.67-16.26) | 14.86% (13.62-16.22) | -0.41% (-3.04-2.2) | 4.11% (-48.02-36.48) |
| **1938** | male | 14.99% (13.73-16.28) | 14.88% (13.66-16.19) | -0.73% (-3.38-2.07) | 6.23% (-26.35-38.8) |
| **1939** | male | 18.32% (16.88-19.79) | 17.79% (16.41-19.24) | -2.84% (-5.43--0.28) | 10.37% (1.05-20.27) |

| | | | | | |
|---|---|---|---|---|---|
| **1940** | male | 15.84% (14.6-17.22) | 15.63% (14.4-17) | -1.3% (-3.9-1.06) | 8.12% (-9.01-28.17) |
| **1941** | male | 12.6% (11.52-13.74) | 12.91% (11.75-14.15) | 2.53% (-0.46-5.02) | 34.34% (-504.76-447.42) |
| **1942** | male | 19.58% (18.05-21.23) | 19.04% (17.5-20.67) | -2.72% (-5.12--0.28) | 8.48% (0.88-16.11) |
| **1943** | male | 15.76% (14.43-17.1) | 15.59% (14.28-16.96) | -1.06% (-3.72-1.44) | 6.08% (-14.08-26.93) |
| **1944** | male | 15.6% (14.34-16.99) | 15.24% (14.02-16.57) | -2.3% (-4.86-0.37) | 15.46% (-3.71-39.81) |
| **1945** | male | 13.27% (12.08-14.46) | 13.27% (12.08-14.46) | 0% (0-0) | 0% (0-0) |
| **1946** | male | 15.26% (13.95-16.71) | 15.15% (13.83-16.56) | -0.73% (-3.43-1.71) | 4.91% (-18.99-33.11) |
| **1947** | male | 17.59% (16.13-19.14) | 17.26% (15.83-18.73) | -1.85% (-4.23-0.48) | 7.61% (-2.11-17.12) |
| **1948** | male | 17.91% (16.55-19.45) | 17.5% (16.11-18.96) | -2.29% (-5.15-0.22) | 8.69% (-0.81-19.25) |
| **1949** | male | 19.13% (17.7-20.67) | 18.69% (17.23-20.13) | -2.3% (-4.59--0.04) | 7.74% (0.13-15.11) |
| **1950** | male | 20.42% (18.88-21.89) | 19.89% (18.45-21.4) | -2.57% (-5.1-0.03) | 7.09% (-0.08-14.35) |
| **1951** | male | 18.94% (17.48-20.46) | 18.78% (17.4-20.26) | -0.79% (-3.27-1.85) | 2.71% (-6.63-11) |
| **1952** | male | 19.24% (17.85-20.68) | 19.15% (17.79-20.64) | -0.47% (-2.93-1.94) | 1.54% (-6.74-9.17) |
| **1953** | male | 19.95% (18.52-21.47) | 19.9% (18.5-21.34) | -0.26% (-2.53-2.2) | 0.87% (-6.86-7.59) |
| **1954** | male | 25.48% (23.88-27.28) | 24.72% (23.12-26.53) | -2.98% (-5.22--0.92) | 6.19% (1.99-11.09) |
| **1955** | male | 22.42% (20.89-24.3) | 21.66% (20.19-23.39) | -3.37% (-5.96--1.01) | 8.17% (2.44-14.21) |
| **1956** | male | 19.49% (18.08-21.12) | 19.3% (17.93-20.86) | -0.99% (-3.38-1.4) | 3.21% (-4.47-10.22) |

| | | | | | |
|---|---|---|---|---|---|
| **1957** | male | 14.61% (13.56-15.78) | 14.81% (13.74-16.01) | 1.36% (-1.33-4.03) | -15.24% (-91.88-17.03) |
| **1958** | male | 18.28% (16.99-19.79) | 18.22% (16.89-19.63) | -0.3% (-2.87-2.27) | 1.09% (-8.85-10.16) |
| **1959** | male | 24.52% (22.85-26.27) | 23.72% (22.14-25.36) | -3.25% (-5.56--0.76) | 7.15% (1.64-12) |
| **1960** | male | 20.13% (18.62-21.7) | 19.7% (18.24-21.24) | -2.14% (-4.85-0.38) | 6.16% (-1.09-14.04) |
| **1961** | male | 18.1% (16.77-19.49) | 17.83% (16.55-19.18) | -1.46% (-4.14-1.07) | 5.14% (-4.55-15.61) |
| **1962** | male | 20.46% (19.06-22.01) | 20.1% (18.74-21.54) | -1.75% (-4.33-0.57) | 4.87% (-1.68-12.24) |
| **1963** | male | 17.08% (15.94-18.33) | 17.34% (16.17-18.6) | 1.53% (-1.1-3.98) | -7.34% (-19.36-4.68) |
| **1964** | male | 16.15% (15.07-17.45) | 16.49% (15.32-17.69) | 2.1% (-0.67-4.72) | -11.76% (-31.42-3.86) |
| **1965** | male | 18.73% (17.19-20.45) | 18.84% (17.24-20.62) | 0.59% (-2.02-3.32) | -1.95% (-13.04-6.76) |
| **1966** | male | 19.83% (17.67-22.1) | 19.85% (17.67-22.29) | 0.09% (-2.66-2.87) | -0.37% (-9.39-7.99) |

*Table S. 7 Predicted Probability of elevated depressive symptoms in natural course and counterfactual scenario, relative difference, and contribution by birth cohort and **stratified by race/ethnicity**. All estimates are presented with 95% confidence intervals. Positive contributions imply that the counterfactual decreases depression risk and negative contributions imply that the counterfactual increases depression risk*

| | | **Probability of elevated depressive symptoms (%(95%CI))** | | **% relative difference** | **% contribution** |
|---|---|---|---|---|---|
| **Birth Cohort** | race/ethnicity | Natural Course Scenario | Counterfactual Scenario | | |
| **1916** | Black | 34.43% (31.33-37.8) | 32.99% (29.9-36.23) | -4.21% (-6.24--2.45) | 10.3% (5.57-15.22) |
| **1917** | Black | 31.78% (29.37-34.28) | 30.17% (27.77-32.65) | -5.08% (-6.73--3.35) | 14.11% (9.17-19.13) |

| | | | | | |
|---|---|---|---|---|---|
| **1918** | Black | 34.3% (31.85-36.59) | 32.91% (30.5-35.21) | -4.04% (-5.62--2.38) | 9.9% (5.85-13.82) |
| **1919** | Black | 26.09% (24.2-27.95) | 25.32% (23.43-27.16) | -2.95% (-4.7--1.15) | 13.2% (4.99-22.9) |
| **1920** | Black | 28.76% (26.76-30.87) | 28.36% (26.29-30.43) | -1.39% (-3.09-0.36) | 4.73% (-1.28-10.38) |
| **1921** | Black | 26.06% (24.13-28.12) | 25.66% (23.71-27.76) | -1.53% (-3.28-0.11) | 6.84% (-0.52-14.91) |
| **1922** | Black | 28.12% (26.16-30.2) | 28.08% (26.1-30.16) | -0.14% (-1.8-1.57) | 0.53% (-5.81-6.77) |
| **1923** | Black | 23.08% (21.35-24.86) | 23.15% (21.37-24.91) | 0.3% (-1.48-2) | -2.56% (-23.08-13.89) |
| **1924** | Black | 24.8% (23.17-26.48) | 24.68% (22.95-26.5) | -0.48% (-2.13-1.19) | 2.87% (-7.12-12.39) |
| **1925** | Black | 21.26% (19.77-22.76) | 21.42% (19.94-22.96) | 0.8% (-1-2.63) | -13.2% (-205.48-148.78) |
| **1926** | Black | 23.79% (22.19-25.26) | 23.26% (21.71-24.76) | -2.2% (-3.85--0.52) | 14.86% (3.4-29.68) |
| **1927** | Black | 19.04% (17.63-20.41) | 19.38% (18.05-20.75) | 1.8% (-0.07-3.65) | 25.88% (-117.27-330.55) |
| **1928** | Black | 24.19% (22.72-25.68) | 23.96% (22.5-25.45) | -0.95% (-2.72-0.73) | 5.75% (-4.43-17.42) |
| **1929** | Black | 26.37% (24.74-28.02) | 26.23% (24.58-27.85) | -0.55% (-2.09-1.17) | 2.38% (-5.25-9.41) |
| **1930** | Black | 24.3% (22.7-25.87) | 24.37% (22.87-25.96) | 0.31% (-1.31-1.97) | -1.49% (-13.11-8.19) |
| **1931** | Black | 21.42% (19.94-22.99) | 21.42% (20.01-23.02) | 0.03% (-1.78-1.63) | 0.48% (-109.68-64.24) |
| **1932** | Black | 21.97% (20.53-23.42) | 22.14% (20.71-23.68) | 0.78% (-0.9-2.35) | -10.21% (-58.78-14.19) |
| **1933** | Black | 20.97% (19.58-22.53) | 20.98% (19.6-22.54) | 0.04% (-1.76-1.9) | 1.89% (-156.19-249.63) |
| **1934** | Black | 19.74% (18.39-21.29) | 20.03% (18.61-21.65) | 1.47% (-0.3-3.24) | 22.81% (-669.71-403.46) |

| | | | | | |
|---|---|---|---|---|---|
| **1935** | Black | 19.95% (18.55-21.49) | 20.01% (18.57-21.64) | 0.31% (-1.51-2.18) | 5.83% (-220.8-268.87) |
| **1936** | Black | 21.19% (19.72-22.84) | 21.11% (19.63-22.7) | -0.4% (-2.21-1.56) | 9.12% (-100.61-132.74) |
| **1937** | Black | 19.52% (18.24-20.94) | 19.39% (18.02-20.85) | -0.62% (-2.46-1.33) | -9.38% (-264.6-199.79) |
| **1938** | Black | 18.26% (16.96-19.55) | 18.18% (16.89-19.58) | -0.43% (-2.25-1.47) | -3.94% (-37.06-15.21) |
| **1939** | Black | 20.09% (18.75-21.52) | 19.75% (18.44-21.16) | -1.72% (-3.52-0.14) | -15.14% (-997.7-530.49) |
| **1940** | Black | 18.27% (16.94-19.63) | 17.86% (16.59-19.21) | -2.25% (-4.17--0.35) | -20.58% (-82.45--2.3) |
| **1941** | Black | 14.52% (13.41-15.71) | 14.64% (13.52-15.86) | 0.85% (-1.15-2.88) | 2.19% (-2.98-7.05) |
| **1942** | Black | 24.97% (23.31-26.53) | 24.36% (22.83-26.11) | -2.44% (-4.01--0.76) | 12.86% (3.92-24.28) |
| **1943** | Black | 21.04% (19.7-22.63) | 20.83% (19.39-22.47) | -0.99% (-2.92-0.7) | 17.02% (-159.36-321.21) |
| **1944** | Black | 23.83% (22.29-25.57) | 23.31% (21.79-24.99) | -2.17% (-3.79--0.45) | 14.34% (3.34-30.39) |
| **1945** | Black | 20.22% (18.83-21.86) | 20.22% (18.83-21.86) | 0% (0-0) | 0% (0-0) |
| **1946** | Black | 26.21% (24.53-28.11) | 26.15% (24.49-28.07) | -0.23% (-1.84-1.61) | 1.15% (-7.32-7.81) |
| **1947** | Black | 24.46% (22.86-26.17) | 23.85% (22.27-25.51) | -2.53% (-4.2--0.99) | 14.58% (5.58-27.7) |
| **1948** | Black | 27.68% (26.06-29.44) | 27.18% (25.63-28.8) | -1.8% (-3.39--0.14) | 6.65% (0.58-13.51) |
| **1949** | Black | 26.86% (25.22-28.5) | 26.52% (24.93-28.1) | -1.25% (-2.89-0.37) | 5.21% (-1.46-11.95) |
| **1950** | Black | 27.13% (25.57-28.76) | 26.73% (25.2-28.39) | -1.46% (-3.14-0.14) | 5.64% (-0.63-12.68) |
| **1951** | Black | 22.44% (20.99-23.86) | 21.88% (20.5-23.31) | -2.49% (-4.18--0.7) | 24.24% (6.5-65.06) |

| | | | | | |
|---|---|---|---|---|---|
| **1952** | Black | 24.49% (23.02-26.05) | 23.89% (22.37-25.43) | -2.45% (-4.28--0.88) | 13.73% (4.62-27.29) |
| **1953** | Black | 24.63% (23.11-26.11) | 23.98% (22.49-25.46) | -2.65% (-4.12--0.88) | 14.93% (5.11-25.98) |
| **1954** | Black | 32.77% (30.85-34.49) | 31.3% (29.49-33.16) | -4.47% (-5.93--2.92) | 11.82% (7.78-15.9) |
| **1955** | Black | 31.66% (29.82-33.49) | 30.49% (28.78-32.32) | -3.7% (-5.35--2.13) | 10.29% (6.01-14.77) |
| **1956** | Black | 25.8% (24.3-27.37) | 24.87% (23.37-26.55) | -3.57% (-5.28--1.74) | 16.7% (7.93-26.4) |
| **1957** | Black | 21.22% (19.89-22.7) | 20.85% (19.49-22.4) | -1.74% (-3.63-0.06) | 31.76% (-231.38-384.42) |
| **1958** | Black | 24.82% (23.28-26.43) | 23.92% (22.35-25.53) | -3.62% (-5.35--1.74) | 19.67% (10.11-32.87) |
| **1959** | Black | 33.11% (31.23-35.19) | 32.08% (30.24-34.07) | -3.11% (-4.63--1.6) | 8.05% (4.05-12.11) |
| **1960** | Black | 24.75% (23.16-26.49) | 24% (22.35-25.65) | -3.05% (-4.79--1.37) | 16.77% (7.82-29.75) |
| **1961** | Black | 22.06% (20.63-23.51) | 21.61% (20.17-23.14) | -2.04% (-3.82--0.3) | 24.06% (2.96-106.46) |
| **1962** | Black | 23.94% (22.44-25.52) | 23.38% (21.84-24.9) | -2.33% (-3.98--0.52) | 15.49% (3.61-29.33) |
| **1963** | Black | 19.43% (18.28-20.71) | 19.04% (17.88-20.22) | -1.98% (-3.92--0.15) | -38.74% (-351.71-315.11) |
| **1964** | Black | 18.11% (16.94-19.27) | 18.11% (16.98-19.3) | 0.01% (-1.85-1.9) | 0.1% (-21.19-19.05) |
| **1965** | Black | 22.35% (20.95-23.76) | 22.23% (20.8-23.78) | -0.55% (-2.27-1.21) | 6.03% (-14.65-31.99) |
| **1966** | Black | 25.57% (23.7-27.55) | 25.49% (23.65-27.53) | -0.31% (-1.83-1.4) | 1.17% (-7.24-8.87) |
| **1916** | Hispanic | 11.91% (9.74-14.4) | 11.06% (9.11-13.41) | -7.12% (-12.14--2.33) | -8.94% (-19.52--2.85) |
| **1917** | Hispanic | 14.78% (12.8-17.18) | 13.69% (11.83-15.8) | -7.34% (-11.6--3.03) | -17.03% (-36.66--6.08) |

| | | | | | |
|---|---|---|---|---|---|
| **1918** | Hispanic | 22.53% (20.35-24.87) | 21.29% (19.18-23.66) | -5.5% (-8.65--2.16) | 73.48% (-674.8-700.41) |
| **1919** | Hispanic | 21.73% (20.01-23.71) | 21.07% (19.27-23.19) | -3.02% (-6.07--0.07) | 50.54% (-611.06-712.86) |
| **1920** | Hispanic | 29.13% (27.02-31.3) | 28.59% (26.64-30.68) | -1.82% (-4.47-0.76) | 6.63% (-2.88-16.13) |
| **1921** | Hispanic | 30.15% (28.18-32.39) | 29.85% (27.87-31.97) | -0.99% (-3.15-1.45) | 3.46% (-4.97-10.84) |
| **1922** | Hispanic | 33.44% (31.38-35.63) | 33.3% (31.22-35.46) | -0.4% (-2.46-1.64) | 0.89% (-4.67-6.72) |
| **1923** | Hispanic | 26.06% (24.32-27.98) | 25.79% (23.99-27.7) | -1.03% (-3.62-1.43) | 5.59% (-8.06-18.72) |
| **1924** | Hispanic | 25.72% (24.12-27.56) | 25.23% (23.58-26.94) | -1.91% (-4.45-0.53) | 10.63% (-2.79-24.97) |
| **1925** | Hispanic | 21.72% (20.26-23.39) | 21.45% (20.09-23.07) | -1.21% (-4.21-1.72) | 24.62% (-358.59-698.98) |
| **1926** | Hispanic | 22.37% (20.9-23.94) | 21.37% (19.88-22.93) | -4.42% (-7.09--1.52) | 73.77% (-428.35-457.26) |
| **1927** | Hispanic | 17.59% (16.3-19.02) | 17.29% (16-18.71) | -1.67% (-4.55-1.47) | -8.55% (-30.32-6.55) |
| **1928** | Hispanic | 21.32% (19.86-22.85) | 20.43% (19.06-21.96) | -4.13% (-6.95--1.15) | 74.36% (-1371.59-1160.76) |
| **1929** | Hispanic | 25.01% (23.53-26.71) | 24.42% (22.86-26.08) | -2.35% (-5.01-0.37) | 14.62% (-2.86-35.11) |
| **1930** | Hispanic | 23.98% (22.45-25.71) | 23.65% (22.11-25.38) | -1.35% (-3.83-1.23) | 11.39% (-12.89-34.91) |
| **1931** | Hispanic | 20.39% (18.99-22.11) | 19.88% (18.43-21.47) | -2.48% (-5.33-0.55) | -38.65% (-1076.3-432.58) |
| **1932** | Hispanic | 21.8% (20.47-23.41) | 21.68% (20.21-23.29) | -0.52% (-3.24-2.29) | 11.91% (-316.02-513.73) |
| **1933** | Hispanic | 20.54% (19.21-22.05) | 20.33% (18.85-21.98) | -1.04% (-3.81-1.92) | -12.71% (-398.69-416.53) |
| **1934** | Hispanic | 23.77% (22.21-25.5) | 23.78% (22.19-25.52) | 0.02% (-2.35-2.68) | 0.77% (-28.94-23.25) |

| | | | | | |
|---|---|---|---|---|---|
| **1935** | Hispanic | 22.67% (21.15-24.41) | 22.4% (20.89-24.06) | -1.19% (-3.73-1.48) | 16.08% (-47.8-98.7) |
| **1936** | Hispanic | 22.35% (20.9-23.94) | 21.93% (20.43-23.65) | -1.83% (-4.65-0.92) | 30.32% (-157.1-335.18) |
| **1937** | Hispanic | 21.9% (20.35-23.51) | 21.74% (20.21-23.35) | -0.72% (-3.36-2.12) | 15.47% (-260.19-213.22) |
| **1938** | Hispanic | 21.99% (20.57-23.61) | 22% (20.45-23.7) | 0.06% (-2.6-2.72) | 3.5% (-216.71-542.03) |
| **1939** | Hispanic | 22.26% (20.79-24.01) | 21.95% (20.44-23.55) | -1.39% (-4.01-1.18) | 23.71% (-115.55-421.03) |
| **1940** | Hispanic | 21.05% (19.64-22.67) | 20.97% (19.5-22.52) | -0.38% (-2.96-2.48) | 11.74% (-370.4-490.87) |
| **1941** | Hispanic | 17.08% (15.86-18.46) | 17.38% (16.13-18.76) | 1.75% (-1.21-4.64) | 7.22% (-5.59-20.57) |
| **1942** | Hispanic | 26.56% (25.07-28.34) | 26.23% (24.61-28.02) | -1.24% (-3.45-1.34) | 6.23% (-6.72-17.63) |
| **1943** | Hispanic | 24.15% (22.69-25.85) | 24.14% (22.56-25.91) | -0.02% (-2.75-2.76) | 0.38% (-25.5-23.07) |
| **1944** | Hispanic | 24.36% (22.77-26.14) | 24.07% (22.47-25.85) | -1.17% (-3.61-1.46) | 8.78% (-13.29-29.56) |
| **1945** | Hispanic | 21.08% (19.59-22.69) | 21.08% (19.59-22.69) | 0% (0-0) | 0% (0-0) |
| **1946** | Hispanic | 24.18% (22.63-25.91) | 24.16% (22.51-25.83) | -0.08% (-2.71-2.79) | 0.32% (-27.56-21.12) |
| **1947** | Hispanic | 21.55% (20.14-23.17) | 20.85% (19.42-22.35) | -3.26% (-6.03--0.5) | 60.79% (-967.54-853.66) |
| **1948** | Hispanic | 24% (22.66-25.59) | 23.63% (22.21-25.15) | -1.51% (-4.06-0.71) | 12.19% (-6.38-39.17) |
| **1949** | Hispanic | 23.15% (21.7-24.6) | 22.83% (21.43-24.34) | -1.39% (-4.01-1.37) | 16.29% (-18.72-60.13) |
| **1950** | Hispanic | 22.91% (21.51-24.45) | 22.54% (21.17-24.06) | -1.62% (-4.12-1.03) | 21.06% (-20.33-78.85) |
| **1951** | Hispanic | 19.52% (18.24-20.9) | 19.28% (18.05-20.62) | -1.2% (-4.36-1.89) | -13.41% (-159.17-27.49) |

| | | | | | |
|---|---|---|---|---|---|
| **1952** | Hispanic | 19.82% (18.53-21.27) | 19.65% (18.34-20.96) | -0.88% (-3.59-1.59) | -10.39% (-137.9-200.88) |
| **1953** | Hispanic | 20.42% (19.09-21.79) | 20.39% (19.14-21.72) | -0.14% (-2.82-2.57) | -0.04% (-201.02-352.97) |
| **1954** | Hispanic | 25.24% (23.77-26.84) | 24.53% (23.06-26.06) | -2.78% (-5.41--0.23) | 16.77% (1.6-34.86) |
| **1955** | Hispanic | 25.78% (24.22-27.42) | 25.59% (24.06-27.2) | -0.7% (-3.14-1.81) | 4.23% (-11.4-16.8) |
| **1956** | Hispanic | 23.51% (22-25.14) | 23.53% (22.05-25.14) | 0.09% (-2.45-2.92) | -0.95% (-41.15-27.69) |
| **1957** | Hispanic | 19.61% (18.34-21) | 19.7% (18.4-21.08) | 0.48% (-2.1-3.25) | 6.38% (-63.49-105.87) |
| **1958** | Hispanic | 21.65% (20.32-23.16) | 21.44% (20.03-22.9) | -0.98% (-3.48-1.68) | 17.59% (-689.66-295.34) |
| **1959** | Hispanic | 31.13% (29.49-32.96) | 30.8% (29.11-32.55) | -1.04% (-3.28-1.46) | 3.37% (-4.54-9.99) |
| **1960** | Hispanic | 24.47% (22.95-26.19) | 23.9% (22.45-25.55) | -2.3% (-4.77-0.29) | 17.28% (-2.48-37.6) |
| **1961** | Hispanic | 22.49% (21.09-23.99) | 22.22% (20.84-23.78) | -1.17% (-3.89-1.56) | 19.12% (-49.98-153.54) |
| **1962** | Hispanic | 25.48% (23.98-27.09) | 25.02% (23.6-26.55) | -1.81% (-4.34-0.73) | 10.64% (-4.31-26.8) |
| **1963** | Hispanic | 20.96% (19.72-22.21) | 20.61% (19.35-21.88) | -1.63% (-4.16-1.39) | -12.47% (-922.17-860.11) |
| **1964** | Hispanic | 19.64% (18.36-20.86) | 19.31% (18.1-20.51) | -1.67% (-4.52-1.33) | -22.65% (-211.09-20.24) |
| **1965** | Hispanic | 22.26% (20.84-23.63) | 21.96% (20.51-23.37) | -1.34% (-3.77-1.45) | 21.95% (-184.74-227.78) |
| **1966** | Hispanic | 25.11% (23.21-27.12) | 24.61% (22.75-26.47) | -1.99% (-4.79-0.68) | 12.33% (-4.9-33.42) |
| **1916** | White | 16.68% (14.56-18.86) | 16.35% (14.46-18.58) | -1.94% (-5.07-1.06) | 6.02% (-3.52-16.15) |
| **1917** | White | 16.22% (14.52-17.95) | 15.58% (13.99-17.28) | -3.96% (-6.81--1.25) | 13.08% (4.12-22.57) |

| | | | | | |
|---|---|---|---|---|---|
| **1918** | White | 18.34% (16.49-20.26) | 17.49% (15.8-19.25) | -4.62% (-7.05--2.28) | 11.73% (6.04-18.62) |
| **1919** | White | 13.07% (11.8-14.4) | 12.69% (11.43-13.99) | -2.92% (-5.5--0.5) | 20.61% (2.75-61.84) |
| **1920** | White | 15.54% (13.82-17.16) | 15.05% (13.51-16.46) | -3.13% (-5.64--0.62) | 11.35% (2.34-21.17) |
| **1921** | White | 14.17% (12.7-15.58) | 13.68% (12.23-15.04) | -3.49% (-6.06--0.89) | 16.59% (4.34-36.09) |
| **1922** | White | 16.33% (14.71-17.88) | 15.84% (14.28-17.32) | -3.03% (-5.3--0.68) | 9.74% (2.3-17.24) |
| **1923** | White | 12.64% (11.37-13.8) | 12.52% (11.28-13.76) | -0.89% (-3.36-1.44) | 7.57% (-17.97-56.65) |
| **1924** | White | 15.09% (13.73-16.31) | 14.79% (13.49-16.04) | -1.95% (-4.41-0.55) | 7.95% (-2.16-16.95) |
| **1925** | White | 12.94% (11.75-14.03) | 12.93% (11.79-14.06) | -0.09% (-2.64-2.33) | 0.24% (-24.65-24.38) |
| **1926** | White | 15.52% (14.2-16.84) | 15.12% (13.79-16.36) | -2.61% (-4.83--0.28) | 9.41% (1.04-18.15) |
| **1927** | White | 12.03% (10.96-13.09) | 12.18% (11.11-13.23) | 1.31% (-1.38-3.88) | -17.41% (-222.33-106.51) |
| **1928** | White | 16.48% (15.18-17.76) | 16.04% (14.77-17.26) | -2.69% (-5.15--0.4) | 8.44% (1.3-16.36) |
| **1929** | White | 17.88% (16.45-19.22) | 17.38% (15.98-18.73) | -2.8% (-4.97--0.57) | 7.55% (1.42-13.83) |
| **1930** | White | 16.34% (15.05-17.59) | 15.96% (14.71-17.23) | -2.3% (-4.7-0.14) | 7.68% (-0.5-15.44) |
| **1931** | White | 13.92% (12.7-15.05) | 13.56% (12.41-14.67) | -2.57% (-5.07--0.18) | 13.18% (0.92-33.11) |
| **1932** | White | 14.43% (13.28-15.51) | 14.07% (12.92-15.13) | -2.54% (-4.93--0.14) | 11.16% (0.64-25.14) |
| **1933** | White | 12.2% (11.09-13.2) | 12% (10.9-12.96) | -1.62% (-4.01-0.86) | 18.43% (-60.12-147.31) |
| **1934** | White | 12.29% (11.1-13.43) | 12.19% (11.05-13.23) | -0.77% (-3.14-1.67) | 9.07% (-47.36-66.78) |

| | | | | | |
|---|---|---|---|---|---|
| **1935** | White | 12.36% (11.09-13.45) | 12.15% (10.95-13.21) | -1.73% (-4.62-0.78) | 16.97% (-30.41-95.16) |
| **1936** | White | 13.11% (11.85-14.21) | 12.83% (11.62-13.92) | -2.14% (-4.51-0.71) | 14.69% (-4.8-49.49) |
| **1937** | White | 12.45% (11.32-13.55) | 12.21% (11.07-13.27) | -1.91% (-4.3-0.73) | 18.06% (-17.82-96.11) |
| **1938** | White | 12.29% (11.15-13.32) | 12.15% (11.04-13.16) | -1.14% (-3.62-1.74) | 12.36% (-51.73-78.39) |
| **1939** | White | 15.41% (14.15-16.58) | 14.92% (13.69-16.04) | -3.18% (-5.29--0.66) | 11.81% (2.43-21.13) |
| **1940** | White | 13.53% (12.41-14.58) | 13.16% (12.05-14.18) | -2.76% (-5.12--0.58) | 15.66% (3.68-38) |
| **1941** | White | 10.39% (9.4-11.24) | 10.52% (9.54-11.38) | 1.24% (-1.3-3.81) | 14.87% (-45.89-201.07) |
| **1942** | White | 18.94% (17.43-20.32) | 18.17% (16.74-19.5) | -4.07% (-6.28--1.83) | 9.94% (4.59-15.62) |
| **1943** | White | 14.34% (13.14-15.5) | 14.06% (12.89-15.22) | -1.92% (-4.31-0.39) | 8.79% (-1.98-21.54) |
| **1944** | White | 14.73% (13.53-15.9) | 14.27% (13.06-15.43) | -3.15% (-5.4--0.72) | 13.17% (3.38-24.78) |
| **1945** | White | 11.21% (10.18-12.16) | 11.21% (10.18-12.16) | 0% (0-0) | 0% (0-0) |
| **1946** | White | 13.66% (12.43-14.79) | 13.51% (12.3-14.65) | -1.14% (-3.7-1.25) | 6.51% (-8.01-22.17) |
| **1947** | White | 13.46% (12.28-14.58) | 12.97% (11.79-14.05) | -3.58% (-5.85--1.15) | 21.21% (7.5-48.71) |
| **1948** | White | 15.69% (14.41-16.91) | 15.23% (13.97-16.39) | -2.9% (-5.34--0.57) | 10.34% (2.01-19.42) |
| **1949** | White | 15.39% (14.13-16.59) | 15.03% (13.76-16.18) | -2.35% (-4.82--0.01) | 8.45% (0.04-17.66) |
| **1950** | White | 16.65% (15.25-17.89) | 16.17% (14.85-17.39) | -2.84% (-5.01--0.5) | 8.64% (1.58-15.78) |
| **1951** | White | 13.53% (12.4-14.65) | 13.3% (12.19-14.36) | -1.67% (-4.07-1.1) | 9.83% (-6.68-27.68) |

| | | | | | |
|---|---|---|---|---|---|
| **1952** | White | 14.67% (13.5-15.77) | 14.42% (13.27-15.49) | -1.73% (-4.38-0.8) | 7.04% (-3.17-18.55) |
| **1953** | White | 14.73% (13.57-15.77) | 14.56% (13.43-15.62) | -1.18% (-3.51-1.22) | 4.72% (-5.2-15.03) |
| **1954** | White | 20.66% (19.24-22.05) | 19.99% (18.51-21.31) | -3.25% (-5.33--1.02) | 7.02% (2.33-11.65) |
| **1955** | White | 19.1% (17.78-20.35) | 18.72% (17.34-19.9) | -1.97% (-4.26-0.25) | 4.7% (-0.58-10.49) |
| **1956** | White | 15.68% (14.51-16.8) | 15.56% (14.36-16.61) | -0.8% (-3.12-1.49) | 2.82% (-5.77-10.89) |
| **1957** | White | 13.07% (12.03-14.01) | 13.22% (12.13-14.13) | 1.17% (-1.59-3.63) | -8.36% (-32.46-12.97) |
| **1958** | White | 15.49% (14.21-16.61) | 15.42% (14.12-16.49) | -0.48% (-3.02-2.11) | 1.62% (-8.48-11.57) |
| **1959** | White | 24.38% (22.66-25.84) | 23.79% (21.92-25.3) | -2.4% (-4.54--0.32) | 4.38% (0.58-8.52) |
| **1960** | White | 18.04% (16.58-19.37) | 17.72% (16.2-19.11) | -1.76% (-4.16-0.61) | 4.51% (-1.61-10.95) |
| **1961** | White | 16.8% (15.56-18) | 16.57% (15.28-17.85) | -1.38% (-3.76-1.06) | 4.24% (-3.26-11.37) |
| **1962** | White | 19.02% (17.58-20.38) | 18.66% (17.24-20.03) | -1.91% (-4.34-0.32) | 4.39% (-0.76-10.62) |
| **1963** | White | 15.65% (14.53-16.76) | 15.47% (14.34-16.55) | -1.15% (-3.58-1.36) | 4.31% (-4.87-13.25) |
| **1964** | White | 15.25% (14.1-16.39) | 15.16% (14.09-16.35) | -0.58% (-3.08-1.84) | 2.09% (-7.31-11.74) |
| **1965** | White | 18.58% (17.05-20.23) | 18.24% (16.75-19.85) | -1.83% (-4.43-0.49) | 4.57% (-1.25-11.07) |
| **1966** | White | 22.44% (20.21-24.92) | 21.8% (19.62-24.14) | -2.84% (-5.38--0.29) | 5.79% (0.56-10.55) |

#### Section 2.3. Physical Activity

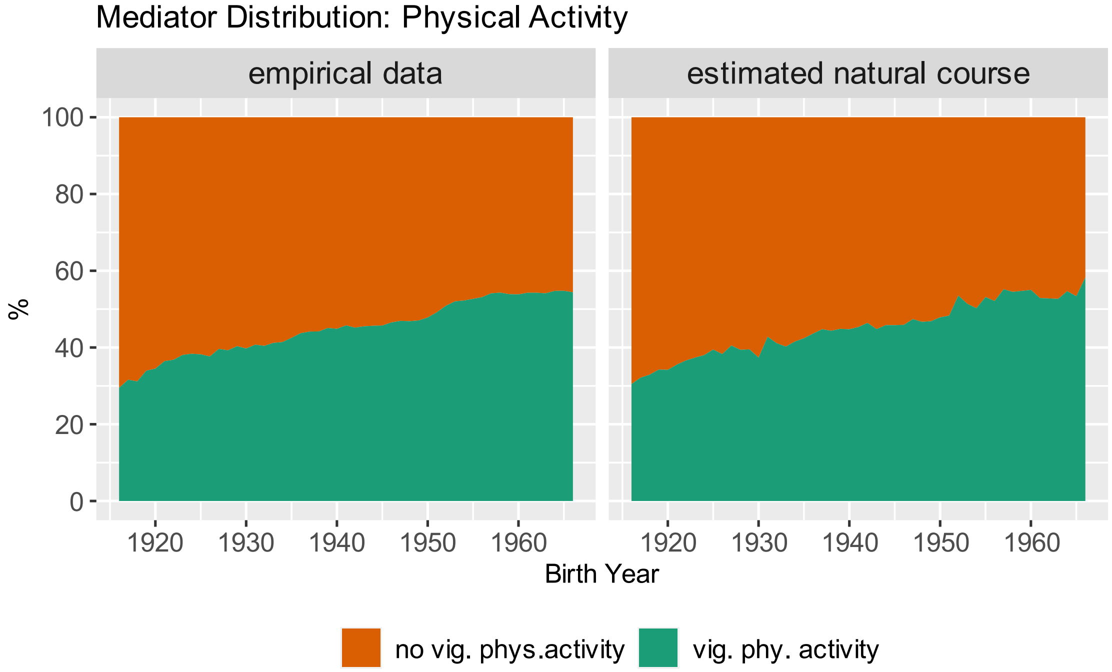

*Figure S. 12 Natural course distribution of smoking by birth cohort. "Empirical data" shows the descriptive distribution as observed in the data. "Estimated natural course" is estimated based on the mediator model without holding age and period constant*

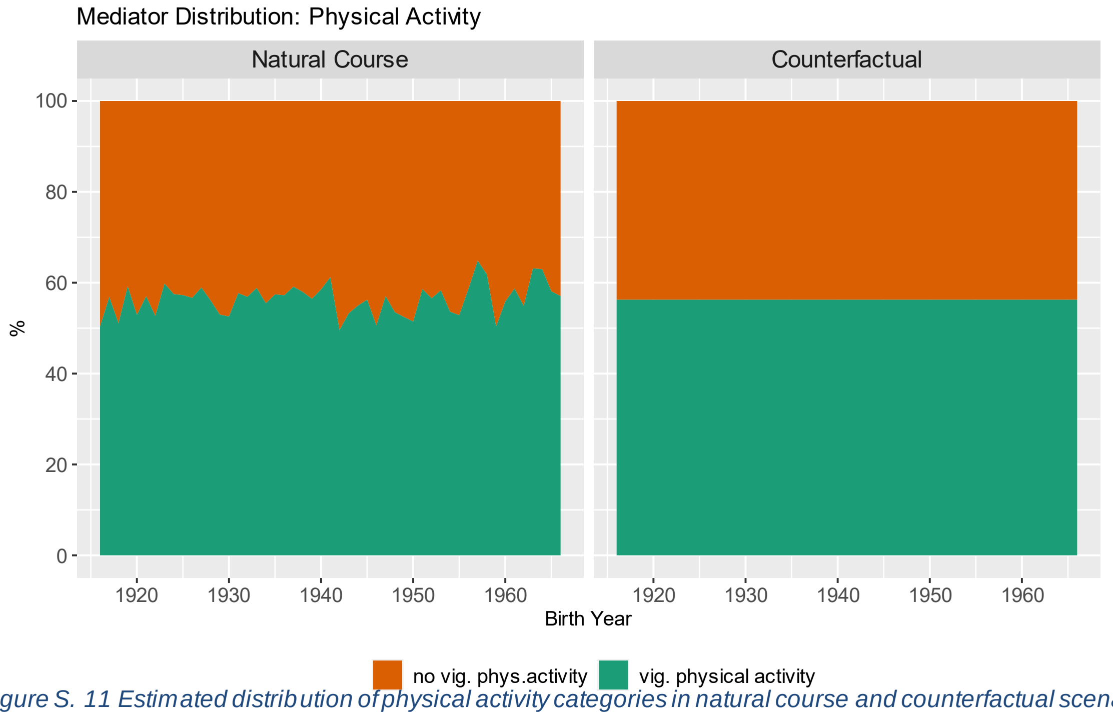

*Figure S. 11 Estimated distribution of physical activity categories in natural course and counterfactual scenario by birth year. Age and period are held constant at age 50 and 1996 respectively*

*Table S. 8 Predicted Probability of elevated depressive symptoms in natural course and counterfactual scenario, relative difference, and contribution by birth cohort. All estimates are presented with 95% confidence intervals. Positive contributions imply that the counterfactual decreases depression risk and negative contributions imply that the counterfactual increases depression risk.*

| | **Probability of elevated depressive symptoms (% (95%CI))** | | **% relative difference** | **% contribution** |
|---|---|---|---|---|
| **Birth Cohort** | Natural Course Scenario | Counterfactual Scenario | | |
| **1916** | 20.35% (18.57-22.17) | 19.56% (17.8-21.37) | -3.87% (-7.55--0.31) | 12.22% (0.97-24.93) |
| **1917** | 19.68% (18.19-21.07) | 19.41% (17.98-20.78) | -1.34% (-5.55-2.16) | 4.45% (-8.26-18.11) |
| **1918** | 22.48% (21.06-23.75) | 21.8% (20.46-23.15) | -3.01% (-6.49-0.99) | 7.89% (-2.67-16.83) |
| **1919** | 16.58% (15.47-17.66) | 16.85% (15.71-17.88) | 1.63% (-2.05-5.73) | -10.32% (-46.89-12.06) |
| **1920** | 19.51% (18.28-20.85) | 19.05% (17.85-20.23) | -2.29% (-5.68-1.38) | 8.28% (-5.02-19.39) |
| **1921** | 17.55% (16.37-18.73) | 17.46% (16.26-18.54) | -0.5% (-4.03-3.23) | 2.68% (-18.32-19.99) |
| **1922** | 20.29% (18.97-21.59) | 19.43% (18.21-20.59) | -4.22% (-7.32--1.01) | 13.43% (3.4-23.44) |
| **1923** | 15.21% (14.25-16.19) | 15.28% (14.3-16.26) | 0.54% (-3.46-4.53) | -4.83% (-141.18-49.95) |
| **1924** | 18.6% (17.49-19.71) | 18.43% (17.35-19.53) | -0.88% (-4.59-2.62) | 3.37% (-11.93-18.22) |
| **1925** | 16.26% (15.37-17.37) | 16.09% (15.08-17.14) | -1.08% (-4.64-2.92) | 7.83% (-24.19-35.81) |
| **1926** | 18.43% (17.37-19.57) | 18.33% (17.26-19.5) | -0.52% (-4.24-3.24) | 2.27% (-14.56-16.86) |
| **1927** | 14.93% (13.98-15.9) | 15.02% (14.04-15.93) | 0.56% (-3.21-4.66) | -9.22% (-234.29-93.23) |

| | | | | |
|---|---|---|---|---|
| **1928** | 18.96% (17.99-19.97) | 18.63% (17.59-19.66) | -1.71% (-5.9-2.12) | 6.24% (-8.53-22.18) |
| **1929** | 20.94% (19.67-22.14) | 20.45% (19.19-21.57) | -2.31% (-5.91-1.39) | 7.06% (-4.52-17.58) |
| **1930** | 20.22% (18.89-21.38) | 19.59% (18.36-20.67) | -3.13% (-6.57-0.32) | 10.07% (-1.09-21.35) |
| **1931** | 16.87% (15.87-17.88) | 16.77% (15.7-17.78) | -0.57% (-4.42-3.21) | 3.56% (-22.7-26.31) |
| **1932** | 18% (17.01-18.99) | 17.54% (16.5-18.57) | -2.53% (-6.12-0.95) | 11.12% (-4.58-27.99) |
| **1933** | 14.57% (13.7-15.45) | 14.71% (13.8-15.64) | 1.02% (-2.56-4.98) | -10.55% (-582.35-626.23) |
| **1934** | 16.1% (15.1-17.1) | 15.86% (14.89-16.91) | -1.49% (-5.23-2.35) | 11.4% (-24.08-39.05) |
| **1935** | 15.21% (14.27-16.18) | 15.23% (14.32-16.22) | 0.19% (-3.88-3.82) | -2.95% (-108.06-56.13) |
| **1936** | 16.32% (15.27-17.29) | 16.03% (15.09-17.06) | -1.76% (-5.58-2.38) | 13.5% (-18.13-37.5) |
| **1937** | 14.38% (13.55-15.26) | 14.54% (13.75-15.46) | 1.15% (-3.07-5.15) | -10.24% (-619.75-806.46) |
| **1938** | 14.94% (14.03-15.95) | 14.99% (14.16-15.95) | 0.33% (-3.77-4.68) | -3.44% (-229.78-163.42) |
| **1939** | 18.12% (17.15-19.25) | 17.76% (16.75-18.78) | -1.99% (-5.72-1.48) | 9.21% (-7.19-24.35) |
| **1940** | 15.37% (14.47-16.31) | 15.23% (14.29-16.18) | -0.89% (-4.66-3.19) | 10.87% (-47.41-55.48) |
| **1941** | 12.76% (11.9-13.69) | 12.96% (12.11-13.83) | 1.63% (-2.14-6) | 16.78% (-36.82-68.63) |
| **1942** | 21.73% (20.45-22.96) | 20.68% (19.41-21.84) | -4.83% (-7.98--1.81) | 13.38% (5.38-22.41) |
| **1943** | 16.95% (15.94-17.96) | 16.59% (15.53-17.59) | -2.1% (-5.67-1.4) | 11.94% (-9.4-34.18) |
| **1944** | 16.27% (15.35-17.29) | 15.98% (15.01-16.93) | -1.72% (-5.23-1.96) | 12.46% (-17.26-39.26) |

| | | | | |
|---|---|---|---|---|
| **1945** | 14% (13.07-14.99) | 14% (13.07-14.99) | 0% (0-0) | 0% (0-0) |
| **1946** | 17.69% (16.56-18.92) | 16.93% (15.74-18.2) | -4.28% (-7.95--0.67) | 20.39% (3.49-36.74) |
| **1947** | 16.8% (15.71-17.91) | 16.65% (15.49-17.76) | -0.83% (-4.56-2.89) | 5.31% (-20.01-26.65) |
| **1948** | 18.63% (17.55-19.78) | 17.99% (16.96-19.09) | -3.43% (-6.61--0.17) | 14.16% (0.74-27.46) |
| **1949** | 19.16% (18.03-20.3) | 18.23% (17.17-19.31) | -4.81% (-8.24--1.62) | 17.7% (6.1-30.7) |
| **1950** | 20.75% (19.64-21.89) | 19.56% (18.35-20.8) | -5.75% (-9.03--2.52) | 17.44% (8.03-28.36) |
| **1951** | 16.62% (15.64-17.64) | 16.41% (15.34-17.4) | -1.28% (-4.88-2.67) | 7.82% (-21.01-33.13) |
| **1952** | 17.86% (16.89-18.93) | 17.69% (16.68-18.73) | -0.95% (-4.47-3.08) | 4.77% (-16.61-20.73) |
| **1953** | 18.43% (17.44-19.4) | 18.44% (17.44-19.49) | 0.08% (-3.66-3.91) | -0.06% (-16.89-14.54) |
| **1954** | 24.26% (23.1-25.58) | 23.59% (22.4-24.83) | -2.73% (-5.92-0.51) | 6.69% (-1.25-13.75) |
| **1955** | 22.92% (21.92-24.19) | 22.29% (21.2-23.45) | -2.72% (-6.23-0.69) | 7.11% (-1.82-15.71) |
| **1956** | 18.98% (18-20.03) | 19.15% (18.16-20.16) | 0.91% (-2.81-4.66) | -3.76% (-19.63-10.48) |
| **1957** | 14.61% (13.77-15.54) | 15.18% (14.36-16.05) | 3.95% (-0.06-8.36) | -75.37% (-990.94-1235.25) |
| **1958** | 17.85% (16.99-18.81) | 18.27% (17.22-19.34) | 2.34% (-1.7-6.46) | -10.35% (-33.57-7.45) |
| **1959** | 28.22% (26.91-29.55) | 27.17% (25.79-28.48) | -3.71% (-6.84--0.67) | 7.21% (1.36-13.61) |
| **1960** | 21.57% (20.37-22.82) | 21.26% (20.09-22.43) | -1.43% (-4.7-2.11) | 4.54% (-6.26-12.97) |
| **1961** | 19.81% (18.81-20.92) | 19.45% (18.41-20.51) | -1.8% (-5.59-1.8) | 6.15% (-6.92-18.49) |

| | | | | |
|---|---|---|---|---|
| **1962** | 22.08% (20.98-23.23) | 21.77% (20.66-22.88) | -1.38% (-5.01-2.18) | 3.67% (-6.3-13.73) |
| **1963** | 17.1% (16.37-17.99) | 17.77% (16.94-18.68) | 3.98% (-0.32-7.76) | -22.63% (-48.46-1.59) |
| **1964** | 16.8% (16.01-17.65) | 17.3% (16.5-18.22) | 2.98% (-0.72-6.85) | -17.63% (-47.93-3.89) |
| **1965** | 20.97% (19.93-22.23) | 20.99% (19.85-22.3) | 0.14% (-3.48-4.24) | -0.11% (-13.13-10.13) |
| **1966** | 22.67% (21.12-24.34) | 22.4% (20.91-24.09) | -1.16% (-5.16-2.64) | 2.98% (-6.87-12.97) |

*Table S. 9 Predicted Probability of elevated depressive symptoms in natural course and counterfactual scenario, relative difference, and contribution by birth cohort and* ***stratified by sex.*** *All estimates are presented with 95% confidence intervals. Positive contributions imply that the counterfactual decreases depression risk and negative contributions imply that the counterfactual increases depression risk.*

| | | **Probability of elevated depressive symptoms (% (95%CI))** | | **% relative difference** | **% contribution** |
|---|---|---|---|---|---|
| **Birth Cohort** | sex | Natural Course Scenario | Counterfactual Scenario | | |
| **1916** | female | 21.58% (19.07-24.13) | 20.92% (18.38-23.35) | -3.02% (-6.47-0.68) | 10.11% (-2.59-22.57) |
| **1917** | female | 23.84% (21.62-25.99) | 23.2% (21-25.26) | -2.68% (-6.06-0.73) | 7.36% (-2.04-16.21) |
| **1918** | female | 25.31% (23.2-27.29) | 24.6% (22.5-26.54) | -2.78% (-5.55--0.06) | 6.85% (0.14-14.2) |
| **1919** | female | 20.1% (18.38-21.74) | 20.16% (18.36-21.94) | 0.34% (-3.29-3.69) | -1.27% (-17.26-11.96) |
| **1920** | female | 20.78% (18.82-22.72) | 20.61% (18.6-22.36) | -0.83% (-4.48-2.64) | 3.1% (-10.71-15.84) |
| **1921** | female | 20.38% (18.45-22.23) | 20.4% (18.42-22.22) | 0.13% (-2.99-3.74) | -0.57% (-17.91-12.6) |
| **1922** | female | 20.97% (18.8-22.84) | 20.81% (18.75-22.62) | -0.75% (-3.94-2.49) | 2.9% (-10.03-13.84) |

| | | | | | |
|---|---|---|---|---|---|
| **1923** | female | 16.61% (14.94-18.06) | 17.12% (15.38-18.57) | 3.07% (-0.52-6.89) | -31.01% (-254.92-98.92) |
| **1924** | female | 21.18% (19.46-22.79) | 21.15% (19.32-22.74) | -0.12% (-3.25-3.19) | 0.16% (-11.26-11.81) |
| **1925** | female | 17.95% (16.42-19.42) | 18.15% (16.54-19.65) | 1.11% (-2.15-4.48) | -7.05% (-38.43-12.91) |
| **1926** | female | 22.44% (20.83-23.99) | 22.21% (20.49-23.77) | -1% (-4.11-2.22) | 2.91% (-7.25-12.73) |
| **1927** | female | 15.87% (14.5-17.16) | 16.29% (14.87-17.53) | 2.63% (-0.98-6.44) | -36.76% (-1020.85-456.2) |
| **1928** | female | 21.69% (20.15-23.26) | 21.48% (19.83-22.98) | -0.97% (-4.05-2.31) | 3.38% (-8.18-13.37) |
| **1929** | female | 22.76% (21.03-24.48) | 22.48% (20.84-24.29) | -1.19% (-4.58-2.09) | 3.57% (-6.71-13.88) |
| **1930** | female | 21.69% (20.09-23.4) | 21.33% (19.71-23.07) | -1.67% (-4.68-1.41) | 5.41% (-4.91-15.22) |
| **1931** | female | 18.88% (17.44-20.39) | 18.79% (17.35-20.37) | -0.47% (-4.07-2.97) | 1.95% (-17.97-21.27) |
| **1932** | female | 19.13% (17.81-20.71) | 18.86% (17.36-20.39) | -1.42% (-4.81-1.82) | 6.99% (-10.22-24.25) |
| **1933** | female | 15.94% (14.65-17.31) | 15.99% (14.76-17.24) | 0.34% (-3.82-3.95) | -3.03% (-272.18-247.55) |
| **1934** | female | 16.22% (14.89-17.5) | 16.2% (14.91-17.42) | -0.11% (-3.44-3.65) | 3.37% (-164.45-144.59) |
| **1935** | female | 15.94% (14.61-17.21) | 15.99% (14.78-17.18) | 0.35% (-3-3.82) | -0.24% (-234.79-245.73) |
| **1936** | female | 16.81% (15.54-18.06) | 16.68% (15.36-17.93) | -0.77% (-3.99-2.96) | 8.58% (-52.93-59.18) |
| **1937** | female | 15.72% (14.47-16.91) | 15.77% (14.48-16.95) | 0.33% (-3.08-3.9) | -0.02% (-553.52-427.67) |
| **1938** | female | 15.57% (14.4-16.82) | 15.71% (14.43-16.9) | 0.92% (-2.88-4.85) | -4.28% (-549.28-486.86) |
| **1939** | female | 19.85% (18.46-21.16) | 19.43% (18.04-20.77) | -2.09% (-5.29-0.98) | 8.82% (-4.39-22.19) |

| | | | | | |
|---|---|---|---|---|---|
| **1940** | female | 16.83% (15.48-18.18) | 16.63% (15.3-17.98) | -1.17% (-4.42-2.38) | 12.12% (-35.17-61.44) |
| **1941** | female | 13.56% (12.49-14.69) | 13.92% (12.73-15.12) | 2.63% (-0.89-6.47) | 22.39% (-9.87-86.85) |
| **1942** | female | 22.05% (20.52-23.64) | 21.17% (19.53-22.86) | -3.99% (-7.04--0.73) | 12.55% (2.35-22.43) |
| **1943** | female | 17.6% (16.18-18.93) | 17.36% (15.89-18.75) | -1.37% (-4.78-2.18) | 10.64% (-17.78-36.6) |
| **1944** | female | 18.27% (16.77-19.6) | 17.96% (16.57-19.2) | -1.67% (-5.2-1.73) | 10.19% (-10.61-30.12) |
| **1945** | female | 15.15% (13.9-16.34) | 15.15% (13.9-16.34) | 0% (0-0) | 0% (0-0) |
| **1946** | female | 17.8% (16.3-19.07) | 17.36% (15.92-18.61) | -2.49% (-5.9-0.79) | 16.65% (-7.78-45.46) |
| **1947** | female | 20.11% (18.62-21.41) | 19.44% (17.96-20.68) | -3.32% (-6.59--0.28) | 13.06% (1.37-27.75) |
| **1948** | female | 19.43% (17.93-20.69) | 18.74% (17.37-20.05) | -3.51% (-6.76--0.56) | 15.87% (2.87-31.63) |
| **1949** | female | 19.35% (17.97-20.68) | 18.61% (17.28-19.83) | -3.8% (-6.82--0.37) | 17.59% (1.63-33.3) |
| **1950** | female | 19.78% (18.43-21.1) | 18.93% (17.6-20.22) | -4.31% (-7.76--1.28) | 18.34% (5.63-32.61) |
| **1951** | female | 17.69% (16.47-18.95) | 17.31% (16.1-18.56) | -2.13% (-5.89-1.23) | 15.23% (-10.14-41.53) |
| **1952** | female | 18.71% (17.39-19.93) | 18.34% (17.1-19.62) | -1.99% (-5.08-1.39) | 10.72% (-7.89-27.73) |
| **1953** | female | 20.09% (18.8-21.33) | 19.79% (18.49-21.06) | -1.51% (-4.89-1.55) | 5.77% (-7.02-19.63) |
| **1954** | female | 26.6% (24.97-28.09) | 25.48% (23.86-27.09) | -4.2% (-7.06--1.34) | 9.7% (3.17-16.37) |
| **1955** | female | 24.34% (22.88-25.78) | 23.42% (21.8-24.88) | -3.79% (-6.88--0.73) | 10.27% (1.92-18.04) |
| **1956** | female | 21.75% (20.24-23.29) | 21.35% (19.81-22.73) | -1.83% (-4.93-1.66) | 6.2% (-5.91-16.02) |

| | | | | | |
|---|---|---|---|---|---|
| **1957** | female | 16.6% (15.42-17.71) | 17.02% (15.67-18.18) | 2.52% (-1.04-6.33) | -27.75% (-140.96-19.69) |
| **1958** | female | 21.68% (20.23-23.02) | 21.43% (19.95-22.86) | -1.13% (-4.32-1.88) | 4.11% (-6.24-14.39) |
| **1959** | female | 29.75% (27.9-31.48) | 28.38% (26.58-30.06) | -4.62% (-7.28--2) | 9.32% (4.21-14.71) |
| **1960** | female | 23.28% (21.69-24.84) | 22.62% (20.95-24.14) | -2.82% (-5.97-0.35) | 8.2% (-0.98-17.33) |
| **1961** | female | 21.59% (20.01-23.07) | 21.05% (19.55-22.47) | -2.48% (-5.5-0.8) | 8.47% (-2.83-18.54) |
| **1962** | female | 24.28% (22.78-25.78) | 23.41% (21.91-24.83) | -3.57% (-6.31--0.5) | 9.73% (1.35-16.6) |
| **1963** | female | 20.83% (19.59-22.12) | 20.63% (19.46-21.88) | -0.93% (-4.59-2.42) | 3.73% (-9.48-15.73) |
| **1964** | female | 19.71% (18.43-21.07) | 19.73% (18.45-21.1) | 0.11% (-3.52-3.7) | -0.47% (-16.96-15.24) |
| **1965** | female | 24% (22.01-25.79) | 23.43% (21.62-25.27) | -2.37% (-5.44-0.88) | 6.52% (-2.42-14.92) |
| **1966** | female | 25.67% (23.38-27.97) | 24.99% (22.7-27.34) | -2.62% (-5.55-0.66) | 6.46% (-1.61-13.91) |
| **1916** | male | 21.82% (19.04-24.56) | 20.73% (17.8-23.51) | -5.03% (-9.07--0.72) | 13.22% (2.14-24.8) |
| **1917** | male | 22.9% (20.45-25.52) | 21.71% (19.32-24.1) | -5.19% (-9--1.82) | 12.27% (4.63-22.53) |
| **1918** | male | 24.42% (21.94-26.75) | 23.21% (20.85-25.58) | -4.94% (-8.5--1.16) | 11.17% (2.51-18.99) |
| **1919** | male | 19.25% (17.19-21.06) | 18.64% (16.58-20.42) | -3.15% (-6.95-0.65) | 10.84% (-2.83-23.86) |
| **1920** | male | 19.76% (17.74-21.62) | 19.04% (17.04-20.98) | -3.67% (-7.3-0.38) | 11.56% (-1.13-24.35) |
| **1921** | male | 18.41% (16.52-20.23) | 17.8% (15.73-19.75) | -3.31% (-7.27-0.52) | 12.55% (-2.08-28.28) |
| **1922** | male | 18.55% (16.56-20.35) | 17.89% (15.9-19.79) | -3.55% (-7.54-0.75) | 13.19% (-2.67-28.73) |

| | | | | | |
|---|---|---|---|---|---|
| **1923** | male | 14.6% (13.03-16.25) | 14.56% (12.95-16.14) | -0.24% (-4.54-3.97) | 5.29% (-134.28-121.95) |
| **1924** | male | 17.41% (15.53-19.16) | 17.01% (15.17-18.59) | -2.26% (-5.98-2.02) | 10.31% (-9.97-26.88) |
| **1925** | male | 14.94% (13.43-16.51) | 14.67% (13.14-16.17) | -1.75% (-5.42-2.54) | 16.91% (-82.52-119.37) |
| **1926** | male | 18.27% (16.56-19.89) | 17.67% (15.88-19.35) | -3.29% (-7.54-0.96) | 12.76% (-3.7-29.14) |
| **1927** | male | 13.67% (12.23-15.21) | 13.73% (12.29-15.09) | 0.48% (-3.82-5.03) | 6.05% (-772.97-384.6) |
| **1928** | male | 18.28% (16.47-19.98) | 17.64% (15.88-19.16) | -3.45% (-7.08-0.62) | 13.69% (-2.54-28.49) |
| **1929** | male | 19.68% (17.78-21.45) | 19.16% (17.22-20.73) | -2.67% (-6.36-1.23) | 8.8% (-4.26-20.12) |
| **1930** | male | 19.6% (17.76-21.29) | 19% (17.05-20.64) | -3.04% (-6.81-1.31) | 9.35% (-4.12-23.07) |
| **1931** | male | 17.84% (16.14-19.53) | 17.51% (15.72-18.98) | -1.86% (-5.57-2.07) | 8.31% (-8.78-24.77) |
| **1932** | male | 17.69% (15.96-19.26) | 17.34% (15.58-18.84) | -1.92% (-5.99-2.01) | 7.71% (-9.75-26.39) |
| **1933** | male | 15.52% (13.96-17.05) | 15.45% (13.84-16.94) | -0.39% (-4.45-3.65) | 3.75% (-40.96-40.58) |
| **1934** | male | 16.35% (14.71-17.95) | 16.38% (14.73-17.95) | 0.2% (-3.84-4.46) | -0.73% (-32.81-24.22) |
| **1935** | male | 15.95% (14.39-17.48) | 16.03% (14.4-17.52) | 0.52% (-4.16-4.92) | -4.18% (-43.65-26.04) |
| **1936** | male | 16.5% (14.97-18) | 16.52% (14.86-18.13) | 0.12% (-4.09-4.26) | -0.75% (-27.6-22.42) |
| **1937** | male | 15.2% (13.78-16.6) | 15.42% (13.86-16.89) | 1.47% (-2.73-6.52) | -11.97% (-113.92-28.58) |
| **1938** | male | 15.21% (13.72-16.62) | 15.43% (13.9-16.96) | 1.5% (-2.76-5.98) | -12.65% (-93.84-32.51) |
| **1939** | male | 18.49% (16.83-20.03) | 18.31% (16.65-19.81) | -0.98% (-5.02-3.3) | 3.25% (-12.88-18.6) |

| | | | | | |
|---|---|---|---|---|---|
| **1940** | male | 16.04% (14.48-17.61) | 16.11% (14.56-17.54) | 0.51% (-4.02-4.95) | -4.29% (-39.31-25.94) |
| **1941** | male | 12.91% (11.54-14.11) | 13.31% (11.93-14.55) | 3.15% (-1.41-8.17) | 41.95% (-753.42-863.49) |
| **1942** | male | 19.73% (18.1-21.4) | 19.34% (17.57-21.06) | -1.96% (-5.52-2.07) | 6.21% (-6.68-17.54) |
| **1943** | male | 15.91% (14.4-17.37) | 15.85% (14.37-17.29) | -0.32% (-4.25-4.18) | 2.35% (-37.99-30.69) |
| **1944** | male | 15.67% (14.22-17.15) | 15.41% (13.98-16.85) | -1.63% (-5.96-2.45) | 11.02% (-21.51-53.34) |
| **1945** | male | 13.44% (12.13-14.87) | 13.44% (12.13-14.87) | 0% (0-0) | 0% (0-0) |
| **1946** | male | 15.42% (13.94-17.07) | 15.17% (13.66-16.72) | -1.65% (-5.93-2.34) | 12.43% (-24.9-60.35) |
| **1947** | male | 17.73% (16.1-19.42) | 17.26% (15.67-18.88) | -2.62% (-6.6-1.08) | 10.66% (-4.74-28.35) |
| **1948** | male | 17.98% (16.48-19.63) | 17.56% (16.04-19.25) | -2.31% (-6.15-1.96) | 9.53% (-8.22-24.03) |
| **1949** | male | 19.2% (17.55-20.8) | 18.86% (17.18-20.54) | -1.78% (-5.93-2.11) | 5.86% (-6.89-20.76) |
| **1950** | male | 20.49% (18.73-22.25) | 20.17% (18.41-21.92) | -1.57% (-5.25-2.37) | 5.07% (-7.39-15.18) |
| **1951** | male | 19.1% (17.26-20.76) | 19.19% (17.45-20.82) | 0.46% (-3.7-4.5) | -1.5% (-17.57-12.32) |
| **1952** | male | 19.51% (17.72-21.15) | 19.64% (17.89-21.34) | 0.72% (-3.28-4.77) | -2.52% (-16.26-10.92) |
| **1953** | male | 20.15% (18.28-21.76) | 20.43% (18.7-22.1) | 1.4% (-2.58-5.42) | -3.98% (-17.2-7.28) |
| **1954** | male | 25.48% (23.52-27.54) | 25.36% (23.4-27.26) | -0.46% (-3.97-3) | 0.74% (-6.37-8.45) |
| **1955** | male | 22.45% (20.71-24.27) | 22.52% (20.75-24.24) | 0.3% (-3.5-3.99) | -0.58% (-10.54-8.38) |
| **1956** | male | 19.64% (18.03-21.27) | 20.27% (18.58-21.88) | 3.23% (-0.95-7.3) | -10.02% (-24.47-2.86) |

| | | | | | |
|---|---|---|---|---|---|
| **1957** | male | 14.87% (13.48-16.25) | 15.84% (14.43-17.26) | 6.59% (1.87-11.6) | -66.12% (-516.4--4.87) |
| **1958** | male | 18.41% (16.81-20.08) | 19.27% (17.66-20.83) | 4.66% (0.13-8.86) | -17.44% (-37.07--0.43) |
| **1959** | male | 24.51% (22.58-26.53) | 24.95% (22.95-26.82) | 1.81% (-1.62-5.68) | -3.9% (-12.89-3.61) |
| **1960** | male | 20.2% (18.51-21.88) | 21.03% (19.2-22.74) | 4.13% (-0.03-8.44) | -12.05% (-28.32-0.08) |
| **1961** | male | 18.2% (16.64-19.84) | 19.1% (17.4-20.75) | 4.97% (0.44-9.3) | -19.52% (-40--1.59) |
| **1962** | male | 20.48% (18.96-22.11) | 21.36% (19.68-22.96) | 4.33% (0.02-8.75) | -12.63% (-27--0.05) |
| **1963** | male | 17.17% (15.95-18.3) | 18.44% (17.08-19.81) | 7.45% (2.83-12.34) | -34.61% (-67.91--11.61) |
| **1964** | male | 16.3% (15.1-17.6) | 17.54% (16.18-18.92) | 7.61% (2.86-12.5) | -43.01% (-93.44--14.38) |
| **1965** | male | 18.78% (17.11-20.41) | 19.73% (18.08-21.45) | 5.09% (0.74-9.99) | -17.92% (-39.41--2.4) |
| **1966** | male | 19.8% (17.75-22.06) | 20.67% (18.64-22.92) | 4.38% (0.12-8.95) | -13.73% (-30.06--0.33) |

*Table S. 10 Predicted Probability of elevated depressive symptoms in natural course and counterfactual scenario, relative difference, and contribution by birth cohort and* ***stratified by race/ethnicity****. All estimates are presented with 95% confidence intervals. Positive contributions imply that the counterfactual decreases depression risk and negative contributions imply that the counterfactual increases depression risk*

| | | **Probability of elevated depressive symptoms (% (95%CI))** | | **% relative difference** | **% contribution** |
|---|---|---|---|---|---|
| **Birth Cohort** | race/ethnicity | Natural Course Scenario | Counterfactual Scenario | | |
| **1916** | Black | 34.26% (31.07-37.53) | 31.95% (29.13-35.09) | -6.74% (-9.11--4.3) | 16.66% (10.14-22.8) |
| **1917** | Black | 31.54% (29.1-34.02) | 30.27% (28-32.85) | -4.03% (-6.32--1.59) | 11.39% (4.26-18.35) |

| | | | | | |
|---|---|---|---|---|---|
| **1918** | Black | 34.09% (31.79-36.37) | 32.4% (30.23-34.79) | -4.95% (-7.34--2.67) | 12.22% (6.91-17.84) |
| **1919** | Black | 25.95% (24.14-27.83) | 25.55% (23.8-27.4) | -1.56% (-3.99-1.05) | 7.15% (-4.92-19.44) |
| **1920** | Black | 28.69% (26.86-30.75) | 27.78% (25.97-29.75) | -3.17% (-5.6--0.75) | 10.79% (2.66-19.6) |
| **1921** | Black | 25.98% (24.14-28.11) | 25.41% (23.6-27.3) | -2.19% (-4.55-0.61) | 10.25% (-2.85-21.6) |
| **1922** | Black | 28.17% (26.36-30.16) | 26.91% (25.08-28.81) | -4.47% (-6.64--1.84) | 16.14% (6.64-24.72) |
| **1923** | Black | 23.14% (21.46-24.8) | 22.79% (21.22-24.41) | -1.5% (-4.07-1.37) | 12.69% (-13.25-39.22) |
| **1924** | Black | 24.85% (23.18-26.5) | 24.42% (22.84-26.06) | -1.72% (-3.98-0.79) | 9.52% (-4.45-23.42) |
| **1925** | Black | 21.31% (19.82-22.88) | 21.09% (19.67-22.59) | -1.03% (-3.66-1.47) | 19.65% (-163.86-332.31) |
| **1926** | Black | 23.71% (22.12-25.36) | 23.6% (22.08-25.18) | -0.47% (-2.8-2.39) | 3.6% (-21.47-21.11) |
| **1927** | Black | 19.15% (17.81-20.58) | 19.31% (17.97-20.76) | 0.81% (-1.9-3.73) | 11.74% (-92.32-146.72) |
| **1928** | Black | 24.23% (22.75-25.92) | 24.05% (22.59-25.66) | -0.73% (-3.03-1.74) | 4.67% (-12.01-20.02) |
| **1929** | Black | 26.43% (24.74-28.22) | 25.79% (24.23-27.46) | -2.38% (-4.57--0.01) | 10.23% (0.04-19.97) |
| **1930** | Black | 24.37% (22.8-26.03) | 23.93% (22.44-25.52) | -1.8% (-4.33-0.89) | 10.78% (-6-27.12) |
| **1931** | Black | 21.45% (19.99-23.09) | 21.65% (20.19-23.19) | 0.97% (-1.63-3.75) | -12.74% (-437.61-360.57) |
| **1932** | Black | 22.07% (20.69-23.62) | 21.97% (20.57-23.5) | -0.41% (-2.84-2.34) | 6.59% (-42.83-61.43) |
| **1933** | Black | 21% (19.49-22.49) | 21.27% (19.75-22.74) | 1.27% (-1.28-4.28) | -17.28% (-509.06-413.67) |
| **1934** | Black | 19.78% (18.37-21.21) | 19.88% (18.49-21.25) | 0.55% (-2.06-3.56) | 12.09% (-269.44-378.93) |

| | | | | | |
|---|---|---|---|---|---|
| **1935** | Black | 19.96% (18.52-21.42) | 20.21% (18.76-21.68) | 1.29% (-1.54-4.18) | 18.26% (-398.77-494.07) |
| **1936** | Black | 21.23% (19.77-22.77) | 21.11% (19.58-22.6) | -0.54% (-3.23-2.38) | 10.3% (-357.65-193.49) |
| **1937** | Black | 19.49% (18.09-20.95) | 19.47% (18.15-20.91) | -0.08% (-3.06-2.85) | 0.4% (-242.9-235.94) |
| **1938** | Black | 18.26% (16.94-19.75) | 18.1% (16.88-19.43) | -0.86% (-3.6-2.34) | -7.98% (-70.56-20.88) |
| **1939** | Black | 20.07% (18.77-21.51) | 19.69% (18.34-21.21) | -1.91% (-4.56-0.8) | -20.16% (-462.08-638.74) |
| **1940** | Black | 18.27% (16.95-19.76) | 17.83% (16.54-19.27) | -2.37% (-5.02-0.63) | -21.38% (-122.23-5.62) |
| **1941** | Black | 14.63% (13.5-15.94) | 14.63% (13.48-15.87) | 0% (-3.07-3.07) | 0.06% (-8.66-8.24) |
| **1942** | Black | 25.03% (23.4-26.73) | 23.73% (22.16-25.41) | -5.17% (-7.39--2.83) | 27.64% (13.95-43.76) |
| **1943** | Black | 21.1% (19.56-22.81) | 20.4% (18.96-21.97) | -3.3% (-5.77--0.67) | 60.49% (-639.84-628.98) |
| **1944** | Black | 23.84% (22.23-25.59) | 23.11% (21.53-24.84) | -3.03% (-5.44--0.54) | 20.39% (3.74-41.27) |
| **1945** | Black | 20.29% (18.89-21.86) | 20.29% (18.89-21.86) | 0% (0-0) | 0% (0-0) |
| **1946** | Black | 26.25% (24.55-28.11) | 25.42% (23.79-27.23) | -3.16% (-5.59--0.56) | 13.74% (2.56-26.55) |
| **1947** | Black | 24.35% (22.73-26.01) | 24.51% (22.95-26.18) | 0.69% (-1.98-3.47) | -3.72% (-25.14-11.21) |
| **1948** | Black | 27.68% (26.08-29.36) | 27.19% (25.56-28.83) | -1.76% (-3.89-0.67) | 6.61% (-2.57-14.38) |
| **1949** | Black | 26.81% (25.22-28.48) | 26.23% (24.61-27.89) | -2.16% (-4.46-0.37) | 8.91% (-1.53-18.77) |
| **1950** | Black | 27.11% (25.49-28.79) | 26.42% (24.78-28.18) | -2.56% (-4.76--0.05) | 10.53% (0.2-19.86) |
| **1951** | Black | 22.34% (20.95-23.81) | 22.51% (21.1-24.08) | 0.78% (-1.87-3.67) | -8.48% (-65.33-22.01) |

| | | | | | |
|---|---|---|---|---|---|
| **1952** | Black | 24.4% (22.85-26.06) | 24.38% (22.87-25.98) | -0.08% (-2.47-2.42) | 0.73% (-16.25-15.4) |
| **1953** | Black | 24.57% (23.16-26.17) | 24.56% (23.05-26.15) | -0.01% (-2.66-2.55) | 0.28% (-16.53-14.69) |
| **1954** | Black | 32.52% (30.75-34.43) | 31.95% (30.16-33.84) | -1.76% (-4.13-0.54) | 4.6% (-1.5-10.57) |
| **1955** | Black | 31.48% (29.78-33.25) | 30.65% (28.93-32.38) | -2.61% (-4.95--0.41) | 7.55% (1.14-13.55) |
| **1956** | Black | 25.65% (24.16-27.4) | 25.58% (24.05-27.19) | -0.28% (-2.75-2.4) | 1.56% (-12.22-13.26) |
| **1957** | Black | 21.22% (19.83-22.51) | 21.56% (20.14-22.98) | 1.61% (-0.94-4.41) | -29.36% (-438.86-207.66) |
| **1958** | Black | 24.61% (23.09-26.15) | 24.68% (23.15-26.19) | 0.29% (-2.23-3.02) | -1.38% (-19.38-12.49) |
| **1959** | Black | 32.91% (31.14-34.67) | 31.75% (29.92-33.53) | -3.53% (-5.71--1.48) | 9.09% (4.05-14.85) |
| **1960** | Black | 24.58% (23.07-26.24) | 24.27% (22.69-25.87) | -1.22% (-3.56-1.51) | 7% (-9.46-21.18) |
| **1961** | Black | 21.98% (20.59-23.41) | 21.68% (20.32-23.08) | -1.39% (-4.12-1.49) | 18.87% (-29.34-67.73) |
| **1962** | Black | 23.75% (22.34-25.3) | 23.52% (22.17-24.99) | -0.98% (-3.77-1.72) | 6.73% (-14.18-24.78) |
| **1963** | Black | 19.31% (18.19-20.54) | 19.77% (18.67-21.05) | 2.37% (-0.38-5.23) | 41.19% (-170.66-274.46) |
| **1964** | Black | 18.05% (16.79-19.28) | 18.54% (17.42-19.84) | 2.73% (-0.06-5.72) | 21.36% (-0.42-57.82) |
| **1965** | Black | 22.31% (20.75-24.09) | 22.39% (20.8-24.15) | 0.35% (-2.23-3.16) | -3.11% (-54.82-28.27) |
| **1966** | Black | 25.57% (23.3-27.69) | 25.3% (23.12-27.56) | -1.04% (-3.69-1.76) | 5.3% (-9.76-18.89) |
| **1916** | Hispanic | 12.33% (10.38-14.48) | 9.55% (7.89-11.33) | -22.57% (-25.78--18.92) | -31.44% (-51.89--21.04) |
| **1917** | Hispanic | 15.33% (13.38-17.38) | 12.6% (10.89-14.39) | -17.84% (-21.48--13.91) | -47.15% (-88.81--29.73) |

| | | | | | |
|---|---|---|---|---|---|
| **1918** | Hispanic | 23.16% (20.99-25.45) | 19.25% (17.31-21.39) | -16.87% (-19.99--13.5) | 178.53% (-331.51-924.33) |
| **1919** | Hispanic | 22.36% (20.55-24.33) | 19.68% (18.03-21.41) | -11.97% (-15.29--8.75) | 176.54% (-1216.84-1829.7) |
| **1920** | Hispanic | 29.51% (27.43-31.49) | 25.69% (23.84-27.67) | -12.92% (-15.75--9.98) | 45.31% (33.68-58.68) |
| **1921** | Hispanic | 30.44% (28.51-32.62) | 27.56% (25.61-29.65) | -9.43% (-12.16--6.46) | 30.77% (20.01-40.98) |
| **1922** | Hispanic | 33.57% (31.29-35.59) | 29.93% (27.94-32.11) | -10.85% (-13.46--8.12) | 29.26% (21.39-37.02) |
| **1923** | Hispanic | 26.07% (24.29-27.87) | 24.06% (22.48-25.93) | -7.72% (-10.92--4.58) | 40.42% (22.87-62.16) |
| **1924** | Hispanic | 25.7% (24.13-27.26) | 23.86% (22.34-25.61) | -7.15% (-10.25--3.87) | 39.45% (21.69-64.11) |
| **1925** | Hispanic | 21.74% (20.29-23.24) | 20.27% (18.88-21.86) | -6.76% (-9.98--3.12) | 145.39% (-1878.13-2076.91) |
| **1926** | Hispanic | 22.27% (20.77-23.62) | 21.32% (19.94-22.89) | -4.24% (-7.5--0.73) | 75.73% (-267.31-463.39) |
| **1927** | Hispanic | 17.63% (16.45-18.87) | 16.38% (15.17-17.75) | -7.04% (-11.22--3.49) | -35.69% (-74.45--15.04) |
| **1928** | Hispanic | 21.33% (19.94-22.78) | 19.7% (18.37-21.16) | -7.65% (-11.06--4.27) | 147.49% (-3950.22-3103.13) |
| **1929** | Hispanic | 25.17% (23.52-26.86) | 22.44% (20.83-24.14) | -10.84% (-14.02--7.79) | 66.48% (43.45-108.67) |
| **1930** | Hispanic | 24.17% (22.48-25.84) | 21.66% (20.04-23.39) | -10.4% (-13.73--7.23) | 81.97% (48.24-165.61) |
| **1931** | Hispanic | 20.53% (19.07-22.22) | 18.7% (17.26-20.18) | -8.9% (-12.44--5.7) | -160.46% (-1895.21-1865.88) |
| **1932** | Hispanic | 21.97% (20.42-23.51) | 19.93% (18.54-21.48) | -9.26% (-12.65--5.71) | 181.8% (-1565.97-1849.38) |
| **1933** | Hispanic | 20.65% (19.23-22.22) | 19.44% (18.08-20.95) | -5.88% (-9.56--2.12) | -92.76% (-2468.66-2884.05) |
| **1934** | Hispanic | 23.78% (22.16-25.51) | 22.75% (21.17-24.42) | -4.32% (-7.73--0.84) | 37.51% (7.69-91) |

| | | | | | |
|---|---|---|---|---|---|
| **1935** | Hispanic | 22.64% (21.09-24.24) | 22.19% (20.7-23.8) | -1.96% (-5.42-1.45) | 27.38% (-60.01-140.57) |
| **1936** | Hispanic | 22.25% (20.79-23.81) | 21.65% (20.2-23.34) | -2.69% (-6.43-0.62) | 47.58% (-141.26-371.04) |
| **1937** | Hispanic | 21.77% (20.29-23.4) | 21.57% (20.18-23.2) | -0.91% (-4.52-2.89) | 20.56% (-442.25-406.7) |
| **1938** | Hispanic | 21.88% (20.42-23.49) | 21.52% (20.09-23.18) | -1.62% (-5.37-2.06) | 33.7% (-405.67-507.06) |
| **1939** | Hispanic | 22.2% (20.73-23.76) | 21.44% (20.04-23.14) | -3.44% (-7.04-0.44) | 61.82% (-285.27-365.14) |
| **1940** | Hispanic | 21.03% (19.58-22.65) | 20.37% (18.8-22.01) | -3.14% (-6.63-0.45) | -8% (-1711.43-1310.68) |
| **1941** | Hispanic | 17.03% (15.77-18.4) | 16.58% (15.28-17.99) | -2.61% (-6.86-1.2) | -10.83% (-35.78-4.07) |
| **1942** | Hispanic | 26.54% (24.86-28.35) | 24.63% (22.98-26.42) | -7.18% (-10.34--3.79) | 35.38% (19.18-53.16) |
| **1943** | Hispanic | 24.07% (22.41-25.71) | 23.05% (21.46-24.78) | -4.22% (-7.81--0.67) | 34.35% (6.18-76.57) |
| **1944** | Hispanic | 24.27% (22.69-25.96) | 23.57% (22.03-25.2) | -2.89% (-6.28-0.81) | 21.83% (-8.07-53.65) |
| **1945** | Hispanic | 21.04% (19.59-22.54) | 21.04% (19.59-22.54) | 0% (0-0) | 0% (0-0) |
| **1946** | Hispanic | 24.03% (22.28-25.75) | 23.09% (21.51-24.69) | -3.92% (-7.24--0.24) | 32.97% (2.02-71.45) |
| **1947** | Hispanic | 21.36% (19.96-22.75) | 21.77% (20.32-23.32) | 1.94% (-1.92-5.54) | -8.88% (-885.22-714.43) |
| **1948** | Hispanic | 23.97% (22.5-25.51) | 23.55% (22.04-25.07) | -1.77% (-5.2-1.89) | 15.77% (-18.32-45.99) |
| **1949** | Hispanic | 23.08% (21.58-24.75) | 22.3% (20.82-23.82) | -3.34% (-6.59-0.05) | 38.45% (-2.92-121.06) |
| **1950** | Hispanic | 22.94% (21.33-24.48) | 21.86% (20.38-23.42) | -4.7% (-7.85--1.07) | 58.21% (9.87-180.18) |
| **1951** | Hispanic | 19.6% (18.4-20.98) | 19.65% (18.37-21.16) | 0.27% (-3.45-4.17) | 2.77% (-124.41-105.47) |

| | | | | | |
|---|---|---|---|---|---|
| **1952** | Hispanic | 19.81% (18.4-21.29) | 19.48% (18.15-20.99) | -1.62% (-5.56-2.41) | -22.06% (-540.77-103.63) |
| **1953** | Hispanic | 20.42% (19.1-21.88) | 20.25% (18.9-21.79) | -0.84% (-4.44-3.5) | -9.12% (-596.28-500) |
| **1954** | Hispanic | 25.28% (23.65-26.94) | 24.5% (23.05-26.27) | -3.07% (-6.19-0.41) | 17.84% (-2.23-40.37) |
| **1955** | Hispanic | 25.71% (24.17-27.49) | 24.78% (23.26-26.52) | -3.61% (-6.93--0.32) | 20.18% (1.69-39.09) |
| **1956** | Hispanic | 23.5% (22.05-25.09) | 23.31% (21.9-24.96) | -0.78% (-4.22-3.19) | 8.82% (-39.3-42.34) |
| **1957** | Hispanic | 19.59% (18.27-20.93) | 19.5% (18.24-20.97) | -0.44% (-4.28-3.16) | -4.56% (-159.68-70.8) |
| **1958** | Hispanic | 21.66% (20.16-23.21) | 21.25% (19.84-22.74) | -1.86% (-5.46-1.88) | 45.4% (-619.1-519.73) |
| **1959** | Hispanic | 31.23% (29.55-33.03) | 29.02% (27.35-30.85) | -7.07% (-10.05--4.25) | 21.82% (12.97-31.27) |
| **1960** | Hispanic | 24.46% (22.82-26.29) | 23.18% (21.72-24.76) | -5.21% (-8.34--1.78) | 37.37% (14.68-67.77) |
| **1961** | Hispanic | 22.57% (21.26-24.11) | 21.34% (19.98-22.81) | -5.46% (-8.76--2.02) | 80% (21.51-357.18) |
| **1962** | Hispanic | 25.51% (24.08-27.21) | 24.31% (22.94-25.92) | -4.7% (-8.3--1.37) | 26.77% (7.56-47.71) |
| **1963** | Hispanic | 20.96% (19.75-22.24) | 21.02% (19.87-22.47) | 0.31% (-3.17-4.29) | 19.88% (-635.18-1144.38) |
| **1964** | Hispanic | 19.59% (18.39-20.98) | 19.81% (18.71-21.16) | 1.14% (-3.45-5.02) | 15.79% (-86.91-110.94) |
| **1965** | Hispanic | 22.15% (20.78-23.56) | 21.99% (20.71-23.46) | -0.73% (-4.31-3.4) | 17.59% (-158.28-243.05) |
| **1966** | Hispanic | 24.94% (23.18-26.97) | 24.73% (23-26.65) | -0.84% (-4.39-3.07) | 5.76% (-23.38-29.05) |
| **1916** | White | 16.92% (14.83-19.21) | 16.19% (14.18-18.4) | -4.27% (-8.45-0.34) | 13.34% (-1.01-27.76) |
| **1917** | White | 16.35% (14.62-18.13) | 15.91% (14.27-17.72) | -2.71% (-6.64-1.67) | 8.76% (-6.26-22.72) |

| | | | | | |
|---|---|---|---|---|---|
| **1918** | White | 18.46% (16.72-20.17) | 17.73% (16.12-19.43) | -3.94% (-7.96--0.08) | 10.19% (0.22-19.81) |
| **1919** | White | 13.19% (11.92-14.49) | 13.3% (12.04-14.67) | 0.86% (-3.69-5.04) | -5.42% (-58.06-27.83) |
| **1920** | White | 15.72% (14.27-17.24) | 15.29% (13.81-16.83) | -2.67% (-6.72-1.63) | 9.67% (-6.1-23.99) |
| **1921** | White | 14.32% (12.89-15.89) | 14.07% (12.6-15.49) | -1.67% (-5.91-3.07) | 7.93% (-16.67-27.84) |
| **1922** | White | 16.55% (15.04-18.08) | 15.64% (14.13-17.05) | -5.51% (-9.44--1.52) | 17.5% (5.03-29.33) |
| **1923** | White | 12.88% (11.56-14.15) | 12.72% (11.42-14.02) | -1.2% (-5.44-3.28) | 11.15% (-44.32-65.27) |
| **1924** | White | 15.35% (13.97-16.67) | 14.9% (13.54-16.22) | -2.96% (-6.89-1.03) | 11.4% (-4.32-26.61) |
| **1925** | White | 13.24% (12.13-14.41) | 12.93% (11.78-14.22) | -2.29% (-6.15-1.54) | 16.57% (-14.3-48.85) |
| **1926** | White | 15.71% (14.38-17.06) | 15.39% (14.05-16.79) | -2.01% (-6.08-2.19) | 7.31% (-8.41-21.43) |
| **1927** | White | 12.37% (11.31-13.44) | 12.25% (11.17-13.34) | -1.01% (-5.43-3.53) | 11.52% (-132.71-131.94) |
| **1928** | White | 16.7% (15.36-18.06) | 16.06% (14.78-17.43) | -3.76% (-8--0.12) | 11.65% (0.4-24.86) |
| **1929** | White | 18.08% (16.6-19.56) | 17.13% (15.76-18.56) | -5.23% (-8.83--1.3) | 14.32% (3.96-23.57) |
| **1930** | White | 16.53% (15.2-17.92) | 15.7% (14.39-17.06) | -4.97% (-9.23--1.08) | 15.88% (3.54-27.91) |
| **1931** | White | 14.09% (12.86-15.33) | 13.7% (12.55-14.96) | -2.76% (-6.94-1.43) | 14.13% (-8.79-36.63) |
| **1932** | White | 14.63% (13.47-15.86) | 14.02% (12.84-15.25) | -4.13% (-7.95--0.38) | 18.54% (1.86-36.22) |
| **1933** | White | 12.37% (11.33-13.47) | 12.25% (11.25-13.37) | -0.92% (-4.94-3.62) | 13.13% (-114.17-100.13) |
| **1934** | White | 12.49% (11.44-13.63) | 12.32% (11.35-13.45) | -1.34% (-5.36-3.1) | 15.84% (-80.83-78.68) |

| | | | | | |
|---|---|---|---|---|---|
| **1935** | White | 12.52% (11.46-13.68) | 12.4% (11.33-13.6) | -0.91% (-5.21-3.57) | 9.98% (-69.13-96.83) |
| **1936** | White | 13.29% (12.1-14.4) | 12.88% (11.78-14.16) | -3.01% (-7.11-1.66) | 21.61% (-13.94-56.3) |
| **1937** | White | 12.61% (11.51-13.76) | 12.41% (11.4-13.62) | -1.54% (-5.61-2.75) | 16.23% (-64.93-76.69) |
| **1938** | White | 12.48% (11.46-13.56) | 12.24% (11.22-13.4) | -1.89% (-5.93-2.36) | 21.17% (-45.49-121.41) |
| **1939** | White | 15.55% (14.3-16.8) | 14.82% (13.62-16.13) | -4.72% (-8.55--0.75) | 18.11% (2.65-32.1) |
| **1940** | White | 13.63% (12.37-14.76) | 13.15% (11.98-14.3) | -3.5% (-7.64-0.91) | 20.49% (-5.81-50.18) |
| **1941** | White | 10.6% (9.63-11.58) | 10.56% (9.61-11.53) | -0.29% (-5-4.43) | -0.13% (-200.63-272.07) |
| **1942** | White | 19% (17.54-20.38) | 17.62% (16.19-19.01) | -7.25% (-11.14--3.15) | 18.19% (7.63-28.28) |
| **1943** | White | 14.46% (13.26-15.71) | 13.87% (12.65-15.13) | -4.01% (-8-0.08) | 18.78% (-0.42-37.57) |
| **1944** | White | 14.79% (13.55-16.06) | 14.17% (12.96-15.41) | -4.16% (-8.37--0.04) | 17.95% (0.19-35.34) |
| **1945** | White | 11.32% (10.33-12.38) | 11.32% (10.33-12.38) | 0% (0-0) | 0% (0-0) |
| **1946** | White | 13.82% (12.61-15.03) | 13.21% (12.1-14.41) | -4.41% (-8.5-0.23) | 25.61% (-1.49-53.54) |
| **1947** | White | 13.53% (12.29-14.7) | 13.23% (12.12-14.53) | -2.2% (-6.48-2.27) | 12.96% (-16-42.15) |
| **1948** | White | 15.78% (14.57-17.15) | 14.99% (13.89-16.29) | -4.95% (-8.89--0.96) | 17.75% (3.52-31.17) |
| **1949** | White | 15.53% (14.32-16.86) | 14.49% (13.28-15.79) | -6.63% (-10.33--2.89) | 25.14% (10.44-38.8) |
| **1950** | White | 16.82% (15.46-18.18) | 15.46% (14.13-16.82) | -8.06% (-10.99--4.18) | 25.27% (13.21-34.49) |
| **1951** | White | 13.66% (12.46-14.95) | 13.13% (11.98-14.32) | -3.88% (-7.7-0.22) | 23.29% (-1.05-49.44) |

| | | | | | |
|---|---|---|---|---|---|
| **1952** | White | 14.87% (13.63-16.16) | 14.2% (12.99-15.36) | -4.5% (-7.97--0.66) | 19.38% (2.86-36.17) |
| **1953** | White | 14.89% (13.76-16.04) | 14.33% (13.14-15.41) | -3.71% (-7.68-0.66) | 15.56% (-2.63-32.6) |
| **1954** | White | 20.71% (19.12-22.34) | 19.41% (18.01-20.85) | -6.21% (-10.01--2.7) | 13.67% (6.05-21.36) |
| **1955** | White | 19.21% (17.75-20.64) | 18.14% (16.84-19.46) | -5.55% (-9.25--1.61) | 13.77% (4.03-22.27) |
| **1956** | White | 15.77% (14.52-16.96) | 15.45% (14.28-16.7) | -1.98% (-5.81-1.78) | 7% (-6.92-20.12) |
| **1957** | White | 13.21% (12.21-14.21) | 13.24% (12.2-14.33) | 0.22% (-3.82-4.19) | -1.39% (-39.26-27.08) |
| **1958** | White | 15.53% (14.4-16.68) | 15.47% (14.28-16.72) | -0.36% (-4.5-3.93) | 1.95% (-15.44-15.48) |
| **1959** | White | 24.3% (22.62-25.91) | 23.02% (21.37-24.61) | -5.27% (-8.85--1.46) | 9.99% (2.9-16.29) |
| **1960** | White | 18.01% (16.59-19.52) | 17.57% (16.11-19.03) | -2.41% (-6.05-1.96) | 6.33% (-5.51-16.4) |
| **1961** | White | 16.82% (15.53-18) | 16.38% (15.08-17.75) | -2.61% (-6.49-1.59) | 8.42% (-4.85-19) |
| **1962** | White | 18.95% (17.59-20.31) | 18.55% (17.21-19.94) | -2.07% (-5.79-1.88) | 5.14% (-5.02-14.13) |
| **1963** | White | 15.64% (14.5-16.79) | 15.84% (14.72-16.98) | 1.33% (-2.91-5.79) | -5% (-22.55-9.31) |
| **1964** | White | 15.3% (14.23-16.51) | 15.33% (14.26-16.49) | 0.2% (-3.71-4.35) | -0.85% (-17.88-14.14) |
| **1965** | White | 18.6% (17.16-20.07) | 18.08% (16.58-19.77) | -2.79% (-6.75-0.9) | 6.91% (-2.37-16.88) |
| **1966** | White | 22.34% (20.07-24.73) | 21.17% (19.06-23.46) | -5.23% (-8.82--1.48) | 10.72% (2.99-17.99) |

#### Section 2.4: BMI

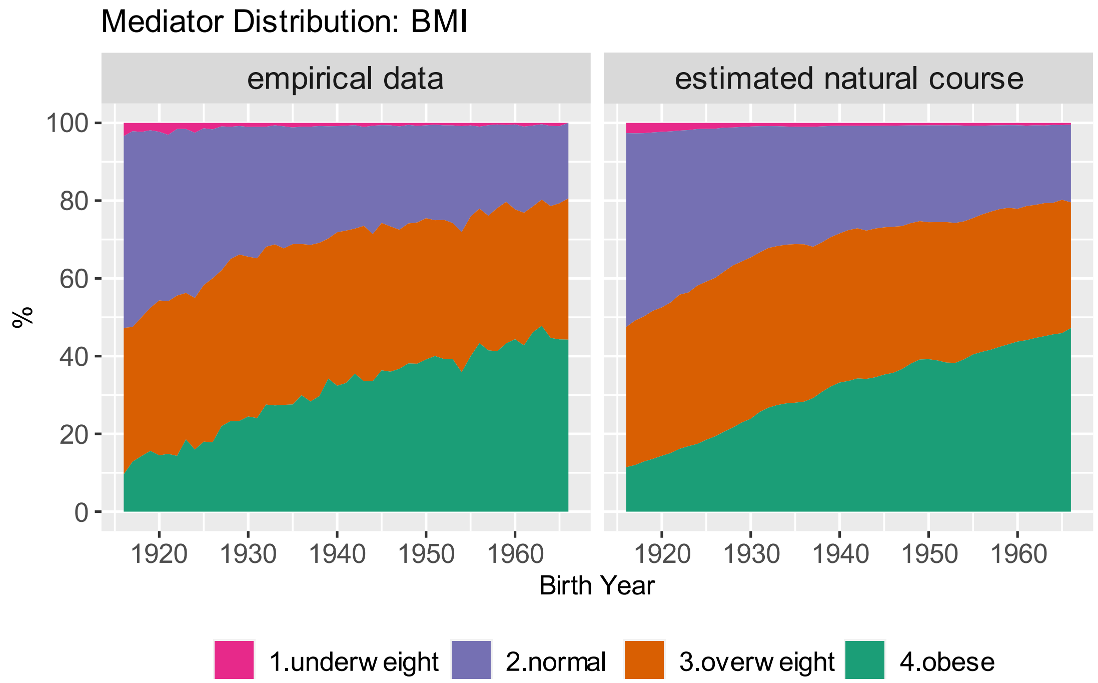

*Figure S. 13 Natural course distribution of smoking by birth cohort. "Empirical data" shows the descriptive distribution as observed in the data. "Estimated natural course" is estimated based on the mediator model without holding age and period constant.*

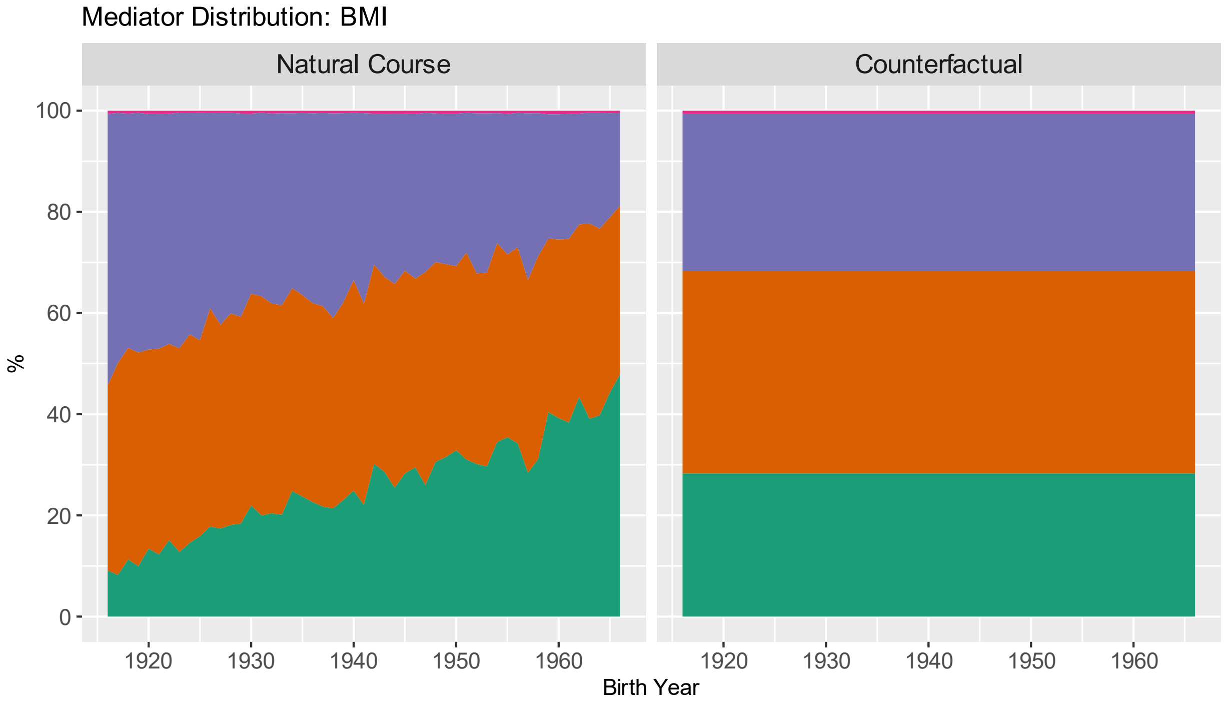

*Figure S. 14 Estimated distribution of BMI categories in natural course and counterfactual scenario by birth year. Age and period are held constant at age 50 and 1996 respectively.*

*Table S. 11 Predicted probability of elevated depressive symptoms in natural course and counterfactual scenario, relative difference, and contribution by birth cohort. All estimates are presented with 95% confidence intervals. Positive contributions imply that the counterfactual decreases depression risk and negative contributions imply that the counterfactual increases depression risk*

| | **Probability of elevated depressive symptoms (% (95%CI))** | | **% relative difference** | **% contribution** |
|---|---|---|---|---|
| **Birth Cohort** | Natural Course Scenario | Counterfactual Scenario | | |
| **1916** | 20.2% (18.32-22.09) | 20.9% (19.03-22.89) | 3.5% (2.02-4.88) | -11.09% (-17.56--6.19) |
| **1917** | 19.59% (18.21-21.11) | 20.33% (18.86-21.9) | 3.74% (2.29-5.22) | -12.84% (-18.63--7.72) |
| **1918** | 22.24% (20.96-23.69) | 22.96% (21.65-24.48) | 3.24% (1.86-4.58) | -8.74% (-12.46--4.86) |
| **1919** | 16.51% (15.49-17.69) | 17.16% (16.09-18.41) | 3.93% (2.59-5.32) | -24.65% (-43.45--14.55) |
| **1920** | 19.26% (18.1-20.63) | 19.81% (18.61-21.25) | 2.83% (1.48-4.37) | -10.06% (-16.31--5.22) |
| **1921** | 17.41% (16.23-18.73) | 17.95% (16.77-19.25) | 3.1% (1.65-4.51) | -15.21% (-26.04--7.75) |
| **1922** | 20.05% (18.78-21.43) | 20.45% (19.24-21.85) | 2.01% (0.65-3.41) | -6.44% (-11.92--2.09) |
| **1923** | 15.11% (14.17-16.14) | 15.56% (14.66-16.6) | 3% (1.49-4.48) | -37.3% (-175.83--14.34) |
| **1924** | 18.42% (17.42-19.48) | 18.88% (17.85-19.98) | 2.51% (1.33-3.88) | -10.03% (-16.74--5.39) |
| **1925** | 16.07% (15.15-17.03) | 16.43% (15.52-17.45) | 2.27% (0.73-3.81) | -16.61% (-35.31--5.09) |
| **1926** | 18.21% (17.23-19.22) | 18.62% (17.68-19.72) | 2.24% (0.7-3.6) | -9.4% (-16.74--3.06) |
| **1927** | 14.76% (13.86-15.64) | 15.09% (14.2-16.01) | 2.26% (0.71-3.81) | -35.63% (-226.73-48.34) |
| **1928** | 18.74% (17.75-19.7) | 19.09% (18.03-20.2) | 1.89% (0.54-3.19) | -7.4% (-12.99--2.11) |

| | | | | |
|---|---|---|---|---|
| **1929** | 20.74% (19.53-21.91) | 21.12% (19.83-22.27) | 1.86% (0.39-3.29) | -5.48% (-10.17--1.15) |
| **1930** | 19.99% (18.81-21.09) | 20.22% (18.98-21.42) | 1.19% (-0.11-2.55) | -3.88% (-8.51-0.36) |
| **1931** | 16.74% (15.65-17.71) | 17.02% (15.91-18.04) | 1.65% (0.06-3.16) | -9.36% (-21.21--0.29) |
| **1932** | 17.8% (16.72-18.82) | 18.01% (16.96-19.15) | 1.2% (-0.23-2.63) | -5.42% (-12.6-1.06) |
| **1933** | 14.46% (13.61-15.33) | 14.7% (13.84-15.57) | 1.72% (0.04-3.26) | -35.62% (-327.54-314.39) |
| **1934** | 15.92% (14.9-16.88) | 15.99% (15-16.99) | 0.46% (-1.06-1.94) | -3.62% (-19.49-8.03) |
| **1935** | 15.07% (14.14-16.05) | 15.21% (14.28-16.22) | 0.96% (-0.58-2.49) | -12.08% (-67.8-8.31) |
| **1936** | 16.17% (15.22-17.27) | 16.26% (15.28-17.43) | 0.62% (-0.85-2.25) | -4.1% (-18.33-6.44) |
| **1937** | 14.27% (13.39-15.15) | 14.46% (13.55-15.39) | 1.32% (-0.06-2.9) | -22.27% (-640.96-622.55) |
| **1938** | 14.78% (13.84-15.77) | 14.96% (13.98-15.98) | 1.19% (-0.29-2.78) | -17.53% (-153.62-34.76) |
| **1939** | 17.91% (16.86-18.99) | 18% (16.98-19.11) | 0.52% (-0.91-2) | -2.51% (-9.84-4.18) |
| **1940** | 15.23% (14.32-16.09) | 15.26% (14.3-16.15) | 0.22% (-1.4-1.73) | -2.25% (-29.39-18.81) |
| **1941** | 12.6% (11.72-13.43) | 12.72% (11.84-13.57) | 0.92% (-0.77-2.58) | 9.31% (-9.2-41.16) |
| **1942** | 21.48% (20.33-22.6) | 21.36% (20.19-22.5) | -0.56% (-1.78-0.67) | 1.62% (-1.84-5.14) |
| **1943** | 16.74% (15.76-17.67) | 16.72% (15.75-17.7) | -0.08% (-1.61-1.53) | 0.51% (-9.18-9.99) |
| **1944** | 16.12% (15.25-17.04) | 16.17% (15.31-17.13) | 0.34% (-1.22-1.73) | -2.8% (-14.25-8.97) |
| **1945** | 13.87% (13.02-14.74) | 13.87% (13.02-14.74) | 0% (0-0) | 0% (0-0) |

| | | | | |
|---|---|---|---|---|
| **1946** | 17.49% (16.33-18.61) | 17.37% (16.37-18.47) | -0.67% (-2.23-0.78) | 3.29% (-3.99-11.36) |
| **1947** | 16.64% (15.64-17.63) | 16.68% (15.68-17.7) | 0.28% (-1.17-1.69) | -1.57% (-11.33-6.66) |
| **1948** | 18.49% (17.46-19.51) | 18.35% (17.34-19.37) | -0.77% (-2.26-0.62) | 3.08% (-2.41-8.72) |
| **1949** | 18.99% (17.98-20.04) | 18.73% (17.76-19.81) | -1.38% (-2.77-0.06) | 5.13% (-0.2-10.34) |
| **1950** | 20.59% (19.57-21.75) | 20.2% (19.2-21.36) | -1.9% (-3.25--0.6) | 5.75% (1.82-10.06) |
| **1951** | 16.49% (15.6-17.47) | 16.33% (15.43-17.36) | -1% (-2.55-0.63) | 6.27% (-4.15-16.75) |
| **1952** | 17.65% (16.78-18.72) | 17.56% (16.66-18.65) | -0.53% (-1.92-0.83) | 2.31% (-3.98-9.03) |
| **1953** | 18.26% (17.43-19.27) | 18.19% (17.32-19.22) | -0.4% (-1.8-1.05) | 1.65% (-4.44-7.37) |
| **1954** | 24.08% (22.96-25.21) | 23.76% (22.68-24.91) | -1.34% (-2.67--0.11) | 3.21% (0.28-6.19) |
| **1955** | 22.76% (21.78-23.9) | 22.38% (21.4-23.51) | -1.67% (-3.07--0.35) | 4.27% (0.87-7.83) |
| **1956** | 18.88% (17.97-19.85) | 18.63% (17.74-19.64) | -1.28% (-2.69-0.24) | 4.8% (-0.91-10.22) |
| **1957** | 14.55% (13.83-15.35) | 14.5% (13.8-15.22) | -0.3% (-1.75-1.38) | 6.39% (-93.19-105.56) |
| **1958** | 17.8% (16.94-18.73) | 17.67% (16.81-18.6) | -0.72% (-2.2-0.77) | 3.31% (-3.38-10.18) |
| **1959** | 28.11% (26.9-29.45) | 27.47% (26.28-28.72) | -2.29% (-3.53--1.08) | 4.49% (2.12-6.99) |
| **1960** | 21.5% (20.44-22.66) | 20.99% (19.88-22.12) | -2.36% (-3.68--1.04) | 6.69% (2.95-10.33) |
| **1961** | 19.76% (18.79-20.8) | 19.24% (18.31-20.28) | -2.6% (-4.09--1.11) | 8.67% (3.74-13.75) |
| **1962** | 22.08% (21.1-23.15) | 21.37% (20.41-22.44) | -3.21% (-4.64--1.6) | 8.66% (4.26-12.59) |

| | | | | |
|---|---|---|---|---|
| **1963** | 17.11% (16.37-17.9) | 16.75% (16.01-17.57) | -2.09% (-3.63--0.55) | 11.1% (3.04-19.51) |
| **1964** | 16.81% (16.07-17.55) | 16.38% (15.62-17.13) | -2.52% (-3.95--1.08) | 14.3% (6.67-23.15) |
| **1965** | 20.97% (19.92-22.03) | 20.29% (19.3-21.31) | -3.23% (-4.56--1.86) | 9.52% (5.55-13.48) |
| **1966** | 22.73% (21.29-24.19) | 21.85% (20.5-23.21) | -3.85% (-5.24--2.5) | 9.81% (6.56-13.69) |

*Table S. 12 Predicted Probability of elevated depressive symptoms in natural course and counterfactual scenario, relative difference, and contribution by birth cohort and* ***stratified by sex****. All estimates are presented with 95% confidence intervals. Positive contributions imply that the counterfactual decreases depression risk and negative contributions imply that the counterfactual increases depression risk*

| | | **Probability of elevated depressive symptoms (% (95%CI))** | | **% relative difference** | **% contribution** |
|---|---|---|---|---|---|
| **Birth Cohort** | sex | Natural Course Scenario | Counterfactual Scenario | | |
| **1916** | female | 21.2% (18.84-23.75) | 22.35% (19.96-24.92) | 5.4% (3.63-7.2) | -18.57% (-31.7--11.81) |
| **1917** | female | 23.61% (21.55-25.74) | 24.87% (22.74-27.11) | 5.34% (3.8-6.86) | -14.71% (-20.66--10.29) |
| **1918** | female | 24.96% (22.99-26.97) | 26.13% (24.08-28.21) | 4.67% (3.12-6.38) | -11.65% (-16.66--7.64) |
| **1919** | female | 19.96% (18.42-21.63) | 21.09% (19.45-22.79) | 5.65% (4-7.43) | -22.91% (-35.48--14.94) |
| **1920** | female | 20.51% (18.81-22.07) | 21.34% (19.65-23.06) | 4.07% (2.37-5.8) | -15.27% (-24.24--8.54) |
| **1921** | female | 20.23% (18.55-21.8) | 21.2% (19.47-22.92) | 4.78% (2.99-6.41) | -18.87% (-29.81--11.34) |
| **1922** | female | 20.73% (19.08-22.42) | 21.47% (19.73-23.25) | 3.57% (1.96-5.21) | -13.11% (-20.67--6.69) |
| **1923** | female | 16.53% (15.16-17.82) | 17.45% (16.05-18.89) | 5.56% (3.68-7.4) | -60.58% (-294.8--24.22) |
| **1924** | female | 21.02% (19.46-22.57) | 21.81% (20.22-23.43) | 3.77% (2.07-5.41) | -13.15% (-21.44--6.91) |

| | | | | | |
|---|---|---|---|---|---|
| **1925** | female | 17.79% (16.41-19.14) | 18.42% (17.04-19.85) | 3.59% (1.84-5.62) | -23.34% (-49.47--10.47) |
| **1926** | female | 22.22% (20.66-23.84) | 22.89% (21.3-24.44) | 3.05% (1.47-4.69) | -9.51% (-15.28--4.52) |
| **1927** | female | 15.72% (14.55-16.99) | 16.33% (15.11-17.64) | 3.91% (2.06-5.61) | -71.07% (-1024.74-617.62) |
| **1928** | female | 21.49% (20.05-22.98) | 22.19% (20.74-23.8) | 3.28% (1.58-4.76) | -10.97% (-17.47--5.07) |
| **1929** | female | 22.57% (21.05-24.32) | 23.36% (21.81-25.08) | 3.48% (1.72-5.01) | -10.5% (-15.97--5.29) |
| **1930** | female | 21.44% (19.91-22.98) | 21.94% (20.37-23.51) | 2.34% (0.71-3.85) | -7.91% (-14.11--2.29) |
| **1931** | female | 18.74% (17.4-20.12) | 19.28% (17.87-20.78) | 2.9% (0.95-4.73) | -14.75% (-27.7--5.03) |
| **1932** | female | 18.93% (17.65-20.3) | 19.37% (18.05-20.77) | 2.34% (0.62-3.98) | -11.47% (-22.07--3.08) |
| **1933** | female | 15.83% (14.65-16.99) | 16.29% (15.04-17.56) | 2.92% (0.97-4.67) | -50.89% (-649.69-492.51) |
| **1934** | female | 16.05% (14.81-17.33) | 16.23% (15-17.52) | 1.13% (-0.96-2.9) | -15.66% (-135.08-122.41) |
| **1935** | female | 15.8% (14.57-17.01) | 16.09% (14.87-17.32) | 1.87% (0-3.76) | -30.44% (-421.68-381.64) |
| **1936** | female | 16.64% (15.4-17.95) | 16.94% (15.66-18.23) | 1.8% (-0.05-3.76) | -18.65% (-74.63-1.84) |
| **1937** | female | 15.56% (14.39-16.73) | 15.99% (14.78-17.21) | 2.73% (0.89-4.52) | -50.98% (-836.55-705.27) |
| **1938** | female | 15.42% (14.25-16.57) | 15.9% (14.68-17.11) | 3.12% (1.24-4.78) | -56.66% (-1264.91-958.06) |
| **1939** | female | 19.65% (18.32-20.97) | 19.9% (18.52-21.22) | 1.27% (-0.27-2.77) | -5.49% (-13.17-1.15) |
| **1940** | female | 16.66% (15.38-18.03) | 16.84% (15.57-18.2) | 1.1% (-0.73-2.8) | -11.79% (-67.23-8.78) |
| **1941** | female | 13.45% (12.42-14.6) | 13.8% (12.73-14.99) | 2.59% (0.8-4.51) | 21.38% (5.19-83.55) |

| | | | | | |
|---|---|---|---|---|---|
| **1942** | female | 21.76% (20.27-23.28) | 21.68% (20.19-23.29) | -0.35% (-1.97-1.35) | 1.06% (-4.54-6.33) |
| **1943** | female | 17.43% (16.28-18.73) | 17.48% (16.29-18.78) | 0.31% (-1.39-1.97) | -2.27% (-19.25-10.24) |
| **1944** | female | 18.13% (16.98-19.43) | 18.36% (17.13-19.66) | 1.22% (-0.39-2.94) | -7.35% (-20.47-2.45) |
| **1945** | female | 15.04% (13.92-16.2) | 15.04% (13.92-16.2) | 0% (0-0) | 0% (0-0) |
| **1946** | female | 17.66% (16.41-18.94) | 17.58% (16.31-18.9) | -0.47% (-2.24-1.23) | 3.4% (-10.21-16.51) |
| **1947** | female | 19.95% (18.66-21.35) | 19.94% (18.52-21.27) | -0.03% (-1.86-1.64) | 0.06% (-7.37-7.42) |
| **1948** | female | 19.27% (17.98-20.51) | 19.12% (17.86-20.38) | -0.73% (-2.33-0.99) | 3.51% (-4.92-10.72) |
| **1949** | female | 19.21% (17.99-20.45) | 18.93% (17.66-20.22) | -1.44% (-3.14-0.27) | 6.91% (-1.37-14.7) |
| **1950** | female | 19.61% (18.32-20.83) | 19.21% (17.97-20.5) | -2.05% (-3.65--0.43) | 8.86% (1.72-16.74) |
| **1951** | female | 17.54% (16.47-18.85) | 17.4% (16.28-18.63) | -0.84% (-2.65-0.92) | 5.87% (-7.24-21.52) |
| **1952** | female | 18.55% (17.46-19.87) | 18.49% (17.4-19.69) | -0.28% (-2.14-1.56) | 1.41% (-8.3-11.11) |
| **1953** | female | 19.95% (18.78-21.29) | 19.9% (18.78-21.18) | -0.25% (-2.13-1.41) | 0.81% (-6.09-8.14) |
| **1954** | female | 26.43% (24.9-28.1) | 25.95% (24.5-27.64) | -1.82% (-3.19--0.44) | 4.3% (1.03-7.44) |
| **1955** | female | 24.2% (22.78-25.63) | 23.6% (22.27-25.09) | -2.47% (-3.96--0.94) | 6.54% (2.49-10.69) |
| **1956** | female | 21.61% (20.21-23.07) | 21.17% (19.88-22.59) | -2.04% (-3.88--0.44) | 6.54% (1.54-12.93) |
| **1957** | female | 16.51% (15.4-17.62) | 16.61% (15.54-17.82) | 0.61% (-1.22-2.49) | -6.68% (-48.96-18.2) |
| **1958** | female | 21.55% (20.21-23.01) | 21.37% (20.05-22.85) | -0.85% (-2.67-0.87) | 2.69% (-2.84-8.58) |

| | | | | | |
|---|---|---|---|---|---|
| **1959** | female | 29.59% (27.98-31.26) | 28.64% (26.99-30.39) | -3.2% (-4.61--1.72) | 6.5% (3.52-9.55) |
| **1960** | female | 23.21% (21.79-24.77) | 22.45% (20.97-23.92) | -3.29% (-4.89--1.63) | 9.32% (4.46-13.97) |
| **1961** | female | 21.51% (20.18-22.86) | 20.81% (19.47-22.13) | -3.26% (-4.92--1.77) | 10.89% (6.01-17.1) |
| **1962** | female | 24.23% (22.93-25.74) | 23.13% (21.87-24.48) | -4.53% (-6.04--2.94) | 11.96% (7.74-16.32) |
| **1963** | female | 20.79% (19.71-22.02) | 20.18% (19.08-21.48) | -2.96% (-4.54--1.33) | 10.72% (5.01-16.79) |
| **1964** | female | 19.68% (18.56-20.92) | 19.1% (18.04-20.34) | -2.93% (-4.61--1.16) | 12.58% (4.99-20.19) |
| **1965** | female | 23.97% (22.35-25.93) | 22.96% (21.42-24.79) | -4.2% (-5.81--2.53) | 11.4% (6.84-15.88) |
| **1966** | female | 25.66% (23.46-28.17) | 24.51% (22.44-26.91) | -4.51% (-6.01--2.85) | 10.98% (7-15.08) |
| **1916** | male | 21.45% (18.59-24.35) | 21.09% (18.24-24.03) | -1.68% (-3.21--0.12) | 4.54% (0.33-8.9) |
| **1917** | male | 22.58% (20.11-24.97) | 22.33% (19.92-24.8) | -1.13% (-2.51-0.24) | 2.77% (-0.55-6.29) |
| **1918** | male | 24.07% (21.89-26.31) | 24.03% (21.77-26.35) | -0.19% (-1.5-1.2) | 0.5% (-2.76-3.31) |
| **1919** | male | 19.05% (17.2-20.97) | 19.03% (17.13-20.96) | -0.13% (-1.53-1.1) | 0.41% (-3.88-5.05) |
| **1920** | male | 19.62% (17.71-21.69) | 19.62% (17.7-21.59) | 0.01% (-1.12-1.36) | 0.04% (-4.23-3.64) |
| **1921** | male | 18.29% (16.49-20.31) | 18.33% (16.5-20.32) | 0.23% (-0.98-1.53) | -0.82% (-6.41-3.68) |
| **1922** | male | 18.38% (16.51-20.37) | 18.44% (16.62-20.42) | 0.33% (-0.96-1.56) | -1.1% (-5.52-3.51) |
| **1923** | male | 14.49% (13.02-16.02) | 14.62% (13.12-16.26) | 0.85% (-0.3-2.11) | -8.6% (-91.95-73.08) |
| **1924** | male | 17.22% (15.55-18.89) | 17.32% (15.73-19.05) | 0.59% (-0.66-2.02) | -2.49% (-9.39-2.85) |

| | | | | | |
|---|---|---|---|---|---|
| **1925** | male | 14.75% (13.25-16.31) | 14.76% (13.24-16.35) | 0.07% (-1.25-1.36) | -0.38% (-38.93-24.01) |
| **1926** | male | 18.06% (16.28-19.77) | 18.1% (16.3-19.91) | 0.24% (-0.96-1.44) | -0.76% (-6.13-4.1) |
| **1927** | male | 13.47% (12.02-14.83) | 13.54% (12.11-14.99) | 0.51% (-0.71-1.93) | -3.12% (-129.59-239.32) |
| **1928** | male | 18.04% (16.3-19.61) | 18.08% (16.39-19.77) | 0.2% (-0.98-1.42) | -0.73% (-6.05-4.03) |
| **1929** | male | 19.44% (17.61-21.21) | 19.57% (17.7-21.41) | 0.7% (-0.57-1.94) | -2.16% (-6.56-1.85) |
| **1930** | male | 19.36% (17.51-21.1) | 19.39% (17.56-21.26) | 0.14% (-1.16-1.37) | -0.43% (-4.38-3.48) |
| **1931** | male | 17.63% (16.03-19.26) | 17.7% (16.04-19.4) | 0.37% (-0.91-1.74) | -1.57% (-7.04-4.16) |
| **1932** | male | 17.47% (15.84-19.14) | 17.46% (15.81-19.16) | -0.01% (-1.29-1.1) | -0.12% (-5.01-5.55) |
| **1933** | male | 15.32% (13.93-16.85) | 15.34% (13.95-16.87) | 0.1% (-1.28-1.32) | -0.84% (-14.4-10.82) |
| **1934** | male | 16.09% (14.53-17.73) | 16.09% (14.52-17.68) | -0.02% (-1.22-1.34) | 0.12% (-8.14-8.56) |
| **1935** | male | 15.72% (14.23-17.27) | 15.7% (14.21-17.26) | -0.11% (-1.34-1.17) | 1.08% (-9.65-10.22) |
| **1936** | male | 16.28% (14.8-17.83) | 16.22% (14.75-17.81) | -0.32% (-1.55-0.96) | 1.76% (-6.64-9.86) |
| **1937** | male | 14.96% (13.63-16.4) | 14.97% (13.57-16.4) | 0.05% (-1.14-1.3) | -0.38% (-25.28-16.04) |
| **1938** | male | 14.99% (13.61-16.4) | 14.96% (13.54-16.38) | -0.17% (-1.51-1.16) | 1.77% (-15.5-20.44) |
| **1939** | male | 18.24% (16.58-19.86) | 18.16% (16.52-19.81) | -0.46% (-1.79-0.81) | 1.76% (-3.23-6.57) |
| **1940** | male | 15.8% (14.33-17.3) | 15.79% (14.32-17.29) | -0.08% (-1.4-1.2) | 0.56% (-9.58-10.78) |
| **1941** | male | 12.7% (11.45-14.02) | 12.69% (11.43-14.06) | -0.15% (-1.51-1.3) | -2.96% (-118.97-118.9) |

| | | | | | |
|---|---|---|---|---|---|
| **1942** | male | 19.48% (17.76-21.26) | 19.47% (17.7-21.27) | -0.02% (-1.26-1.22) | -0.02% (-3.82-4.01) |
| **1943** | male | 15.68% (14.21-17.16) | 15.71% (14.2-17.19) | 0.21% (-1.07-1.63) | -1.23% (-13.06-7.52) |
| **1944** | male | 15.53% (14.07-17.1) | 15.55% (14.09-17.08) | 0.15% (-1.04-1.43) | -0.9% (-13.31-8.54) |
| **1945** | male | 13.27% (11.91-14.75) | 13.27% (11.91-14.75) | 0% (0-0) | 0% (0-0) |
| **1946** | male | 15.25% (13.75-16.88) | 15.26% (13.79-16.86) | 0.07% (-1.1-1.3) | -0.5% (-16.58-11.15) |
| **1947** | male | 17.55% (15.95-19.24) | 17.57% (15.99-19.25) | 0.12% (-1.15-1.42) | -0.49% (-6.38-4.94) |
| **1948** | male | 17.8% (16.31-19.36) | 17.78% (16.3-19.37) | -0.13% (-1.27-1.07) | 0.49% (-4.45-5.36) |
| **1949** | male | 19.04% (17.52-20.63) | 18.99% (17.45-20.57) | -0.24% (-1.44-0.9) | 0.77% (-3.07-4.9) |
| **1950** | male | 20.33% (18.65-22.04) | 20.24% (18.55-21.9) | -0.43% (-1.58-0.77) | 1.32% (-2.16-4.67) |
| **1951** | male | 18.95% (17.42-20.57) | 18.87% (17.24-20.53) | -0.4% (-1.55-1.04) | 1.5% (-3.36-5.45) |
| **1952** | male | 19.33% (17.74-20.95) | 19.24% (17.57-20.89) | -0.46% (-1.63-0.79) | 1.36% (-2.58-5.43) |
| **1953** | male | 19.96% (18.33-21.69) | 19.86% (18.2-21.6) | -0.5% (-1.69-0.63) | 1.52% (-1.85-4.91) |
| **1954** | male | 25.34% (23.43-27.37) | 25.22% (23.25-27.2) | -0.47% (-1.55-0.56) | 1% (-1.19-3.29) |
| **1955** | male | 22.28% (20.58-24.04) | 22.12% (20.37-23.94) | -0.7% (-1.85-0.48) | 1.71% (-1.13-4.58) |
| **1956** | male | 19.47% (17.87-21.12) | 19.38% (17.79-21.07) | -0.44% (-1.7-0.8) | 1.34% (-2.49-5.45) |
| **1957** | male | 14.74% (13.43-16.14) | 14.69% (13.35-16.1) | -0.33% (-1.59-0.93) | 3.08% (-16.95-32.9) |
| **1958** | male | 18.27% (16.76-19.78) | 18.18% (16.65-19.75) | -0.48% (-1.67-0.77) | 1.76% (-3-6.25) |

| | | | | | |
|---|---|---|---|---|---|
| **1959** | male | 24.33% (22.43-26.28) | 24.1% (22.19-26) | -0.92% (-2.08-0.28) | 2.04% (-0.64-4.55) |
| **1960** | male | 20.08% (18.36-21.84) | 19.88% (18.11-21.55) | -1.03% (-2.51-0.3) | 2.98% (-0.89-7.65) |
| **1961** | male | 18.08% (16.57-19.58) | 17.87% (16.39-19.38) | -1.15% (-2.51-0.19) | 4.23% (-0.69-10.01) |
| **1962** | male | 20.35% (18.9-21.93) | 20.11% (18.58-21.74) | -1.19% (-2.51--0.01) | 3.46% (0.02-7.22) |
| **1963** | male | 17.1% (15.88-18.46) | 16.95% (15.75-18.36) | -0.85% (-2.04-0.63) | 3.88% (-2.94-10.02) |
| **1964** | male | 16.27% (14.96-17.65) | 16.11% (14.86-17.52) | -1.02% (-2.38-0.22) | 5.37% (-1.11-14.93) |
| **1965** | male | 18.78% (17.14-20.45) | 18.5% (16.85-20.24) | -1.49% (-2.79--0.11) | 5.18% (0.38-9.85) |
| **1966** | male | 19.91% (17.83-21.93) | 19.58% (17.51-21.6) | -1.67% (-3.08--0.25) | 5.08% (0.67-10.19) |

*Table S. 13 Predicted Probability of elevated depressive symptoms in natural course and counterfactual scenario, relative difference, and contribution by birth cohort and* ***stratified by race/ethnicity****. All estimates are presented with 95% confidence intervals. Positive contributions imply that the counterfactual decreases depression risk and negative contributions imply that the counterfactual increases depression risk*

| | | **Probability of elevated depressive symptoms (% (95%CI))** | | **% relative difference** | **% contribution** |
|---|---|---|---|---|---|
| **Birth Cohort** | race/ethnicity | Natural Course Scenario | Counterfactual Scenario | | |
| **1916** | Black | 33.41% (30.17-36.63) | 33.46% (30.29-36.69) | 0.17% (-0.78-1.06) | -0.51% (-2.82-1.99) |
| **1917** | Black | 30.97% (28.36-33.64) | 31.09% (28.46-33.74) | 0.37% (-0.56-1.2) | -1.16% (-3.37-1.58) |
| **1918** | Black | 33.56% (31.1-35.92) | 33.69% (31.29-36.06) | 0.39% (-0.51-1.25) | -0.96% (-3.14-1.26) |
| **1919** | Black | 25.66% (23.53-27.59) | 25.87% (23.82-27.79) | 0.82% (-0.11-1.72) | -3.96% (-8.83-0.45) |
| **1920** | Black | 28.44% (26.28-30.49) | 28.62% (26.46-30.74) | 0.63% (-0.2-1.41) | -2.19% (-5.35-0.67) |

| | | | | | |
|---|---|---|---|---|---|
| **1921** | Black | 25.83% (23.84-27.79) | 26.04% (24.02-28.04) | 0.8% (-0.09-1.64) | -3.82% (-8.78-0.37) |
| **1922** | Black | 28.01% (25.99-30.07) | 28.14% (26.07-30.12) | 0.45% (-0.41-1.3) | -1.56% (-5.07-1.5) |
| **1923** | Black | 23.03% (21.29-24.79) | 23.22% (21.39-24.95) | 0.8% (-0.08-1.78) | -6.49% (-22.04-0.55) |
| **1924** | Black | 24.72% (23.04-26.4) | 24.89% (23.22-26.61) | 0.69% (-0.12-1.76) | -3.59% (-10.35-0.76) |
| **1925** | Black | 21.21% (19.68-22.74) | 21.37% (19.87-22.94) | 0.75% (-0.16-1.71) | -12.06% (-242.04-110.5) |
| **1926** | Black | 23.61% (21.98-25.29) | 23.82% (22.27-25.41) | 0.88% (-0.11-1.84) | -6.17% (-16.73-0.74) |
| **1927** | Black | 19.09% (17.73-20.51) | 19.25% (17.95-20.7) | 0.84% (-0.19-1.89) | 12.15% (-59.97-106.08) |
| **1928** | Black | 24.18% (22.71-25.74) | 24.33% (22.9-25.95) | 0.65% (-0.21-1.6) | -3.97% (-11.32-1.31) |
| **1929** | Black | 26.37% (24.69-28.21) | 26.5% (24.88-28.35) | 0.49% (-0.39-1.39) | -1.98% (-6.78-1.59) |
| **1930** | Black | 24.37% (22.88-26.09) | 24.47% (22.96-26.13) | 0.42% (-0.43-1.34) | -2.43% (-8.21-2.5) |
| **1931** | Black | 21.51% (20.02-23.01) | 21.62% (20.18-23.15) | 0.55% (-0.33-1.44) | -8.46% (-88.58-36.98) |
| **1932** | Black | 22.11% (20.72-23.57) | 22.14% (20.77-23.6) | 0.16% (-0.8-1.15) | -1.76% (-24.31-10.35) |
| **1933** | Black | 21.07% (19.67-22.58) | 21.14% (19.76-22.66) | 0.31% (-0.65-1.2) | -4.51% (-115.76-80.5) |
| **1934** | Black | 19.81% (18.46-21.3) | 19.81% (18.47-21.27) | -0.01% (-0.93-0.94) | -0.09% (-191.89-139.27) |
| **1935** | Black | 19.99% (18.62-21.43) | 19.99% (18.63-21.46) | 0.01% (-0.87-1.03) | 0.66% (-195.44-99.92) |
| **1936** | Black | 21.22% (19.79-22.71) | 21.19% (19.81-22.69) | -0.16% (-1.05-0.78) | 3.41% (-92.74-125.72) |
| **1937** | Black | 19.46% (18.14-20.84) | 19.48% (18.19-20.78) | 0.07% (-0.82-0.98) | 1.24% (-168.35-88.34) |

| | | | | | |
|---|---|---|---|---|---|
| **1938** | Black | 18.19% (17.05-19.5) | 18.24% (17.06-19.52) | 0.24% (-0.71-1.32) | 2.19% (-7.96-17.19) |
| **1939** | Black | 19.98% (18.66-21.41) | 20.04% (18.76-21.44) | 0.28% (-0.58-1.3) | 3.42% (-107.66-174.66) |
| **1940** | Black | 18.14% (16.89-19.51) | 18.26% (17.02-19.65) | 0.67% (-0.24-1.56) | 5.7% (-2.33-24.43) |
| **1941** | Black | 14.52% (13.44-15.76) | 14.64% (13.54-15.88) | 0.86% (-0.21-1.89) | 2.16% (-0.54-4.86) |
| **1942** | Black | 24.82% (23.18-26.48) | 24.88% (23.2-26.62) | 0.22% (-0.59-0.99) | -1.27% (-6.43-3.11) |
| **1943** | Black | 20.93% (19.51-22.54) | 20.99% (19.57-22.58) | 0.28% (-0.58-1.24) | -3.63% (-194.28-117.71) |
| **1944** | Black | 23.68% (22.14-25.4) | 23.7% (22.24-25.42) | 0.09% (-0.78-0.93) | -0.56% (-7.41-5.39) |
| **1945** | Black | 20.2% (18.79-21.8) | 20.2% (18.79-21.8) | 0% (0-0) | 0% (0-0) |
| **1946** | Black | 26.18% (24.37-28.19) | 26.01% (24.23-27.95) | -0.64% (-1.43-0.23) | 2.86% (-1.08-6.73) |
| **1947** | Black | 24.34% (22.66-26.15) | 24.27% (22.62-26.07) | -0.32% (-1.16-0.6) | 1.96% (-3.73-7.66) |
| **1948** | Black | 27.65% (26.03-29.4) | 27.46% (25.85-29.26) | -0.69% (-1.48-0.12) | 2.56% (-0.46-5.54) |
| **1949** | Black | 26.83% (25.3-28.53) | 26.59% (25.02-28.29) | -0.87% (-1.73--0.04) | 3.42% (0.2-7.09) |
| **1950** | Black | 27.14% (25.6-28.78) | 26.88% (25.33-28.54) | -0.95% (-1.82--0.18) | 3.72% (0.72-7.3) |
| **1951** | Black | 22.37% (20.98-23.83) | 22.24% (20.86-23.67) | -0.57% (-1.5-0.32) | 5.83% (-3.85-20.33) |
| **1952** | Black | 24.44% (23.04-25.95) | 24.27% (22.84-25.77) | -0.71% (-1.54-0.17) | 4.24% (-0.97-9.55) |
| **1953** | Black | 24.57% (23.16-26.1) | 24.4% (22.99-25.93) | -0.69% (-1.56-0.31) | 3.85% (-1.79-9.64) |
| **1954** | Black | 32.49% (30.77-34.3) | 32.25% (30.54-34) | -0.74% (-1.61-0.04) | 1.92% (-0.1-4.18) |

| | | | | | |
|---|---|---|---|---|---|
| **1955** | Black | 31.45% (29.74-33.22) | 31.17% (29.48-32.88) | -0.89% (-1.64--0.1) | 2.52% (0.27-4.5) |
| **1956** | Black | 25.62% (24.03-27.22) | 25.45% (23.93-27.09) | -0.66% (-1.6-0.25) | 3.18% (-1.21-7.99) |
| **1957** | Black | 21.2% (19.86-22.58) | 21.07% (19.74-22.43) | -0.62% (-1.6-0.27) | 10.16% (-130.19-119.55) |
| **1958** | Black | 24.62% (23.15-26.18) | 24.46% (23.03-26.1) | -0.65% (-1.52-0.22) | 3.71% (-1.5-9.5) |
| **1959** | Black | 32.91% (31.17-34.78) | 32.58% (30.83-34.41) | -1.02% (-1.85--0.23) | 2.62% (0.59-4.85) |
| **1960** | Black | 24.62% (23.05-26.16) | 24.42% (22.84-26) | -0.82% (-1.75-0.12) | 4.68% (-0.74-10.08) |
| **1961** | Black | 22.03% (20.66-23.43) | 21.83% (20.43-23.26) | -0.93% (-1.83--0.01) | 10.77% (-0.55-46.14) |
| **1962** | Black | 23.83% (22.38-25.25) | 23.62% (22.17-25.02) | -0.85% (-1.63--0.07) | 5.6% (0.47-12.12) |
| **1963** | Black | 19.39% (18.25-20.56) | 19.29% (18.14-20.46) | -0.52% (-1.49-0.48) | -9.75% (-164.42-190.92) |
| **1964** | Black | 18.15% (16.97-19.38) | 18.02% (16.84-19.2) | -0.73% (-1.6-0.27) | -6.45% (-25.88-3.03) |
| **1965** | Black | 22.4% (21.04-23.9) | 22.2% (20.79-23.68) | -0.88% (-1.75-0.07) | 8.75% (-0.73-27.74) |
| **1966** | Black | 25.63% (23.76-27.5) | 25.36% (23.52-27.27) | -1.05% (-1.9--0.21) | 4.93% (0.97-10.18) |
| **1916** | Hispanic | 12.13% (10.03-14.15) | 12.47% (10.32-14.5) | 2.81% (1.28-4.47) | 3.76% (1.64-6.8) |
| **1917** | Hispanic | 14.98% (13.05-16.91) | 15.45% (13.46-17.38) | 3.18% (1.75-4.72) | 7.79% (4.22-14.25) |
| **1918** | Hispanic | 22.72% (20.57-24.73) | 23.21% (21.01-25.33) | 2.15% (0.86-3.47) | -25.73% (-197.77-229.64) |
| **1919** | Hispanic | 22.08% (20.4-23.78) | 22.62% (20.94-24.35) | 2.48% (1.29-3.81) | -41.11% (-352.64-446.09) |
| **1920** | Hispanic | 29.44% (27.56-31.42) | 29.81% (27.89-31.78) | 1.26% (0.19-2.53) | -4.32% (-9.05--0.64) |

| | | | | | |
|---|---|---|---|---|---|
| **1921** | Hispanic | 30.48% (28.6-32.42) | 30.92% (29.01-32.98) | 1.45% (0.34-2.56) | -4.75% (-8.42--1.09) |
| **1922** | Hispanic | 33.73% (31.75-35.69) | 33.93% (32-35.88) | 0.58% (-0.51-1.59) | -1.53% (-4.21-1.32) |
| **1923** | Hispanic | 26.32% (24.6-28.01) | 26.64% (24.95-28.28) | 1.22% (0.04-2.41) | -6.1% (-13.09--0.23) |
| **1924** | Hispanic | 25.93% (24.21-27.48) | 26.22% (24.54-27.7) | 1.12% (0.11-2.17) | -6.08% (-12.99--0.55) |
| **1925** | Hispanic | 21.95% (20.45-23.33) | 22.17% (20.68-23.55) | 0.98% (-0.23-2.22) | -17.3% (-247.75-300.88) |
| **1926** | Hispanic | 22.39% (20.9-23.79) | 22.78% (21.28-24.21) | 1.73% (0.53-3.01) | -26.06% (-155.65-175.07) |
| **1927** | Hispanic | 17.83% (16.61-19.06) | 18.06% (16.79-19.24) | 1.29% (0.07-2.57) | 6.94% (0.42-15.79) |
| **1928** | Hispanic | 21.44% (20.08-22.86) | 21.77% (20.45-23.15) | 1.58% (0.43-2.84) | -32.14% (-810.64-733.31) |
| **1929** | Hispanic | 25.1% (23.52-26.74) | 25.45% (23.87-27.07) | 1.39% (0.2-2.58) | -8.63% (-18.94--1.19) |
| **1930** | Hispanic | 24.2% (22.69-25.84) | 24.52% (22.97-26.16) | 1.33% (0.15-2.59) | -10.51% (-25.82--1.11) |
| **1931** | Hispanic | 20.64% (19.21-22.15) | 20.96% (19.52-22.42) | 1.57% (0.4-2.99) | 29.06% (-592.69-707.11) |
| **1932** | Hispanic | 22.11% (20.72-23.58) | 22.25% (20.92-23.63) | 0.66% (-0.67-1.82) | -11.62% (-197.97-79.39) |
| **1933** | Hispanic | 20.71% (19.38-22.1) | 21.01% (19.66-22.35) | 1.42% (0.01-2.75) | 27.23% (-346.55-500.41) |
| **1934** | Hispanic | 23.78% (22.3-25.29) | 23.77% (22.34-25.32) | -0.03% (-1.2-1.21) | 0.17% (-12.09-12.85) |
| **1935** | Hispanic | 22.65% (21.24-24.02) | 22.63% (21.26-24.02) | -0.1% (-1.39-1.27) | 0.87% (-31.88-29.24) |
| **1936** | Hispanic | 22.31% (20.9-23.75) | 22.22% (20.86-23.59) | -0.41% (-1.68-0.97) | 6.87% (-51.68-72.28) |
| **1937** | Hispanic | 21.86% (20.44-23.3) | 21.71% (20.33-23.08) | -0.65% (-1.93-0.71) | 13.48% (-82.42-170.75) |

| | | | | | |
|---|---|---|---|---|---|
| **1938** | Hispanic | 21.92% (20.47-23.36) | 21.76% (20.37-23.17) | -0.72% (-2.05-0.65) | 13.9% (-188.12-179.22) |
| **1939** | Hispanic | 22.32% (20.88-23.75) | 22.07% (20.66-23.5) | -1.1% (-2.44-0.25) | 18.02% (-71.39-101.9) |
| **1940** | Hispanic | 21.04% (19.63-22.52) | 20.99% (19.62-22.45) | -0.21% (-1.64-1.13) | 2.63% (-221.54-247.27) |
| **1941** | Hispanic | 17.15% (15.96-18.4) | 17.1% (15.88-18.33) | -0.26% (-1.77-1.25) | -1.22% (-9.61-5.62) |
| **1942** | Hispanic | 26.61% (24.97-28.35) | 26.32% (24.74-27.94) | -1.1% (-2.46-0.14) | 5.32% (-0.69-12.13) |
| **1943** | Hispanic | 24.07% (22.6-25.69) | 23.92% (22.43-25.49) | -0.63% (-1.83-0.6) | 4.99% (-5.49-16.44) |
| **1944** | Hispanic | 24.28% (22.84-25.76) | 24.22% (22.77-25.68) | -0.27% (-1.49-0.95) | 2.02% (-7.17-11.97) |
| **1945** | Hispanic | 21.04% (19.74-22.4) | 21.04% (19.74-22.4) | 0% (0-0) | 0% (0-0) |
| **1946** | Hispanic | 24.09% (22.71-25.62) | 23.97% (22.59-25.42) | -0.49% (-1.76-0.74) | 3.96% (-6.17-14.82) |
| **1947** | Hispanic | 21.41% (20.14-22.81) | 21.48% (20.3-22.86) | 0.31% (-1.02-1.66) | -0.5% (-314.82-272.48) |
| **1948** | Hispanic | 23.89% (22.61-25.26) | 23.91% (22.7-25.27) | 0.08% (-1.13-1.36) | -0.32% (-14.4-10.88) |
| **1949** | Hispanic | 22.97% (21.65-24.39) | 22.9% (21.67-24.32) | -0.3% (-1.65-1.09) | 3.66% (-18.36-21.58) |
| **1950** | Hispanic | 22.81% (21.56-24.13) | 22.72% (21.47-24.12) | -0.4% (-1.69-0.95) | 5.44% (-19.58-31.34) |
| **1951** | Hispanic | 19.46% (18.31-20.78) | 19.53% (18.35-20.75) | 0.34% (-0.96-1.77) | 3.86% (-16.38-37.03) |
| **1952** | Hispanic | 19.68% (18.45-20.93) | 19.78% (18.6-21.06) | 0.49% (-0.79-1.81) | 6.37% (-26.91-53.02) |
| **1953** | Hispanic | 20.3% (19.08-21.61) | 20.38% (19.23-21.58) | 0.41% (-0.89-1.84) | 9.25% (-190.99-189.83) |
| **1954** | Hispanic | 25.13% (23.81-26.58) | 25.06% (23.76-26.45) | -0.28% (-1.46-0.97) | 1.75% (-6.41-9.05) |

| | | | | | |
|---|---|---|---|---|---|
| **1955** | Hispanic | 25.6% (24.23-27.13) | 25.47% (24.1-26.91) | -0.49% (-1.79-0.91) | 2.91% (-5.7-10.1) |
| **1956** | Hispanic | 23.37% (22.04-24.79) | 23.23% (21.96-24.55) | -0.6% (-1.95-0.62) | 5.79% (-7.55-21.44) |
| **1957** | Hispanic | 19.51% (18.31-20.79) | 19.34% (18.19-20.57) | -0.88% (-2.22-0.43) | -11.27% (-70.88-10.31) |
| **1958** | Hispanic | 21.53% (20.29-22.87) | 21.33% (20.12-22.6) | -0.9% (-2.09-0.52) | 19.81% (-481.39-241.47) |
| **1959** | Hispanic | 30.94% (29.43-32.59) | 30.29% (28.8-31.85) | -2.1% (-3.18--0.92) | 6.6% (2.86-10.04) |
| **1960** | Hispanic | 24.28% (22.79-25.76) | 23.75% (22.34-25.17) | -2.14% (-3.38--0.96) | 16.28% (6.9-30.78) |
| **1961** | Hispanic | 22.37% (21.08-23.78) | 21.88% (20.57-23.24) | -2.19% (-3.57--0.76) | 36.27% (-1.49-185.84) |
| **1962** | Hispanic | 25.32% (23.97-26.87) | 24.69% (23.34-26.15) | -2.47% (-3.71--1.13) | 14.44% (6.93-24.43) |
| **1963** | Hispanic | 20.9% (19.77-22.17) | 20.51% (19.43-21.69) | -1.86% (-3.18--0.44) | -30.53% (-784.36-722.19) |
| **1964** | Hispanic | 19.61% (18.54-20.79) | 19.17% (18.11-20.27) | -2.28% (-3.76--0.96) | -30.49% (-185.24--6.58) |
| **1965** | Hispanic | 22.28% (20.85-23.7) | 21.78% (20.37-23.19) | -2.26% (-3.64--0.87) | 36.9% (-260.34-296) |
| **1966** | Hispanic | 25.17% (23.21-27.26) | 24.49% (22.59-26.42) | -2.73% (-4.11--1.48) | 16.55% (8.82-33.88) |
| **1916** | White | 16.74% (14.48-18.93) | 17.36% (15.02-19.58) | 3.74% (2.21-5.14) | -11.42% (-19.43--6.22) |
| **1917** | White | 16.16% (14.31-17.89) | 16.76% (14.84-18.57) | 3.71% (2.18-5.14) | -12.28% (-19.86--7.07) |
| **1918** | White | 18.21% (16.41-19.98) | 18.83% (16.94-20.71) | 3.41% (2.05-4.84) | -8.9% (-13.52--5.24) |
| **1919** | White | 13.09% (11.84-14.43) | 13.61% (12.24-14.99) | 3.95% (2.52-5.43) | -28.66% (-78.37--13.73) |
| **1920** | White | 15.53% (14.03-17.04) | 16.04% (14.49-17.54) | 3.29% (1.6-4.78) | -12.06% (-20.42--5.35) |

| | | | | | |
|---|---|---|---|---|---|
| **1921** | White | 14.16% (12.75-15.58) | 14.61% (13.16-16.07) | 3.17% (1.55-4.83) | -15.42% (-31.98--7.05) |
| **1922** | White | 16.33% (14.79-17.92) | 16.73% (15.13-18.31) | 2.5% (0.92-3.95) | -8.14% (-14.05--2.78) |
| **1923** | White | 12.73% (11.51-14.02) | 13.11% (11.84-14.42) | 3.03% (1.45-4.81) | -25.75% (-103.97--7.93) |
| **1924** | White | 15.13% (13.87-16.45) | 15.5% (14.1-16.88) | 2.46% (0.8-4.13) | -9.73% (-18.54--3.41) |
| **1925** | White | 13.07% (11.82-14.25) | 13.4% (12.12-14.6) | 2.48% (0.79-4.15) | -18.08% (-52.25--5.17) |
| **1926** | White | 15.52% (14.14-16.91) | 15.81% (14.39-17.22) | 1.82% (0.31-3.37) | -6.57% (-13.61--1.08) |
| **1927** | White | 12.21% (11.1-13.36) | 12.49% (11.35-13.66) | 2.27% (0.62-3.82) | -28% (-145.6-90.28) |
| **1928** | White | 16.47% (15.13-17.95) | 16.68% (15.26-18.1) | 1.25% (-0.29-2.91) | -4.01% (-9.26-0.96) |
| **1929** | White | 17.84% (16.35-19.42) | 18.01% (16.54-19.63) | 0.95% (-0.65-2.55) | -2.57% (-7.28-1.89) |
| **1930** | White | 16.32% (15.06-17.73) | 16.47% (15.1-17.9) | 0.94% (-0.56-2.51) | -2.94% (-8.49-1.9) |
| **1931** | White | 13.94% (12.69-15.16) | 14.08% (12.83-15.34) | 1.05% (-0.6-2.74) | -5.13% (-17.64-2.98) |
| **1932** | White | 14.47% (13.33-15.69) | 14.59% (13.4-15.86) | 0.84% (-0.92-2.56) | -3.73% (-12.64-4.32) |
| **1933** | White | 12.25% (11.18-13.4) | 12.39% (11.3-13.53) | 1.08% (-0.61-2.88) | -12.88% (-105.9-34.41) |
| **1934** | White | 12.35% (11.24-13.48) | 12.49% (11.36-13.64) | 1.11% (-0.62-2.79) | -12.51% (-94.82-14.19) |
| **1935** | White | 12.39% (11.29-13.53) | 12.49% (11.34-13.63) | 0.8% (-0.83-2.34) | -8.2% (-81.71-17.21) |
| **1936** | White | 13.12% (11.98-14.34) | 13.17% (12.02-14.41) | 0.44% (-1.18-2.06) | -3.1% (-18.94-10.23) |
| **1937** | White | 12.47% (11.37-13.65) | 12.54% (11.46-13.65) | 0.59% (-1-2.28) | -5.6% (-44.19-18.79) |

| | | | | | |
|---|---|---|---|---|---|
| **1938** | White | 12.31% (11.24-13.44) | 12.38% (11.28-13.51) | 0.56% (-1.03-2.24) | -6.2% (-71.26-39.51) |
| **1939** | White | 15.34% (14.05-16.65) | 15.33% (14.06-16.58) | -0.05% (-1.8-1.69) | 0.16% (-6.8-7.26) |
| **1940** | White | 13.48% (12.31-14.64) | 13.46% (12.28-14.67) | -0.11% (-1.81-1.6) | 0.7% (-12.27-12.59) |
| **1941** | White | 10.49% (9.47-11.46) | 10.58% (9.57-11.58) | 0.84% (-0.91-2.62) | 10.14% (-53.65-114.43) |
| **1942** | White | 18.76% (17.24-20.33) | 18.59% (17.11-20.15) | -0.89% (-2.4-0.61) | 2.25% (-1.56-6.08) |
| **1943** | White | 14.3% (13.12-15.57) | 14.24% (13.01-15.52) | -0.44% (-1.98-1.2) | 2.14% (-5.75-10.81) |
| **1944** | White | 14.65% (13.49-16.02) | 14.54% (13.34-15.87) | -0.77% (-2.3-0.75) | 3.37% (-3.35-10.36) |
| **1945** | White | 11.25% (10.26-12.35) | 11.25% (10.26-12.35) | 0% (0-0) | 0% (0-0) |
| **1946** | White | 13.67% (12.5-14.97) | 13.58% (12.37-14.82) | -0.72% (-2.37-0.7) | 3.99% (-4.52-15.2) |
| **1947** | White | 13.41% (12.26-14.65) | 13.27% (12.1-14.45) | -1.06% (-2.64-0.62) | 6.54% (-3.94-21.36) |
| **1948** | White | 15.63% (14.37-17.11) | 15.41% (14.18-16.73) | -1.37% (-3.05-0.32) | 4.82% (-1.18-11.1) |
| **1949** | White | 15.37% (14.18-16.78) | 15.11% (13.86-16.44) | -1.66% (-3.4--0.17) | 6.08% (0.58-13.24) |
| **1950** | White | 16.61% (15.3-18.11) | 16.27% (15-17.65) | -2.05% (-3.78--0.42) | 6.3% (1.34-12.31) |
| **1951** | White | 13.55% (12.46-14.73) | 13.3% (12.21-14.56) | -1.84% (-3.54--0.06) | 10.97% (0.39-25.27) |
| **1952** | White | 14.68% (13.54-16.01) | 14.41% (13.31-15.64) | -1.86% (-3.6--0.1) | 7.8% (0.48-16.35) |
| **1953** | White | 14.74% (13.64-16.02) | 14.46% (13.39-15.64) | -1.92% (-3.38--0.28) | 8% (1.26-16.02) |
| **1954** | White | 20.52% (19.08-22.05) | 19.96% (18.61-21.42) | -2.68% (-4.25--1.15) | 5.94% (2.71-9.41) |

| | | | | | |
|---|---|---|---|---|---|
| **1955** | White | 19.03% (17.65-20.45) | 18.53% (17.21-19.97) | -2.63% (-4.24--1.27) | 6.3% (3.08-10.49) |
| **1956** | White | 15.66% (14.45-16.99) | 15.28% (14.13-16.58) | -2.37% (-4--0.89) | 8.33% (3.28-14.65) |
| **1957** | White | 13.12% (12.12-14.27) | 12.87% (11.88-13.99) | -1.91% (-3.6--0.09) | 13.55% (0.4-32.47) |
| **1958** | White | 15.45% (14.29-16.85) | 15.06% (13.93-16.38) | -2.47% (-4.12--0.98) | 9.12% (3.52-16.82) |
| **1959** | White | 24.18% (22.46-26.1) | 23.4% (21.73-25.24) | -3.2% (-4.63--1.7) | 5.94% (3.29-8.9) |
| **1960** | White | 17.94% (16.52-19.46) | 17.32% (15.93-18.8) | -3.47% (-5.21--2) | 9.37% (5.28-14.66) |
| **1961** | White | 16.77% (15.48-18.11) | 16.16% (14.91-17.55) | -3.6% (-5.26--1.83) | 11.1% (6.01-16.86) |
| **1962** | White | 18.93% (17.48-20.51) | 18.16% (16.87-19.63) | -4.1% (-5.71--2.51) | 10.16% (6.28-14.33) |
| **1963** | White | 15.66% (14.52-16.85) | 15.01% (13.97-16.18) | -4.11% (-5.62--2.51) | 14.76% (9.08-21.15) |
| **1964** | White | 15.33% (14.22-16.57) | 14.63% (13.55-15.77) | -4.53% (-6.17--2.87) | 17.09% (10.92-24.7) |
| **1965** | White | 18.65% (17.09-20.26) | 17.66% (16.08-19.13) | -5.33% (-7.05--3.64) | 13.48% (9-18.4) |
| **1966** | White | 22.5% (20.27-24.9) | 21.16% (18.85-23.42) | -5.96% (-7.63--4.38) | 11.92% (8.37-16.15) |

#### Section 3: Sensitivity Analysis

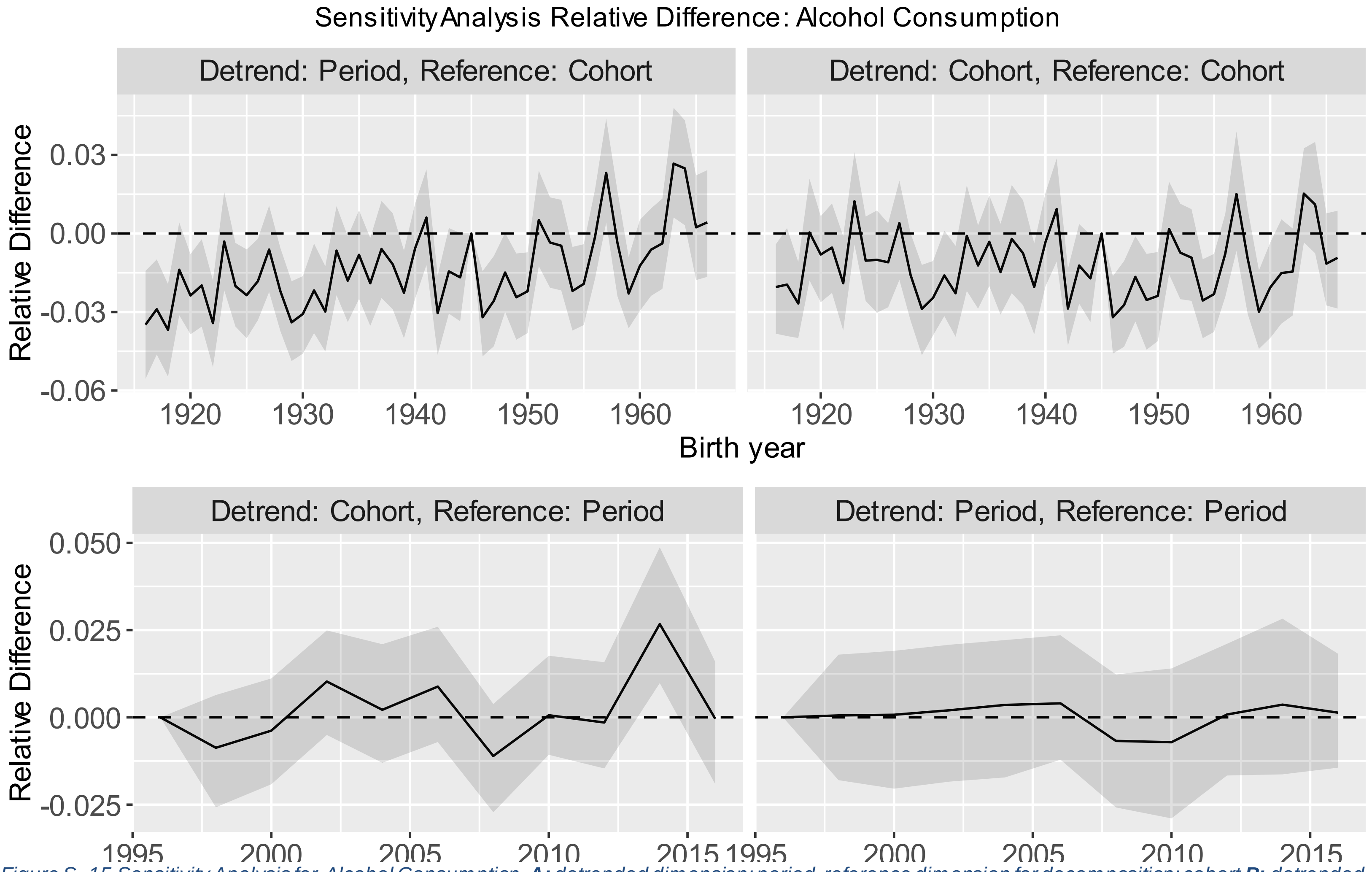

*Figure S. 15 Sensitivity Analysis for Alcohol Consumption.* ***A:*** *detrended dimension: period, reference dimension for decomposition: cohort* ***B:*** *detrended dimension: cohort, reference dimension for decomposition: cohort,* ***C:*** *detrended dimension: cohort, reference dimension for decomposition: period,* ***D:*** *detrended dimension: period, reference dimension for decomposition: period.*

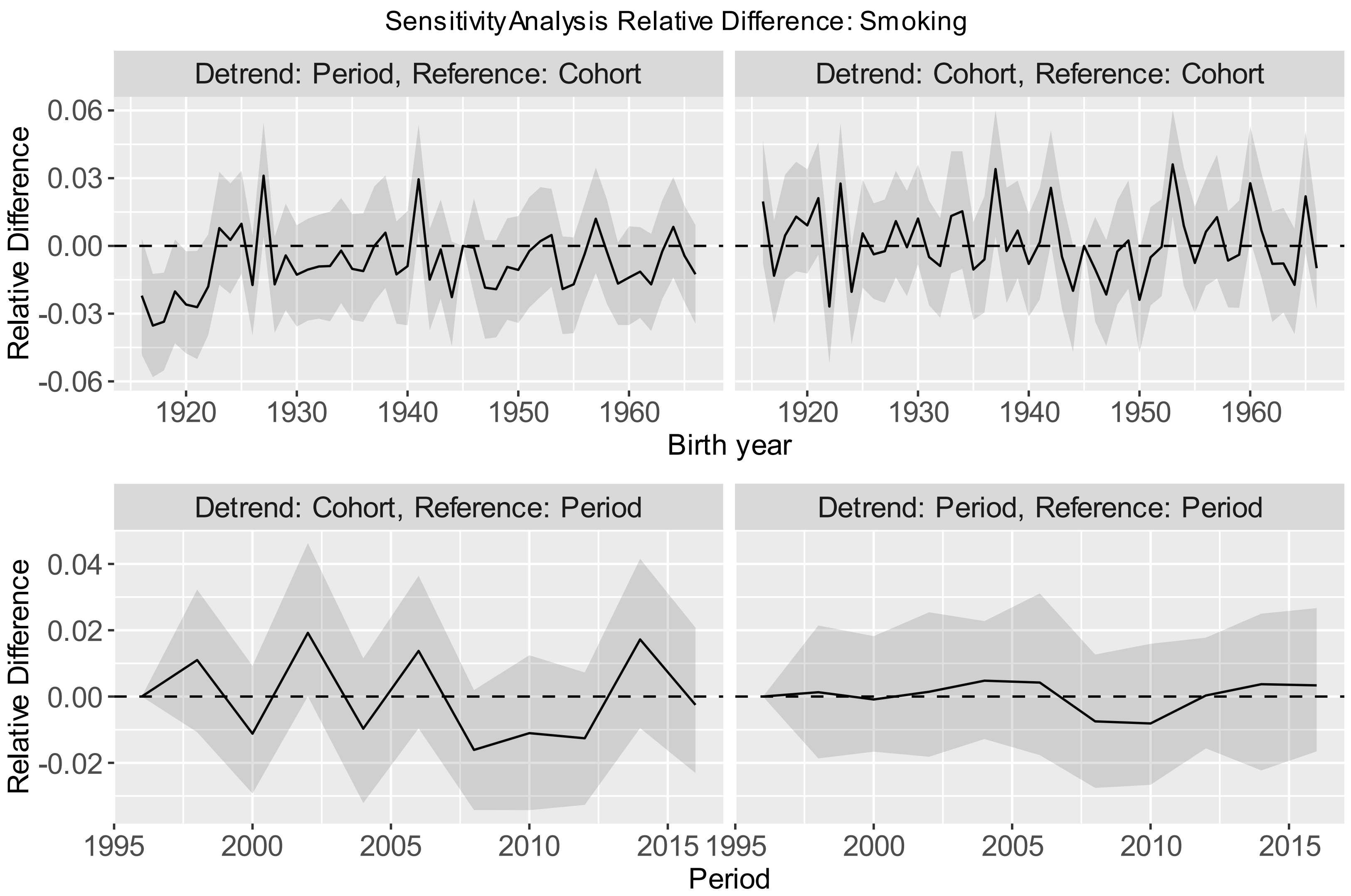

*Figure S. 16 Sensitivity Analysis for Smoking.* ***A:*** *detrended dimension: period, reference dimension for decomposition: cohort* ***B:*** *detrended dimension: cohort, reference dimension for decomposition: cohort,* ***C:*** *detrended dimension: cohort, reference dimension for decomposition: period,* ***D:*** *detrended dimension: period, reference dimension for decomposition: period.*

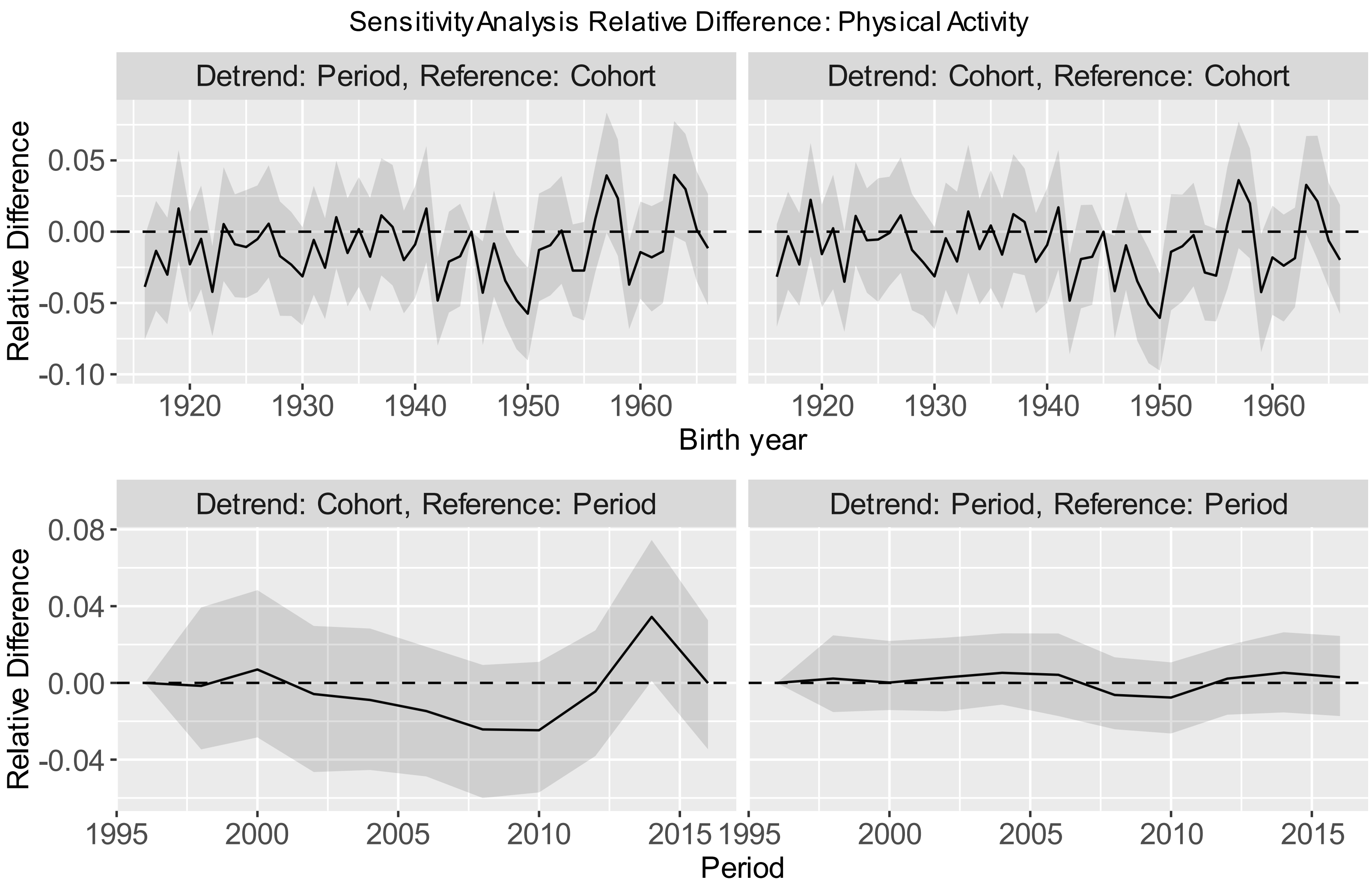

*Figure S. 17 Sensitivity Analysis for Physical Activity. **A:** detrended dimension: period, reference dimension for decomposition: cohort **B:** detrended dimension: cohort, reference dimension for decomposition: cohort, **C:** detrended dimension: cohort, reference dimension for decomposition: period, **D:** detrended dimension: period, reference dimension for decomposition: period.*

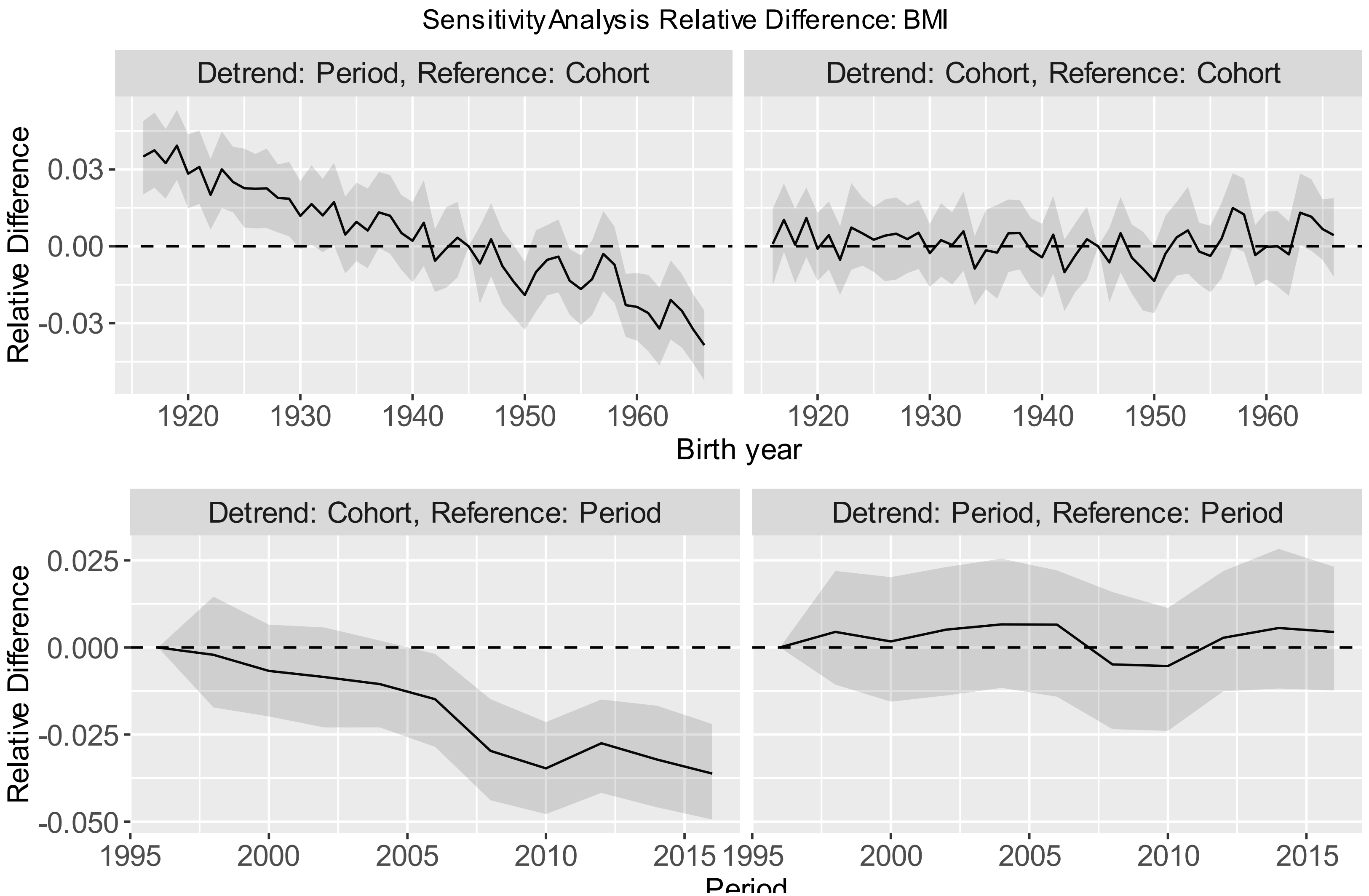

*Figure S. 18 Sensitivity Analysis for BMI.* ***A:*** *detrended dimension: period, reference dimension for decomposition: cohort* ***B:*** *detrended dimension: cohort, reference dimension for decomposition: cohort,* ***C:*** *detrended dimension: cohort, reference dimension for decomposition: period,* ***D:*** *detrended dimension: period, reference dimension for decomposition: period.*

#### Section 4: Additional Analysis

#### Section 4.1: Bootstrap Stability

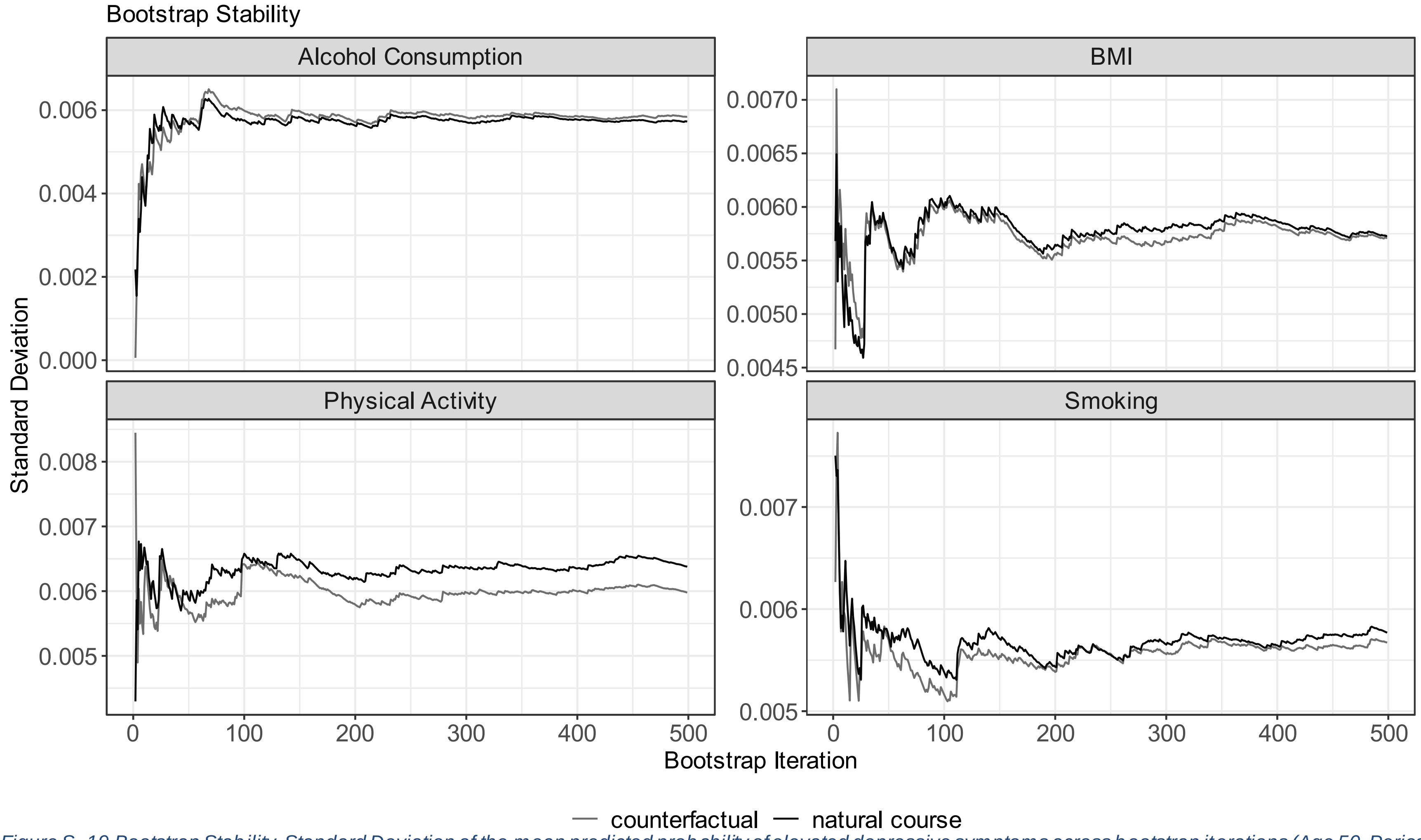

*Figure S. 19 Bootstrap Stability. Standard Deviation of the mean predicted probability of elevated depressive symptoms across bootstrap iterations (Age 50, Period 1996, Cohort 1960). Results are presented by mediator for the natural course and counterfactual scenario. Maximum bootstrap iterations 499. Monte Carlo Iterations: 50.*

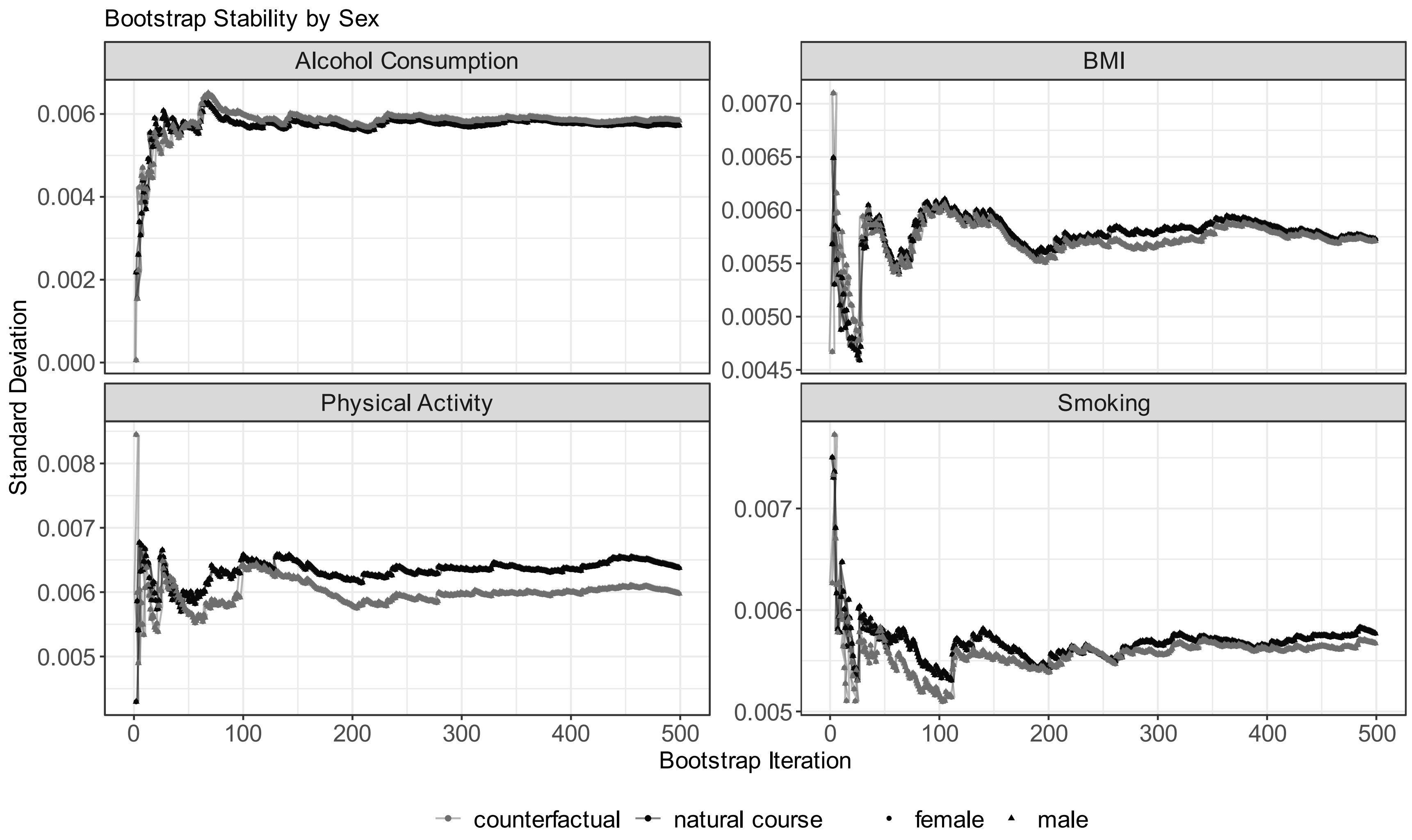

*Figure S. 20 Bootstrap Stability. Standard Deviation of the mean predicted probability of elevated depressive symptoms across bootstrap iterations (Age 50, Period 1996, Cohort 1960). Results are presented by mediator and sex for the natural course and counterfactual scenario. Maximum bootstrap iterations 499. Monte Carlo Iterations: 50*

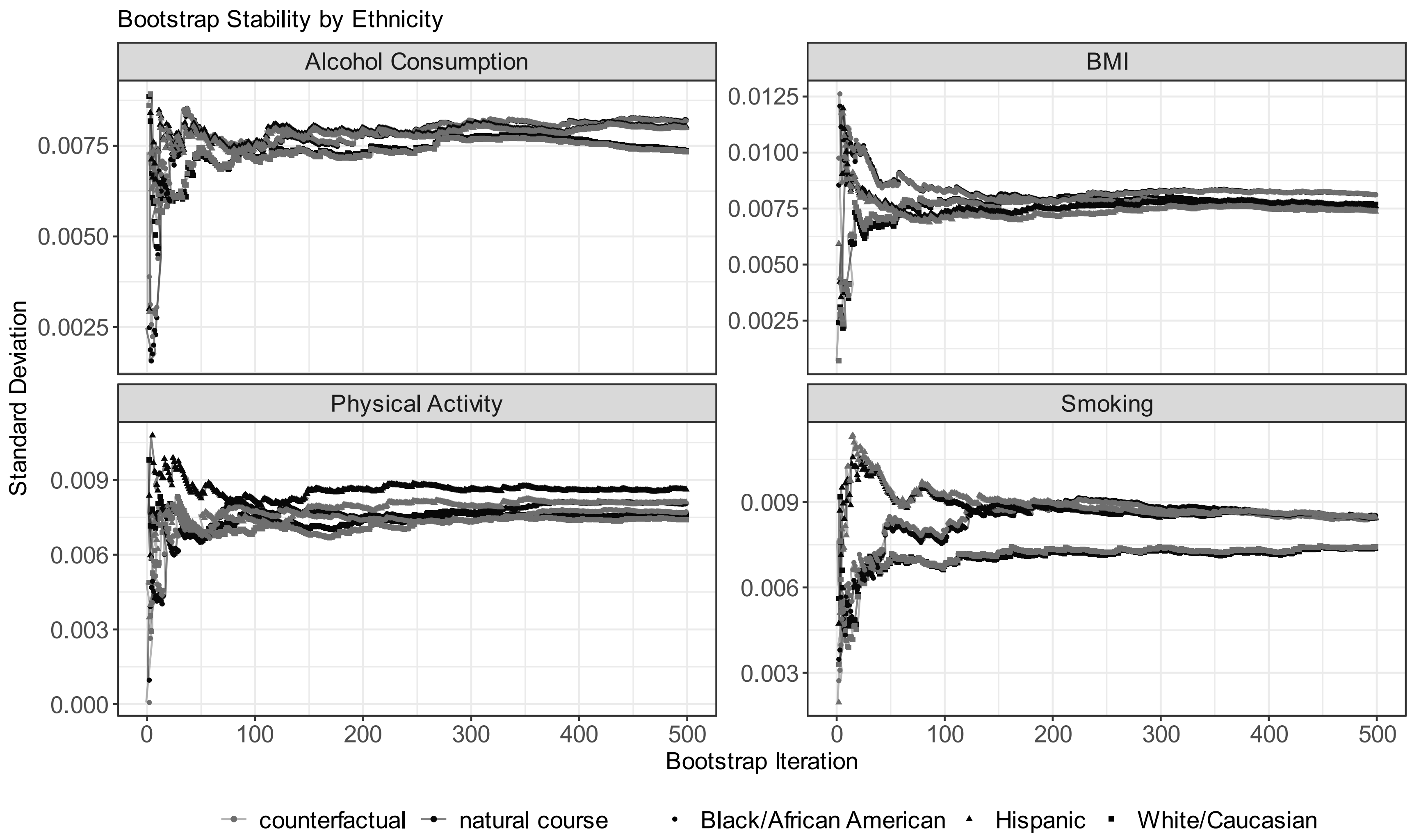

*Figure S. 21 Bootstrap Stability. Standard Deviation of the mean predicted probability of elevated depressive symptoms across bootstrap iterations (Age 50, Period 1996, Cohort 1960). Results are presented by mediator and race/ethnicity for the natural course and counterfactual scenario. Maximum bootstrap iterations 499. Monte Carlo Iterations: 50*

#### Section 4.2: Counterfactual Decomposition for Smoking as Categorical Variable

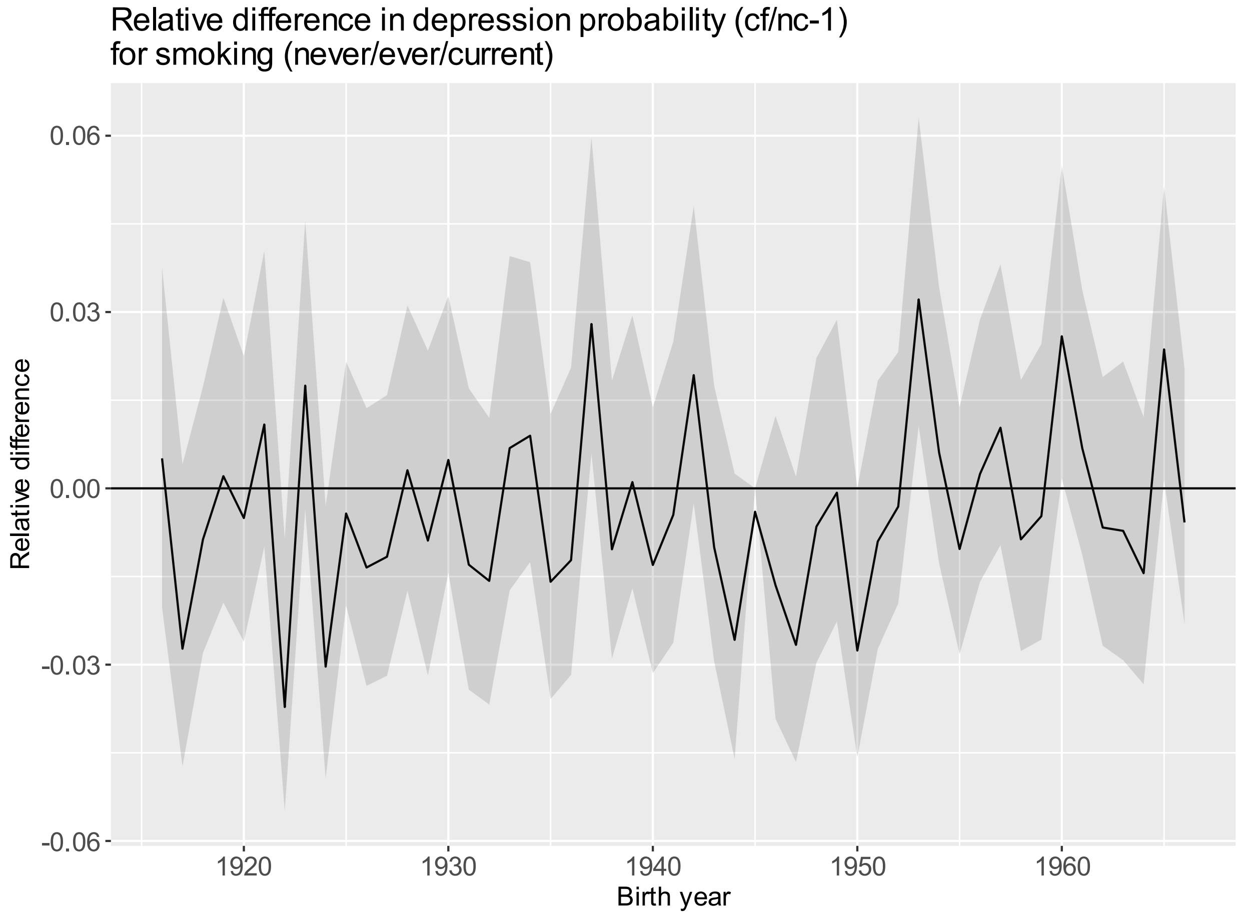

*Figure S. 22 Relative difference (95% CI) between the counterfactual and natural course estimates of probability of elevated depressive symptoms by birth cohort (cf/nc-1) for smoking (never/ever/current). Sample Size: 163,660.*

#### Section 4.3: Assessing the presence of panel attrition and panel conditioning

We performed two tests to assess whether panel attrition and/or panel conditioning are present in our study. We assessed the relationship between elevated depressive symptoms and time-in-sample with a logistic regression model and allowed for non-linearity through natural cubic splines. We adjusted the model for age, sex, race/ethnicity, education, birth cohort, BMI, physical activity, smoking, alcohol consumption and whether the participant responded in the next wave (Figure S.26). We find a non-linear negative association with populations that participated in the sample for more than four waves, showing a decreased probability of elevated depressive symptoms. In a second step, we selected populations that participated in one wave and compared them to populations that participated in two waves. After matching the populations based on all relevant characteristics, we find no difference in elevated depressive symptoms between the two groups (Table S.14). We conclude that there is evidence for panel attrition but not panel conditioning.

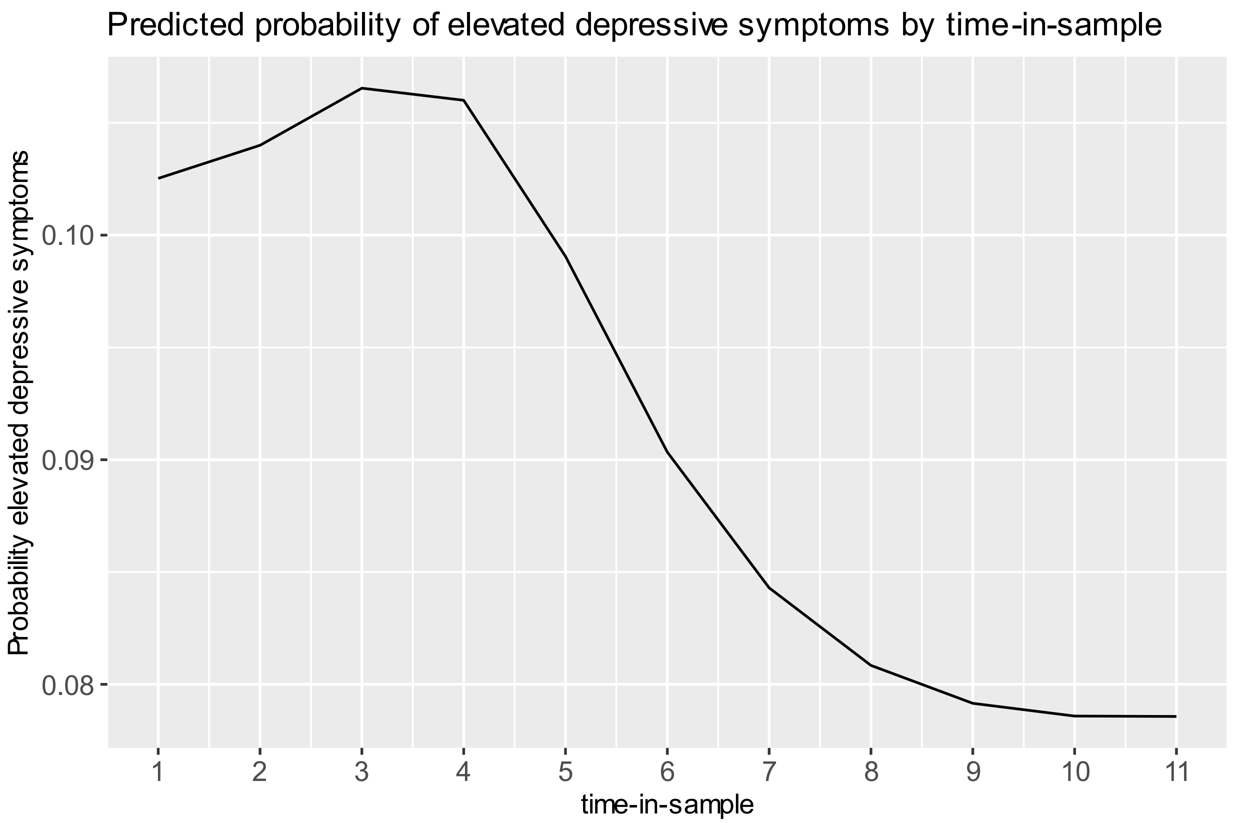

*Figure S. 23 Association between time-in-sample and predicted probability of elevated depressive symptoms. Other covariates are held constant.*

*Table S. 14 Levels of elevated depressive symptoms stratified by time-in-sample before and after matching.*

| | | **Before Matching** | | | **After Matching** | | |
|---|---|---|---|---|---|---|---|
| | | **Time-in-sample** | | | **Time-in-sample** | | |
| | | **1 wave** | **2 waves** | **p-value** | **1 wave** | **2 waves** | **p-value** |
| **N** | | 24670 | 1610 | | 1201 | 1201 | |
| elevated depressive symptoms = 1 N (%) | | 5362 (21.7) | 419 (26.0) | <0.001 | 311 (25.9) | 294 (24.5) | 0.452 |
| **Age (mean (SD))** | | 61.55 (8.12) | 60.13 (8.42) | **<0.001** | 60.50 (7.57) | 60.54 (8.64) | **0.900** |
| **Birth Cohort (in 2-3-year intervals) N (%)** | | | | **<0.001** | | | **1.000** |
| | 1916-1918 | 772 (3.1) | 14 (0.9) | | 12 (1.0) | 12 (1.0) | |
| | 1918-1921 | 728 (3.0) | 12 (0.7) | | 9 (0.7) | 9 (0.7) | |
| | 1921-1923 | 1342 (5.4) | 17 (1.1) | | 15 (1.2) | 15 (1.2) | |
| | 1923-1926 | 903 (3.7) | 85 (5.3) | | 68 (5.7) | 68 (5.7) | |
| | 1926-1928 | 1380 (5.6) | 128 (8.0) | | 102 (8.5) | 102 (8.5) | |
| | 1928-1930 | 927 (3.8) | 100 (6.2) | | 77 (6.4) | 77 (6.4) | |
| | 1930-1933 | 1282 (5.2) | 31 (1.9) | | 24 (2.0) | 24 (2.0) | |
| | 1933-1935 | 1978 (8.0) | 43 (2.7) | | 39 (3.2) | 39 (3.2) | |
| | 1935-1938 | 1428 (5.8) | 37 (2.3) | | 31 (2.6) | 31 (2.6) | |
| | 1938-1940 | 2288 (9.3) | 58 (3.6) | | 48 (4.0) | 48 (4.0) | |

| | | | | | | | |
|---|---|---|---|---|---|---|---|
| | 1940-1942 | 1438 (5.8) | 56 (3.5) | | 42 (3.5) | 42 (3.5) | |
| | 1942-1945 | 1026 (4.2) | 64 (4.0) | | 46 (3.8) | 46 (3.8) | |
| | 1945-1947 | 1347 (5.5) | 106 (6.6) | | 80 (6.7) | 80 (6.7) | |
| | 1947-1950 | 1095 (4.4) | 109 (6.8) | | 73 (6.1) | 73 (6.1) | |
| | 1950-1952 | 1774 (7.2) | 190 (11.8) | | 151 (12.6) | 151 (12.6) | |
| | 1952-1954 | 1252 (5.1) | 162 (10.1) | | 108 (9.0) | 108 (9.0) | |
| | 1954-1957 | 1323 (5.4) | 136 (8.4) | | 91 (7.6) | 91 (7.6) | |
| | 1957-1959 | 1820 (7.4) | 215 (13.4) | | 167 (13.9) | 167 (13.9) | |
| | 1959-1962 | 448 (1.8) | 31 (1.9) | | 17 (1.4) | 17 (1.4) | |
| | 1962-1964 | 119 (0.5) | 16 (1.0) | | 1 (0.1) | 1 (0.1) | |
| **Period (mean (SD))** | | 2001.40 (6.41) | 2004.81 (5.98) | **<0.001** | 2004.31 (6.36) | 2004.49 (5.94) | **0.466** |
| **Sex = Female N (%)** | | 13844 (56.1) | 712 (44.2) | **<0.001** | 578 (48.1) | 578 (48.1) | **1.000** |
| **Education level N (%)** | | | | **0.004** | | | **1.000** |
| | Lt High-school | 5501 (22.3) | 405 (25.2) | | 288 (24.0) | 288 (24.0) | |
| | GED | 1207 (4.9) | 84 (5.2) | | 40 (3.3) | 40 (3.3) | |
| | High-school graduate | 7274 (29.5) | 451 (28.0) | | 346 (28.8) | 346 (28.8) | |
| | Some college | 5649 (22.9) | 391 (24.3) | | 293 (24.4) | 293 (24.4) | |

| | | | | | | | |
|---|---|---|---|---|---|---|---|
| | College and above | 5039 (20.4) | 279 (17.3) | | 234 (19.5) | 234 (19.5) | |
| **Race/ethnicity N (%)** | | | | **<0.001** | | | **1.000** |
| | White | 18684 (75.7) | 1066 (66.2) | | 899 (74.9) | 899 (74.9) | |
| | Black | 4402 (17.8) | 373 (23.2) | | 241 (20.1) | 241 (20.1) | |
| | Hispanic | 927 (3.8) | 90 (5.6) | | 35 (2.9) | 35 (2.9) | |
| | Other | 657 (2.7) | 81 (5.0) | | 26 (2.2) | 26 (2.2) | |
| **BMI N (%)** | | | | **0.003** | | | **1.000** |
| | underweight | 164 (0.7) | 20 (1.2) | | 3 (0.2) | 3 (0.2) | |
| | normal | 7602 (30.8) | 450 (28.0) | | 308 (25.6) | 308 (25.6) | |
| | overweight | 9680 (39.2) | 634 (39.4) | | 496 (41.3) | 496 (41.3) | |
| | obese | 7224 (29.3) | 506 (31.4) | | 394 (32.8) | 394 (32.8) | |
| **Alcohol Consumption N (%)** | | | | **<0.001** | | | **1.000** |
| | No or does not drink | 15800 (64.0) | 982 (61.0) | | 834 (69.4) | 834 (69.4) | |
| | Moderate drinker | 5190 (21.0) | 313 (19.4) | | 228 (19.0) | 228 (19.0) | |
| | Heavy drinker | 2937 (11.9) | 211 (13.1) | | 117 (9.7) | 117 (9.7) | |
| | Excessive drinker | 743 (3.0) | 104 (6.5) | | 22 (1.8) | 22 (1.8) | |
| **Current Smoker = Yes N (%)** | | 4645 (18.8) | 395 (24.5) | **<0.001** | 199 (16.6) | 199 (16.6) | **1.000** |

| | | | | | | |
|---|---|---|---|---|---|---|
| **Vigorous physical activity = yes N (%)** | 12127 (49.2) | 729 (45.3) | **0.003** | 557 (46.4) | 557 (46.4) | **1.000** |
